# The effects of weather and mobility on respiratory viruses dynamics before and after the COVID-19 pandemic

**DOI:** 10.1101/2023.03.28.23287799

**Authors:** Irma Varela-Lasheras, Lília Perfeito, Sara Mesquita, Joana Gonçalves-Sá

## Abstract

The flu season is caused by a combination of different pathogens, including influenza viruses (IVS), that cause the flu, and non-influenza respiratory viruses (NIRVs), that cause common colds or influenza-like illness. These viruses have similar circulation patterns, and weather has been considered a main driver of their dynamics, with peaks in the winter and almost no circulation during the summer in temperate regions. However, after the emergence of SARS-CoV2, in 2019, the dynamics of these respiratory viruses were strongly perturbed worldwide: some infections almost disappeared, others were delayed or occurred “off-season”. This disruption raised questions regarding the dominant role of weather while also providing an unique opportunity to investigate the relevance of different driving factors on the epidemiological dynamics of IVs and NIRVs, including viral interactions, non-pharmacological individual measures (such as masking), or mobility. Here, we use epidemiological surveillance data on several respiratory viruses from Canada and the USA from 2016 to 2023, and tested the effects of weather and mobility in their dynamics before and after the COVID-19 pandemic. Using statistical modelling, we found evidence that whereas in the pre-COVID-19 pandemic period, weather had a strong effect and mobility a limited effect on dynamics; in the post-COVID-19 pandemic period the effect of weather was strongly reduced and mobility played a more relevant role. These results, together with previous studies, indicate that at least some of the behavioral changes resulting from the non-pharmacological interventions implemented during COVID-19 pandemic had a strong effect on the dynamics of respiratory viruses. Furthermore, our results support the idea that these seasonal dynamics are driven by a complex system of interactions between the different factors involved, which probably led to an equilibrium that was disturbed, and perhaps permanently altered, by the COVID-19 pandemic.

## Introduction

The flu season causes about 3 to 5 million cases of severe illness and hundreds of thousands of deaths worldwide per year [1], posing a strong socioeconomic burden on societies and public health systems [2]. Despite being called “flu season”, influenza viruses (IVA and IVB) circulate in parallel with other non-influenza respiratory viruses (NIRVs), which frequently account for more than half of the influenza-like illness (ILI) cases in a season [3, 4]. Furthermore, NIRVs significantly contribute to the morbidity and mortality of this seasonal epidemic. For example, respiratory syncytial virus (RSV) is the predominant viral pathogen associated with acute lower respiratory infection in children younger than 5 [5]; common human coronaviruses (hCoVs) are responsible for 15-30% of upper respiratory track infections and can cause more severe disease in vulnerable groups (neonates, children, elderly, and in individuals with comorbidities) [6]; and metapneumovirus (hMPV) is a leading cause of acute respiratory infection particularly in those same groups [7, 8].

As mentioned above, IVs and NIRVs in temperate regions co-circulate and have similar seasonality patterns, tipycally peaking in the winter and almost disappearing during the summer [9]. In fact, their dynamics are often mimicked using different types of time series analysis and variations of compartmental models, such as the Susceptible-Infected-Recovered (SIR), with cyclic variation assured by exhausting/replenishing the number of susceptible individuals, by fragmenting populations, or by adding external cyclic parameters, such as weather [10–12]. Indeed, the effect of climate in viral circulation has been widely accepted as a main factor responsible for the observed seasonal dynamics in both hemispheres: first, there is biological evidence showing that weather conditions can influence virus survival [13–17], transmission efficiency [15, 16, 18], and host susceptibility [19, 20]; second, there is epidemiological evidence showing that weather conditions, particularly temperature and humidity, play an important role in IV epidemics [21–24], although there is an ongoing discussion regarding their relative roles (see [15, 16]); and third, adding weather variables often improves predictive models [21, 25–30].

However, the COVID-19 pandemic and the measures implemented to contain its spread have deeply perturbed IVs’ and NIRVs’ dynamics: not only the circulation of IVs and NIRVs was reduced to a great extent, which could be explained by the great reduction in physical contact, but, in some cases, and quite unexpectedly, their seasonal patterns were also altered [31–34], with local epidemics appearing “off-season”. Naturally, weather was never presented as the only responsible for seasonal epidemics, and other commonly identified drivers of seasonal patterns include viral interactions and human behaviour [35,36]. Viral interactions, either positive (synergistic) or negative (antagonistic), have been shown to occur between respiratory viruses, with potential implications for their epidemiological dynamics. Positive interactions seem less common [37], but evidence for negative interactions exist both from epidemiological and from experimental studies, either from reduced co-infection rates in patients or from reduced shedding or viral replication in animal and cellular models [38]. There is evidence for negative interactions between IVs, between IVA, RSV and rhinovirus, between RSV and hMPV, among others [38]. Regarding behaviour, it can impact infection numbers in at least three different ways: it may influence transmission rates through the frequency and type of social contacts (e.g. holidays, school periods, international traveling, etc) [39–43], through health-related behaviours (e.g., masking, isolation, hygiene practices, etc. [44, 45]), or at the susceptibility level, through vaccination acceptance or refusal [46].

Understanding the contribution of these different factors to the dynamics of respiratory viruses becomes even more relevant as they are changing at a global scale with climate change, newly emerging viruses, and changes in relevant human behaviours (e.g., large-scale travelling or mask adoption).

However, as these drivers do not exist in isolation (different population structures, weather, viral interactions, and behaviour can influence each other) and often co-occur, disentangling between their relative contributions is a complex problem. In particular, there is a strong link between weather and human behaviour, with the first strongly influencing the second. For example, outdoor weather conditions influence the amount of time spent indoors [47] and therefore can potentially have an effect on both viral viability, and social contact rates or length [43, 48].

Here, we argue that the COVID-19 pandemic and the subsequent disruption in behavioural patterns, offer an unique opportunity to test the relative importance of these different driving factors to the epidemiological dynamics of IVs and NIRVs.

To this end, we took advantage of the data publicly available from epidemiological surveillance systems on respiratory viruses, in the USA and in Canada, and tested the effects of weather and of mobility (as a relevant component of behaviour), in viral dynamics before and after the onset of the COVID-19 pandemic. By comparing the contribution of weather and mobility during these two periods, we were able to better understand their contributions and analyse their respective weights pre- and post-COVID-19 pandemic.

## Materials and methods

### Data Sources, Definitions, and Data Processing

#### Epidemiological data

*Viral positivity rates* (number of positive tests for a given virus divided by the total number of tests for that virus) were collected for the USA and Canada, at nation-wide levels, for IVA, IVB, and all available NIRVs common in both countries. The two countries collect data differently and some harmonization was required to allow comparison. For the USA, human coronaviruses (hCOVs), respiratory syncitial virus (RSV) and human metapneumovirus(hMPV) data was collected from The National Respiratory and Enteric Virus Surveillance System (NREVSS, CDC), from July 2016 until January 2023. The Human Coronavirus positivity rates are reported for the 4 variants separately (i.e. CoVHKU1, CoVNL63, CoVOC43 and CoV229E), so assuming the tests are done in parallel (through multiplexing) and that there are no or few co-infections, these values were summed to obtain the positivity rate for hCOVs. For IVs, the data source was FluView (CDC) [49], [50], from October 2015 until January 2023. For Canada, data was collected from The Respiratory Virus Detection Surveillance System (FluWatch, CIRID) for the same viruses [51], from September 2016 until January 2023. For both countries, the defined pre-COVID-19 pandemic period starts at data collection and ends on the week of 07^*th*^ March, 2020, and the COVID-19 pandemic period starts on the following week, of the 14^*th*^ March, 2020, to include the date when the COVID-19 pandemic was declared by the WHO (see Fig. 1).

**Fig 1.**
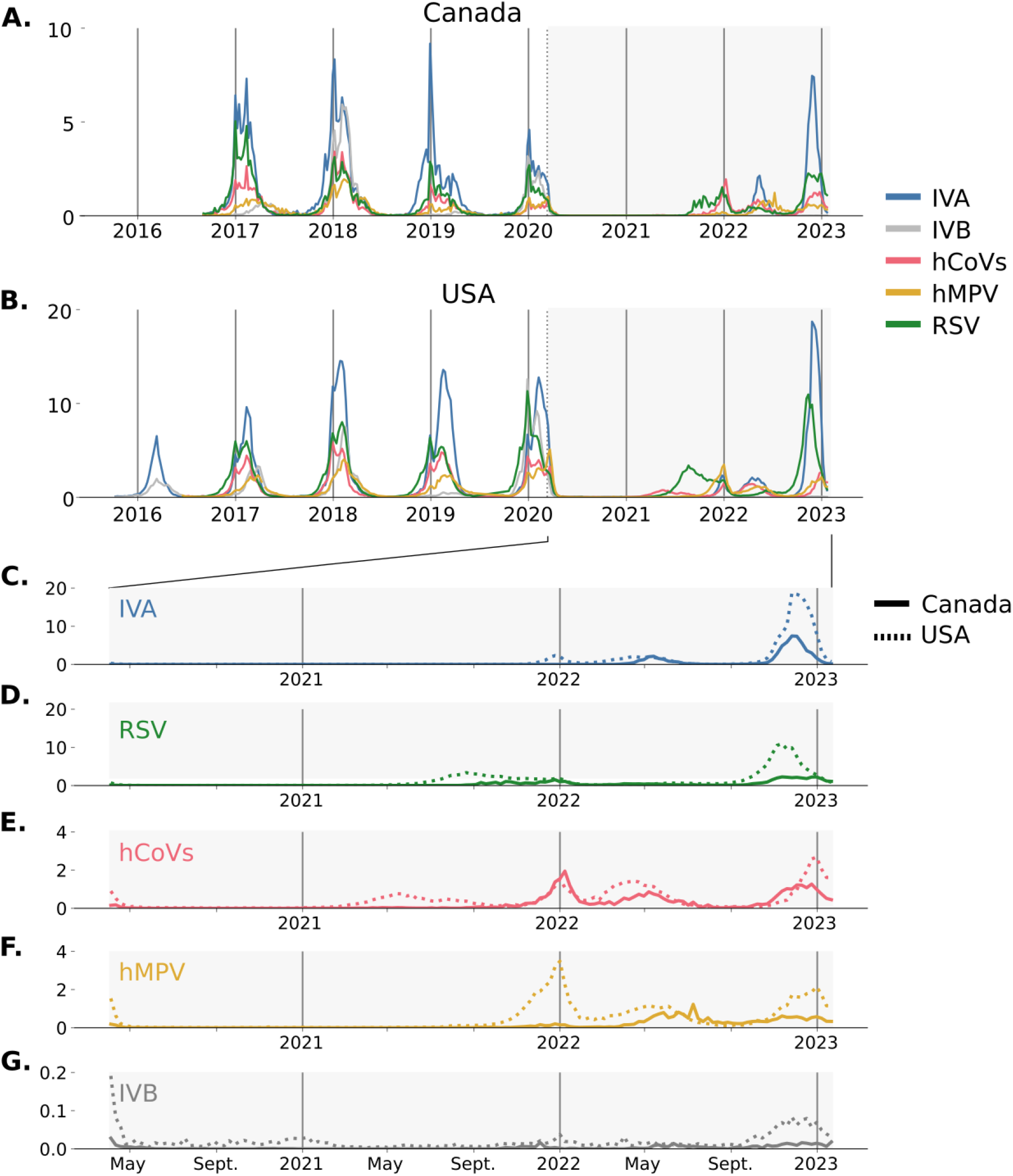
Incidence of different respiratory viruses in Canada and USA. (A) and (B) Incidence for influenza A (IVA), influenza B (IVB), common human coronaviruses (hCoVs), human metapneumovirus (hMPV) and respiratory syncitial virus (RSV) for Canada and the USA, respectively. Solid grey vertical lines, 1st January; grey shaded area, complete post-COVID pandemic period, from WHO pandemic declaration in March 11th, 2020, until January 2023. Y axes have different scales for Canada and the USA. (C) to (G) Incidence of IVA, IVB, hCoVs, HMPV and RSV in both countries in the post-COVID-19 pandemic period (starting at April 1st, 2020). Solid lines, incidence for Canada; dotted lines, incidence for the USA. Y axes have different scales for the different viruses.

*ILI outpatient rates* (number of outpatients visits to sentinel physicians due to ILI divided by the total number of visits) were collected using weekly, nation-wide ILI data from the U.S. Outpatient Influenza-like Illness Surveillance Network (ILINet, CDC) in the case of USA [52] and from the Syndromic/Influenza-like Illness Surveillance (FluWatch, CIRID) in the case of Canada [53].

*Incidence* was defined as the product of the weekly virus-specific positivity rate and the weekly ILI-outpatient rate, per 1000 people, and this rate is referred to by *incidence*, throughout the text. This proxy is regarded as the best relative measure of incidence that can be calculated from surveillance data, even though it involves a series of assumptions which are often not completely met (see [54] for details). Because Canada reports the total number of tests performed for each virus, a different proxy for incidence was calculated by dividing the total number of positive tests for each virus by the average Canadian population between 2017 and 2023. The correlations between these two incidence proxies were high (S1 Fig), supporting the idea that the product of the positivity rates and the ILI outpatients rates offers a good approximation of the real population incidence.

#### Weather data

Intra-daily data on temperature (ºC) and relative humidity (RH, %) were collected from Iowa Environmental Mesonet [55], from October 2015 to January 2023, for all stations available in each region.

Absolute humidity (AH, *gr/m*^3^) was estimated following Eq (1):

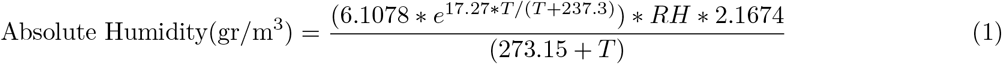

where T is temperature in Celsius and RH is relative humidity in %.

To extrapolate from station data to country data, daily values per station were first averaged to obtain the weekly average (of daily averages). These were calculated first at the state or province level (for the USA and Canada, respectively) and these weekly averages per state/province were weighted according to population to give the whole country average (S2 Fig, S3 Fig) to account for the weather conditions most people were exposed to.

#### Mobility data

Mobility data was first collected from the Bureau of Transportation Statistics (US Department of Transportation, [56]) from January 2019 to March 2022 for the USA. It includes estimations of the “Number of residents staying at home” (i.e., persons who make no trips with a trip end more than one mile away from home), the “Number of trips made by residents” (i.e., movements that include a stay of longer than 10 minutes at an anonymized location away from home) and the number of trips by distance (including number of trips shorter than one mile, between one and three miles, between three and five, etc). These travel statistics are produced from an anonymized national panel of mobile device data from multiple sources and a weighting procedure expands the sample, so that the results are representative of the entire population. From this data set, the variables “Number of residents staying at home” and the “Number of trips made by residents” were chosen for the analysis and are referred to by “Population at home” and “Number of trips”, respectively (S2 Fig).

To be able to analyze the effect of mobility in Canada, unavailable from the US Department of Transportation, data was further extracted from Google’s COVID-19 Community Mobility Reports (GCMR, [57]) available from February 2020 to October 2022 for both countries. This data set shows how visitors to (or time spent in) categorized places changed compared to a certain baseline day (the median value from the 5-week period between January 3 and February 6, 2020), for an aggregate of Android phone users. The “Residential” category shows a change in time spent in residential areas, whereas the other categories (“Recreation”, “Grocery and Pharmacy”, “Parks”, “Transit stations” and “Workplaces”) measure a change in total visitors to these places. From this data set, the variables “Residential” and the “Transit stations” were chosen for the analysis, as these are similar to the Bureau of Transportation Statistics data set (S2 Fig), and are referred to by “Residential time” and “Transit stations visitors”, respectively (S3 Fig).

### Data analysis

#### Clustering

Clustering was used to identify groups of viruses that have more similar temporal dynamics. Virus-specific weekly incidence time series were normalized to the maximum of each season (starting in the last week of August), therefore considering only the variability due to timing and patterns of the waves and not their amplitude. These incidence time series were clustered using hierarchical clustering. The pairwise Euclidean distance between time series was computed and Ward’s linkage method (an agglomerative algorithm) was used to construct the dendrograms. Clustering was performed in Python3, using scipy.cluster.hierarchy.dendrogram.

#### Correlations

Correlations were used to further explore the similarities between the temporal dynamics of the viruses, as well as with the weather variables. Pairwise Pearson correlations were calculated between the time series of the virus-specific weekly incidence, mean temperature, RH, and AH, after normalization to the maximum of each season, as previously described. Correlation matrices are depicted using heatmaps with the viruses sorted by correlation to IVA. The correlations and heatmaps were performed in Python3 using pandas.corr and seaborn.heatmap, respectively.

#### Regressions

Regression models were used to investigate the associations between the incidence of the different viruses, weather conditions, and mobility.

First, incidence was transformed (dividing by 1000) to represent the proportion of people infected in the population. The effects of weather and mobility on incidence were then analyzed using beta regression, appropriate for situations where the variable of interest is continuous and restricted to the interval (0, 1) [58].

To avoid issues with values equal to zero, 2×10^*−*8^ was added to the whole incidence time series. Analysis was performed using the statsmodels.betareg in Python3.

Second, Pearson correlation and Principal Component Analysis (PCA) were used to pre-select the independent weather variables. As temperature and AH are strongly correlated and two clusters appeared (one with T and AH, and the other with RH, S4 Fig), the weather analysis models included the following 5 variable combinations: temperature, AH, temperature-RH, AH-RH and no weather variable.

Third, incidence data was tested for auto-correlation, which was expected for two reasons: a) because it is inherent to any transmission process, meaning that the number of infections at a certain time point has an effect on the number of infections in subsequent time points; and b) because ILI-cases are seasonal, with a period of approximately one year. To account for transmission auto-correlation, the incidence in the previous week or the incidence in the two previous weeks (Y_*t−*1_, Y_*t−*2_) were included as independent variables, and are referred to by auto-correlation (AC) terms. Therefore, in the weather analysis 15 models were tested: the 5 weather combinations with the three AC possibilities (1, 2 or no previous week incidence). In the mobility-weather analysis, the same 5 weather combinations with and without the mobility variable were tested (10 models in total), all including the Y_*t−*1_ AC term. Seasonal auto-correlation was evaluated in the residuals of the models, using the Breusch–Godfrey test (.acorr breusch godfrey, in statsmodels) at lag 53 (weeks) to include the effects up to 1 year (S1 Table to S6 Table).

Finally, the different models for each virus and period were compared using both R^2^ and Akaike Information Criteria (AIC) and considered to be identical if the difference in AIC (ΔAIC) was *<*= 3, and similar when the difference in AIC was *<*= 10. For the pseudo-R^2^, rsquared (Python3 statsmodels) was used to compute the standard Cox-Snell version based on the likelihood ratio. In the paper, for the weather-only analysis the temperature-RH-AC model was chosen to compare the results because, first, there is previous evidence of the relevance of a combination of these two factors on the epidemiological dynamics of some respiratory virus, and, second, the analysis results showed that this model represented an adequate model for comparison of all viruses, regions and periods (see Results). For the models that also include mobility, the temperature-AC, mobility-AC and temperature-mobility-AC were chosen for comparison for the same reasons. In both analysis, the model with the AC term alone was used as the null model for these comparisons. The results of all models can be found in the supplementary tables (S1 Table to S6 Table).

## Results

### Weather effect pre-COVID-19 pandemic

Different respiratory viruses co-occur during the common flu season, but their incidence and dynamics are not exactly the same. Fig. 1 shows the incidence for IVA, IVB, RSV, hCoVs and hMPV, for the USA and Canada, from 2015-2016 to January 2023.

To analyze the effects of weather, we performed three different analysis: first, we looked at the different viruses and identified the ones that tend to co-circulate; second, we asked whether the different groups correlate differently with the different weather variables; and third, we used statistical modelling to infer the effect of weather on the observed dynamics. To identify viruses that have similar circulation patterns, we performed hierarchical clustering on the standardized incidences of all viruses, by country. This standardization removes all variation due to the amplitude, focusing the analysis on timing and shape. Two groups of viruses emerge from this analysis for both countries, one including IVA, RSV and hCoVs; and the other including IVB and hMPV (Fig. 2A, Fig. 2D).

**Fig 2.**
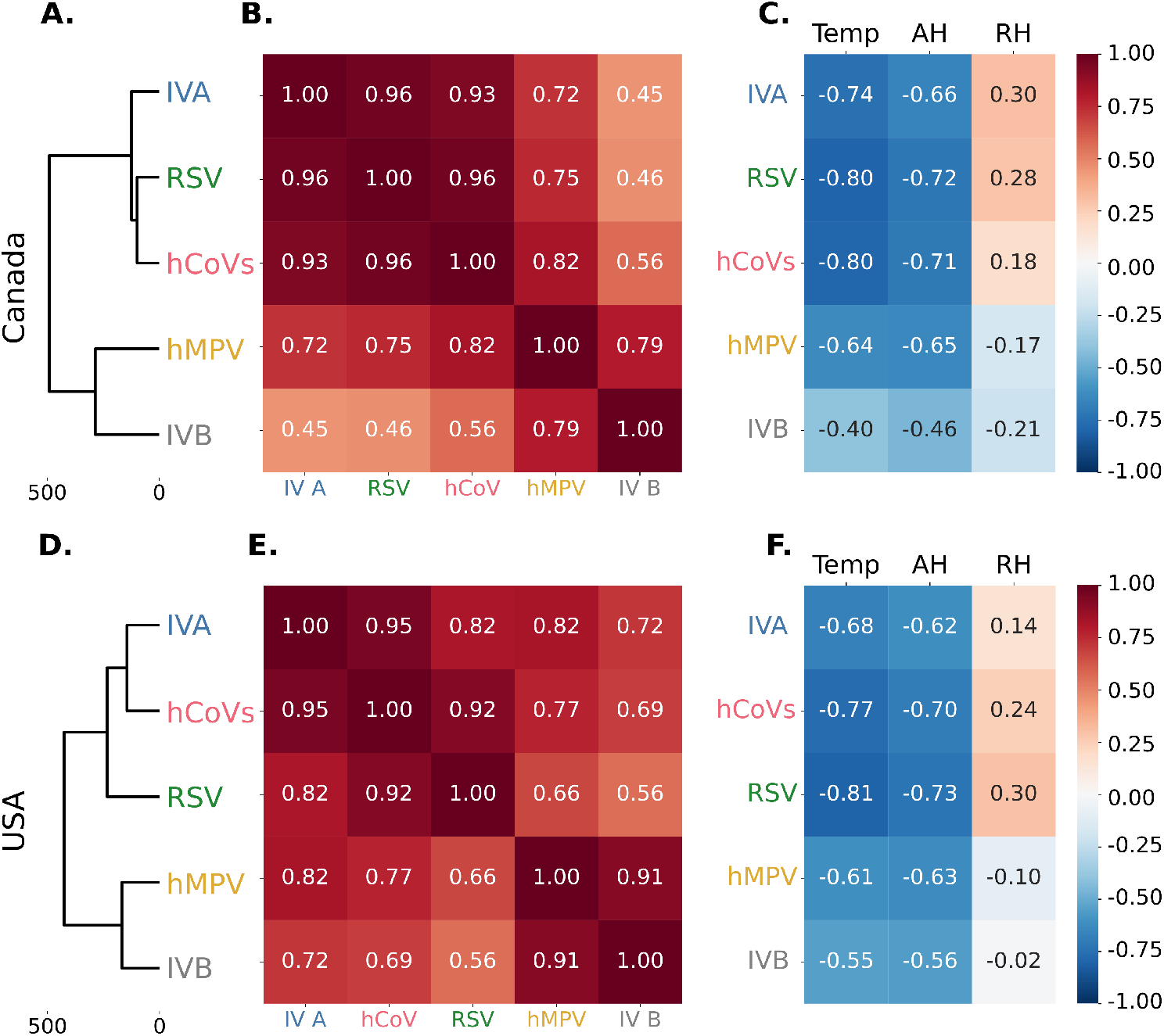
Hierarchical cluster analysis and correlation patterns between different viruses and weather in the pre-COVID-19 pandemic period in Canada and the USA. (A) and (D) Hierarchical clustering dendograms for the different viruses in Canada and the USA, respectively. (B) and (E) Correlations between the incidence of the viruses in Canada and the USA, respectively. (C) and (F) Correlation between the incidence of the viruses and temperature, AH and RH in Canada and the USA, respectively.

We then performed pair-wise correlation analysis of the standardized incidence by country, for all viruses and with the different weather variables. There is a general positive correlation between all viruses, but the two groups identified previously stand out consistently in both countries (Fig. 2B, Fig. 2E). As others before us, we found strong correlations between the viruses, temperature and AH, but weaker or no correlation with RH. Moreover, the two groups of viruses identified earlier by their dynamics reflect different patterns of correlation with the weather variables: the first shows stronger negative correlations with temperature and AH, and weak positive correlations with RH (Fig. 2C, Fig. 2F), whereas the second shows weaker negative correlations with temperature and AH, and weak negative or no correlation with RH (Fig. 2C, Fig. 2F).

To further analyse the weight of the weather variables in the incidence, we performed a beta regression. All models and respective parameters are summarized in the supplementary tables. For comparison purposes we decided to show the model that includes auto-correlation (AC), temperature, and relative humidity (RH), as it was systematically either among the best models and/or had very similar dynamics and regression coefficients to the best models, see S1 Table, S2 Table).

This analysis showed that, for every virus, in both countries, including weather variables considerably improves the models (the AIC difference between model with and without weather variables is larger than 10 units,ΔAIC*>*10) (Table 1, Table 2). Indeed, all best models (AIC difference from the absolute best model ΔAIC*<*=3) included at least one weather variable (plus the AC term) (S1 Table). Furthermore, this analysis confirmed the differential effect of weather, revealing a stronger dependency on temperature (or AH) for the first group of viruses compared with the second (Fig. 3A and 3C, Table 1 and 2). Regarding RH, its effect is much weaker but still significant for many of these viruses (Fig. 3A and 3C, Tables 1 and 2). Indeed, this model is able to explain a considerable proportion of the variation in the incidence of these viruses (ranging from 64% up to 91%, Tables 1 and 2). These results are consistent even when considering only two seasons (S5 Fig).

**Table 1.** Regression coefficients (with p-value between brackets), AIC and R^2^ for the models that include: (1) auto-correlation (AC), (2) temperature and AC, and (3) temperature, RH, and AC for Canada pre-COVID-19 pandemic.

| | Model | Temperature | RH | AC | AIC | $R^2$ |
| --- | --- | --- | --- | --- | --- | --- |
| IVA | 1 | -- | -- | 0.67 (0.0) | -2243.0 | 0.59 |
|  | 2 | -0.77 (0.0) | -- | 0.37 (0.0) | -2382.0 | 0.81 |
|  | 3 | -0.77 (0.0) | 0.06 (0.17) | 0.37 (0.0) | -2382.0 | 0.81 |
| RSV | 1 | -- | -- | 0.61 (0.0) | -2454.0 | 0.58 |
|  | 2 | -0.78 (0.0) | -- | 0.35 (0.0) | -2609.0 | 0.82 |
|  | 3 | -0.78 (0.0) | 0.06 (0.2) | 0.35 (0.0) | -2609.0 | 0.82 |
| hCoVs | 1 | -- | -- | 0.63 (0.0) | -2618.0 | 0.59 |
|  | 2 | -0.72 (0.0) | -- | 0.39 (0.0) | -2742.0 | 0.79 |
|  | 3 | -0.74 (0.0) | -0.11 (0.01) | 0.39 (0.0) | -2746.0 | 0.8 |
| IVB | 1 | -- | -- | 0.7 (0.0) | -2752.0 | 0.54 |
|  | 2 | -0.42 (0.0) | -- | 0.6 (0.0) | -2789.0 | 0.63 |
|  | 3 | -0.46 (0.0) | -0.16 (0.01) | 0.59 (0.0) | -2793.0 | 0.64 |
| hMPV | 1 | -- | -- | 0.62 (0.0) | -2711.0 | 0.64 |
|  | 2 | -0.37 (0.0) | -- | 0.49 (0.0) | -2756.0 | 0.72 |
|  | 3 | -0.46 (0.0) | -0.25 (0.0) | 0.46 (0.0) | -2787.0 | 0.77 |

**Table 2.** Regression coefficients (with p-value between brackets), AIC and R^2^ for the models that include: (1) auto-correlation (AC), (2) temperature and AC, and (3) temperature, RH, and AC for the USA pre-COVID-19 pandemic.

| | Model | Temperature | RH | AC | AIC | $R^2$ |
| --- | --- | --- | --- | --- | --- | --- |
| IVA | 1 | -- | -- | 0.86 (0.0) | -2748.0 | 0.7 |
|  | 2 | -0.59 (0.0) | -- | 0.64 (0.0) | -2851.0 | 0.81 |
|  | 3 | -0.59 (0.0) | 0.03 (0.3) | 0.64 (0.0) | -2850.0 | 0.81 |
| RSV | 1 | -- | -- | 0.76 (0.0) | -2043.0 | 0.71 |
|  | 2 | -0.77 (0.0) | -- | 0.46 (0.0) | -2225.0 | 0.9 |
|  | 3 | -0.83 (0.0) | 0.12 (0.0) | 0.42 (0.0) | -2250.0 | 0.91 |
| hCoVs | 1 | -- | -- | 0.88 (0.0) | -3069.0 | 0.76 |
|  | 2 | -0.63 (0.0) | -- | 0.51 (0.0) | -3162.0 | 0.85 |
|  | 3 | -0.66 (0.0) | 0.13 (0.0) | 0.51 (0.0) | -3183.0 | 0.87 |
| IVB | 1 | -- | -- | 0.59 (0.0) | -2973.0 | 0.55 |
|  | 2 | -0.44 (0.0) | -- | 0.53 (0.0) | -3037.0 | 0.66 |
|  | 3 | -0.47 (0.0) | -0.21 (0.0) | 0.56 (0.0) | -3060.0 | 0.7 |
| hMPV | 1 | -- | -- | 0.77 (0.0) | -2311.0 | 0.76 |
|  | 2 | -0.49 (0.0) | -- | 0.61 (0.0) | -2404.0 | 0.86 |
|  | 3 | -0.49 (0.0) | -0.06 (0.02) | 0.61 (0.0) | -2407.0 | 0.86 |

**Fig 3.**
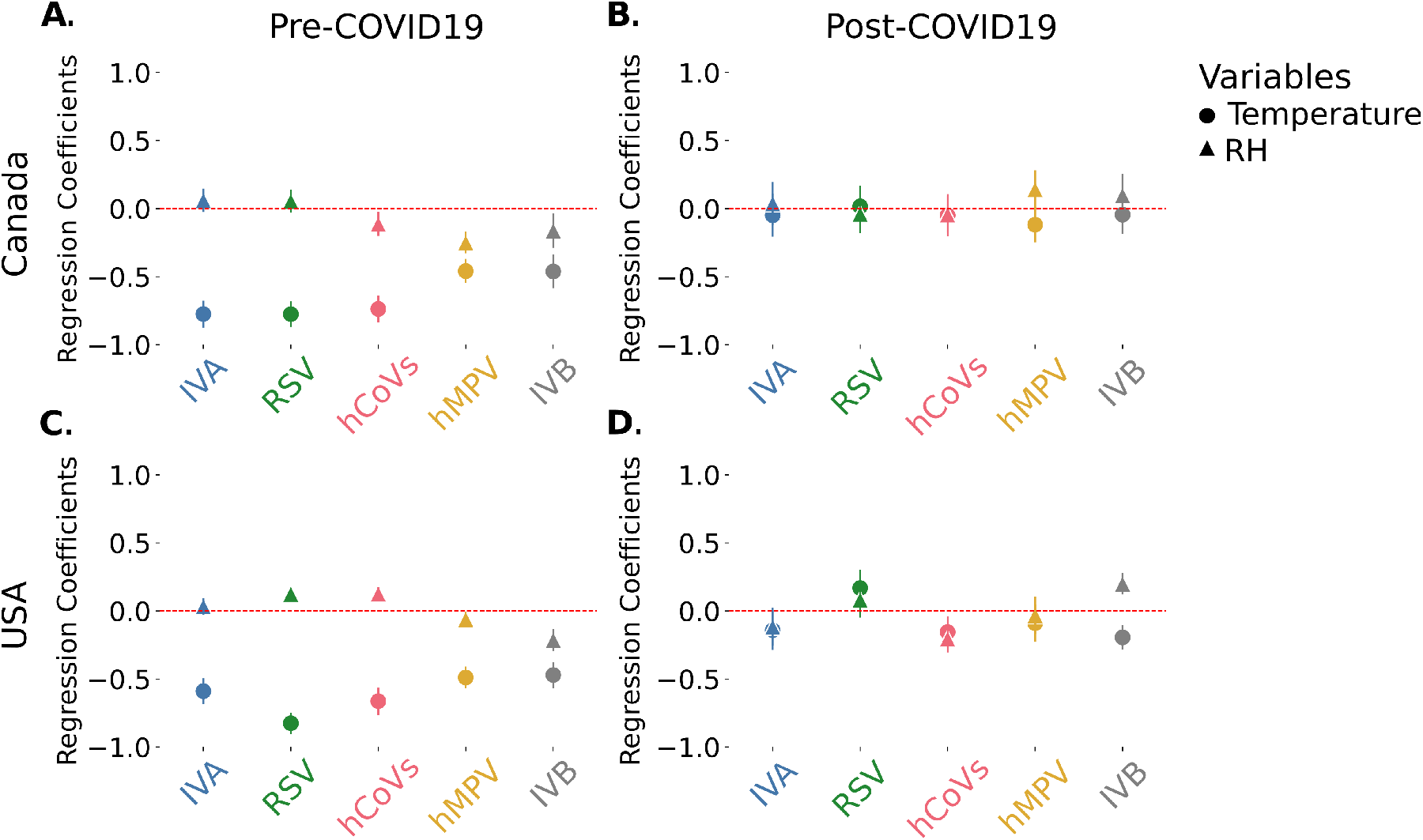
Regression coefficients for the temperature-RH model in the pre-COVID-19 and post-COVID-19 pandemic period in Canada and the USA. Temperature and RH regression coefficients with the 95% confidence intervals for the incidence of all the viruses analyzed for (A) Canada pre-COVID-19 pandemic, (B) Canada post-COVID-19 pandemic, (C) USA pre-COVID-19 pandemic and (D) USA post-COVID-19 pandemic. Circles: temperature coefficients; Triangles: RH coefficients. Coefficients for the AC term are not represented.

### Epidemiological dynamics post-COVID-19

All viruses in the data set showed a pronounced decline in incidence after March 2020 (Fig. 1C-F) and remained very low until the spring-summer 2021. In the spring-summer 2021, we observed a slight increase in hCoVs activity, specially in the USA, and increased RSV activity starting in the summer 2021 until February 2022, in the USA, and just a few weeks later, in Canada. In the winter 21-22 we also observed off-season surges in IVA and hMPV in the USA, as well as in hCoVs in both countries. During the spring-summer 2022 all viruses in both countries showed increased activity or off-season surges. Similarly, an early winter epidemic during the the autumn-winter 2022-2023 was detected for all the viruses in both countries, being less intense for RSV and hMPV in Canada. Finally, IVB showed extremely low levels in both countries from March 2020 (Fig. 1G) and, therefore, we excluded it from the post-COVID-19 pandemic analysis.

### Weather effect post-COVID-19 pandemic

As the observed dynamics had a less obvious seasonal pattern, we repeated the previous analysis, on the incidence of the different viruses and their relation with weather, in the period after the COVID-19 pandemic was declared (March 2020) until January 2023 (Fig. 1).

As expected from observing the incidence time series, and reflecting the disruption in dynamics observed in the incidence patterns for this period in Canada and the USA (Fig. 1), the hierarchical clustering no longer revealed two clear groups and no clustering pattern is shared between Canada and the USA (Fig. 4A and 4D). The results of the correlation analysis were consistent with the clustering results, showing much weaker correlations among the different viruses and revealing no clear groups (Fig. 4B and 4E), after the pandemic started. Regarding the correlations between the viruses and weather in the same period, we observed again a much weaker correlations and no clear patterns (Fig. 4C and 4F).

**Fig 4.**
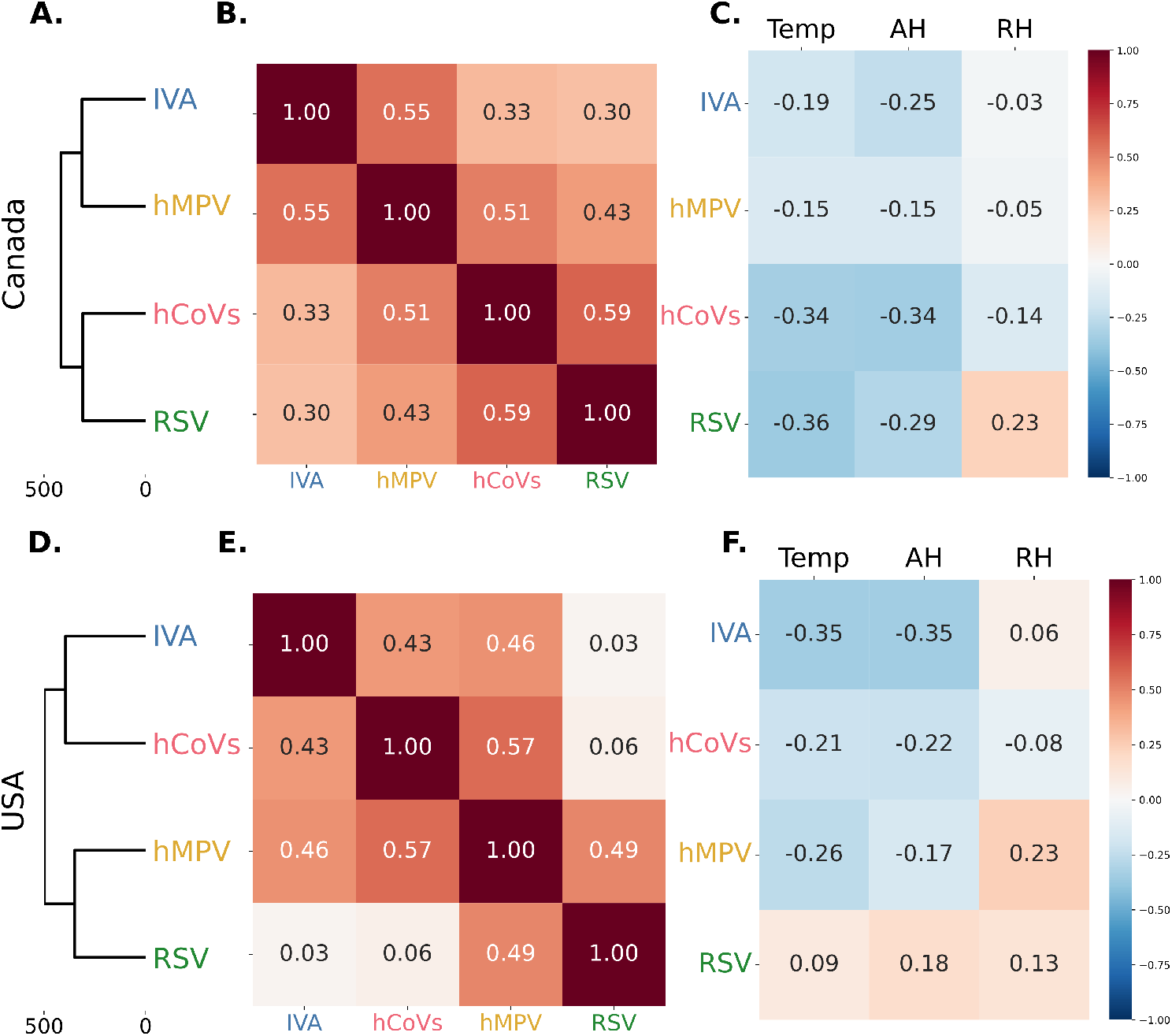
Hierarchical cluster analysis and correlation patterns between different viruses and weather in the post-COVID-19 pandemic period in Canada and the USA. (A) and (D) hierarchical clustering dendograms for the different viruses in Canada and the USA, respectively. Note that the distances between the viruses in the x-axis are larger than in the pre-COVD19 pandemic period (Fig. 2). (B) and (E) Correlations between the incidence of the viruses in Canada and the USA, respectively. (C) and (F) Correlation between the incidence of the viruses and temperature, AH and RH in Canada and the USA, respectively.

We then performed the same regression analysis for the post-COVID-19 period. Adding a weather variable only improved the models for hMPV in Canada and, RSV and hCoVs in the USA. Moreover, it was only is the later case that the models with and without weather were considerably different (ΔAIC*>*10) (Table 3 and 4, S2 Table). To facilitate comparison, we again choose the model including temperature and RH (plus the AC term) as, again it was either among the best models or the regression coefficients were very similar and followed similar patterns, see S2 Table. This regression analysis confirmed a much weaker, and often non-significant, effect of temperature (or AH), in the pandemic period, for all viruses and in both countries (Figures 3B, 3D and Tables 3 and 4). In fact, the models can only explain a limited proportion of the variation in viral dynamics (ranging from 40% to 65%, Tables 3 and 4), in this period. For every virus in both countries, all best models included the AC variable (S2 Table).

**Table 3.** Regression coefficients (with p-value between brackets), AIC and R^2^ for the models that include: (1) auto-correlation (AC), (2) temperature and AC, and (3) temperature, RH, and AC for Canada post-COVID-19 pandemic.

| | Model | Temperature | RH | AC | AIC | $R^2$ |
| --- | --- | --- | --- | --- | --- | --- |
| IVA | 1 | -- | -- | 0.7 (0.0) | -2991.0 | 0.4 |
|  | 2 | -0.06 (0.49) | -- | 0.69 (0.0) | -2990.0 | 0.41 |
|  | 3 | -0.05 (0.54) | 0.04 (0.63) | 0.68 (0.0) | -2988.0 | 0.41 |
| RSV | 1 | -- | -- | 0.84 (0.0) | -2467.0 | 0.61 |
|  | 2 | 0.02 (0.76) | -- | 0.84 (0.0) | -2465.0 | 0.61 |
|  | 3 | 0.02 (0.83) | -0.04 (0.59) | 0.86 (0.0) | -2463.0 | 0.61 |
| hCoVs | 1 | -- | -- | 0.69 (0.0) | -2484.0 | 0.55 |
|  | 2 | -0.04 (0.62) | -- | 0.68 (0.0) | -2482.0 | 0.55 |
|  | 3 | -0.05 (0.53) | -0.04 (0.53) | 0.68 (0.0) | -2481.0 | 0.55 |
| hMPV | 1 | -- | -- | 0.68 (0.0) | -2656.0 | 0.52 |
|  | 2 | -0.15 (0.02) | -- | 0.7 (0.0) | -2660.0 | 0.54 |
|  | 3 | -0.12 (0.07) | 0.14 (0.04) | 0.73 (0.0) | -2662.0 | 0.55 |

**Table 4.** Regression coefficients (with p-value between brackets), AIC and R^2^ for the models that include: (1) auto-correlation (AC), (2) temperature and AC, and (3) temperature, RH, and AC for the USA post-COVID-19 pandemic.

| | Model | Temperature | RH | AC | AIC | $R^2$ |
| --- | --- | --- | --- | --- | --- | --- |
| IVA | 1 | -- | -- | 0.71 (0.0) | -2177.0 | 0.49 |
|  | 2 | -0.14 (0.06) | -- | 0.68 (0.0) | -2179.0 | 0.51 |
|  | 3 | -0.14 (0.06) | -0.12 (0.1) | 0.72 (0.0) | -2179.0 | 0.51 |
| RSV | 1 | -- | -- | 0.65 (0.0) | -1952.0 | 0.56 |
|  | 2 | 0.16 (0.02) | -- | 0.68 (0.0) | -1956.0 | 0.57 |
|  | 3 | 0.17 (0.01) | 0.08 (0.22) | 0.68 (0.0) | -1955.0 | 0.58 |
| hCoVs | 1 | -- | -- | 0.6 (0.0) | -2153.0 | 0.6 |
|  | 2 | -0.12 (0.04) | -- | 0.58 (0.0) | -2155.0 | 0.62 |
|  | 3 | -0.16 (0.01) | -0.2 (0.0) | 0.64 (0.0) | -2167.0 | 0.65 |
| hMPV | 1 | -- | -- | 0.62 (0.0) | -2203.0 | 0.52 |
|  | 2 | -0.09 (0.22) | -- | 0.61 (0.0) | -2202.0 | 0.53 |
|  | 3 | -0.09 (0.2) | -0.03 (0.65) | 0.62 (0.0) | -2200.0 | 0.53 |

### Mobility effect pre- and post-COVID-19 pandemic

As mentioned before, another clear driver of epidemics is human behavior. However, as weather models were so good at explaining seasonality of IVs and NIRVs, prior to the COVID-19 pandemic, behaviour has somewhat received less attention. That we observe not only a disruption in incidence but also in seasonality, post-pandemic, and that these viruses are clearly able to spread during warmer months in temperate countries, raises important questions regarding the relative role of weather in their spread. Thus, we next tested whether mobility patterns, as a relevant component of behaviour, could help better understand the dynamics of the viruses in the period after the advent of COVID-19.

We first used the “number of trips” and “population at home” reported by the US Department of Transportation, which allowed us to compare the effects of mobility before and after the COVID-19 pandemic in the USA. Correlation analysis show that in the pre-COVID-19 pandemic period available for this data set (January 2019-March 2020), the number of trips showed a negative correlation with the incidence of the viruses, whereas the population at home showed positive or no correlation S2 Fig. This is possibly the result of the strong positive correlation in that single season of the number of trips and warm weather S2 Fig. In the post-COVID-pandemic period, although generally weaker, the number of trips showed a positive correlation with the incidence of the viruses, whereas the population at home showed a negative correlation S2 Fig.

Regarding the regression analysis, we compared the models with weather (plus the AC term) to models that also incorporated different mobility measures. For the pre-COVID-19 pandemic period with available mobility data (January 2019-March 2020), including the number of trips did not improve the models or the proportion of variation explained (Table 5, S3 Table). For RSV and hCoVs, the difference in AIC between the best without and with mobility models is just over 3, meaning that the model with mobility is only slightly better than the one without it (S3 Table). As seen previously for longer pre-COVID-19 pandemic periods, the effect of temperature in this single season is, in general, large and negative (Fig. 5A). Similarly, including the population at home did not improve the models or the proportion of variation explained, although, again, for RSV, the model with mobility is slightly better than the one without (S6 Fig, S4 Table).

**Table 5.** Regression coefficients (with p-value between brackets), AIC and R^2^ for the models that include: (1) auto-correlation (AC), (2) temperature and AC, (3)number of trips and (4) AC and temperature, number of trips and AC term for USA pre-COVID-19 pandemic.

| | Model | Temperature | No. Trips | AC | AIC | $R^2$ |
| --- | --- | --- | --- | --- | --- | --- |
| IVA | 1 | -- | -- | 1.02 (0.0) | -688.0 | 0.79 |
|  | 2 | -0.77 (0.0) | -- | 0.69 (0.0) | -727.1 | 0.89 |
|  | 3 | -- | -0.5 (0.0) | 0.72 (0.0) | -702.7 | 0.84 |
|  | 4 | -0.77 (0.0) | 0.0 (0.97) | 0.7 (0.0) | -725.1 | 0.89 |
| RSV | 1 | -- | -- | 0.6147 (0.0) | -436.4 | 0.74 |
|  | 2 | -0.51 (0.0) | -- | 0.47 (0.0) | -472.9 | 0.9 |
|  | 3 | -- | -0.31 (0.0) | 0.45 (0.0) | -443.6 | 0.79 |
|  | 4 | -0.62 (0.0) | 0.16 (0.05) | 0.52 (0.0) | -474.4 | 0.9 |
| hCoVs | 1 | -- | -- | 1.04 (0.0) | -800.2 | 0.83 |
|  | 2 | -0.7 (0.0) | -- | 0.65 (0.0) | -837.6 | 0.91 |
|  | 3 | -- | -0.35 (0.02) | 0.75 (0.0) | -803.6 | 0.85 |
|  | 4 | -0.82 (0.0) | 0.21 (0.06) | 0.75 (0.0) | -838.7 | 0.92 |
| hMPV | 1 | -- | -- | 0.55 (0.0) | -546.9 | 0.85 |
|  | 2 | -0.14 (0.0) | -- | 0.5 (0.0) | -553.6 | 0.88 |
|  | 3 | -- | -0.09 (0.04) | 0.5 (0.0) | -548.8 | 0.87 |
|  | 4 | -0.16 (0.02) | 0.03 (0.64) | 0.5 (0.0) | -551.8 | 0.88 |

Note that the differences between the absolute best with and without mobility (number of trips) models for RSV and hCoVs in the pre-COVID-19 pandemic period are 3.7 and 3.3 units, respectively (S4 Table)

**Fig 5.**
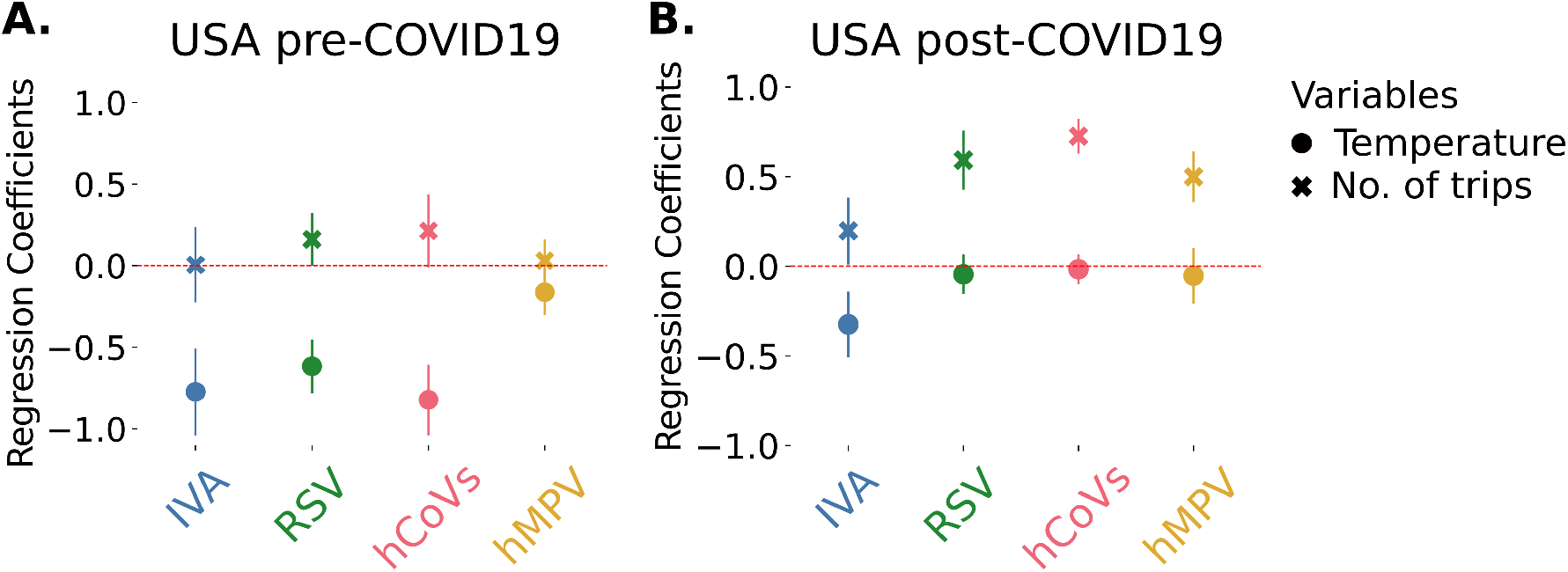
Effects of number of trips during the pre and post-COVID-19 pandemic period in the USA. (A) temperature and number of trips regression coefficients for the incidence of all the viruses with the 95% confidence intervals in the pre-COVID-19 pandemic period. (B) temperature and number of trips regression coefficients for the incidence of all the viruses with the 95% confidence intervals in the post-COVID-19 pandemic period.

However, in the post-COVID-19 pandemic period available for this mobility data set (March 2020-March 2022), including the total number of trips improved the models for all viruses and increased the variation explained by the models (Table 6, S3 Table), the only exception being IVA (where all models are similar, i.e., all model’s AIC are within 10 units, S3 Table). Indeed, the mobility alone model (plus the AC term) is, for every virus, among the best models(S3 Table). In these models, there is a significant positive effect of mobility in most cases, so that the less travel the less incidence we find (Fig. 5B, Table 6). The exceptions are IVA where the effect is borderline significant (p-value=0.04). The effect of mobility is larger in magnitude than the effect of temperature in most models (except for IVA). Similarly, including the population at home improved the models and the proportion of variation explained, except again for IVA (where all models are similar, i.e., all model’s AIC are within 10 units, S6 Fig, S4 Table). In this period, there is a significant negative effect of population at home in the incidence of most of this viruses, so that the less people at home the more incidence is found, with the exceptions again of IVA (S6 Fig, S4 Table).

**Table 6.** Regression coefficients (with p-value between brackets), AIC and R^2^ for the models that include: (1) auto-correlation (AC), (2) temperature and AC, (3) number of trips and AC, and (4)temperature, number of trips and AC for USA post-COVID-19 pandemic.

| | Model | Temperature | No. Trips | AC | AIC | $R^2$ |
| --- | --- | --- | --- | --- | --- | --- |
| IVA | 1 | -- | -- | 0.57 (0.0) | -1771.0 | 0.42 |
|  | 2 | -0.27 (0.0) | -- | 0.55 (0.0) | -1778.0 | 0.47 |
|  | 3 | -- | 0.12 (0.21) | 0.57 (0.0) | -1770.0 | 0.43 |
|  | 4 | -0.32 (0.0) | 0.2 (0.04) | 0.55 (0.0) | -1780.0 | 0.49 |
| RSV | 1 | -- | -- | 1.07 (0.0) | -1578.0 | 0.77 |
|  | 2 | -0.11 (0.07) | -- | 1.12 (0.0) | -1579.0 | 0.78 |
|  | 3 | -- | 0.61 (0.0) | 0.93 (0.0) | -1628.0 | 0.86 |
|  | 4 | -0.04 (0.43) | 0.59 (0.0) | 0.96 (0.0) | -1626.0 | 0.86 |
| hCoVs | 1 | -- | -- | 0.51 (0.0) | -1558.0 | 0.52 |
|  | 2 | -0.1 (0.2) | -- | 0.5 (0.0) | -1558.0 | 0.52 |
|  | 3 | -- | 0.73 (0.0) | 0.67 (0.0) | -1687.0 | 0.86 |
|  | 4 | -0.02 (0.7) | 0.72 (0.0) | 0.67 (0.0) | -1685.0 | 0.86 |
| hMPV | 1 | -- | -- | 0.72 (0.0) | -1678.0 | 0.53 |
|  | 2 | -0.13 (0.14) | -- | 0.71 (0.0) | -1678.0 | 0.54 |
|  | 3 | -- | 0.51 (0.0) | 0.88 (0.0) | -1719.0 | 0.68 |
|  | 4 | -0.05 (0.51) | 0.5 (0.0) | 0.86 (0.0) | -1717.0 | 0.68 |

We then used Google Mobility reports data, in particular the “Residential time” and the “Transit stations” categories, which allowed us to compare the effect of mobility in the available post-COVID-19 pandemic period (March 2020-October 2022) for Canada and the USA. Correlations between the incidences of the viruses and these mobility variables are in general weak, but a positive correlation was observed for visitors to transit stations whereas a negative correlation was observed for residential time (S2 Fig), S3 Fig). Residential time and population at home (from the previous mobility data set) had a strong, positive correlation, similar to the correlation between visitors to transit stations and number of trips (from the previous mobility data set) (S2 Fig).

Regarding the regression analysis, including the visitors at transit stations variable improved the models for all viruses, except for IVA in the USA (where all models are similar, i.e., all model’s AIC are within 10 units, Table 7, S5 Table), increasing slightly the proportion of variation explained by the models (Table 7, S5 Table). In these models, there is a significant positive effect of the mobility variable in both countries, so that the less visitors in transit stations the less incidence is found (Fig. 6A and 6B). This effect is similar or larger in magnitude to the effect of temperature in all models. Similarly, including the residential time variable improved the models and the proportion of variation explained, except again for IVA in the USA (where all models are similar, i.e., all model’s AIC are within 10 units)(S7 Fig, S6 Table). In this period, there is a significant negative effect of residential time in the incidence of most of this viruses, so that the less time people spend in residential areas the more incidence is found (S7 Fig, S6 Table).

**Table 7.** Regression coefficients (with p-value between brackets), AIC and R^2^ for the models that include: (1) auto-correlation (AC), (1)temperature and AC, (3)visitors at transit stations and AC, and (4) temperature, visitors at transit stations and AC term for Canada post-COVID-19 pandemic.

| | Model | Temperature | Transit Stations | AC | AIC | $R^2$ |
| --- | --- | --- | --- | --- | --- | --- |
| IVA | 1 | -- | -- | 0.77 (0.0) | -2924.0 | 0.43 |
|  | 2 | -0.02 (0.81) | -- | 0.77 (0.0) | -2922.0 | 0.43 |
|  | 3 | -- | 0.31 (0.0) | 0.68 (0.0) | -2936.0 | 0.49 |
|  | 4 | -0.11 (0.15) | 0.34 (0.0) | 0.65 (0.0) | -2936.0 | 0.5 |
| RSV | 1 | -- | -- | 0.74 (0.0) | -2338.0 | 0.56 |
|  | 2 | 0.14 (0.07) | -- | 0.79 (0.0) | -2340.0 | 0.57 |
|  | 3 | -- | 0.36 (0.0) | 0.77 (0.0) | -2367.0 | 0.65 |
|  | 4 | -0.08 (0.29) | 0.38 (0.0) | 0.74 (0.0) | -2366.0 | 0.65 |
| hCoV | 1 | -- | -- | 0.58 (0.0) | -2311.0 | 0.47 |
|  | 2 | -0.02 (0.77) | -- | 0.58 (0.0) | -2309.0 | 0.47 |
|  | 3 | -- | 0.37 (0.0) | 0.66 (0.0) | -2330.0 | 0.54 |
|  | 4 | -0.36 (0.0) | 0.53 (0.0) | 0.56 (0.0) | -2345.0 | 0.6 |
| hMPV | 1 | -- | -- | 0.64 (0.0) | -2475.0 | 0.46 |
|  | 2 | -0.08 (0.25) | -- | 0.65 (0.0) | -2475.0 | 0.47 |
|  | 3 | -- | 0.43 (0.0) | 0.49 (0.0) | -2492.0 | 0.53 |
|  | 4 | -0.22 (0.0) | 0.49 (0.0) | 0.47 (0.0) | -2500.0 | 0.57 |

**Table 8.** Regression coefficients (with p-value between brackets), AIC and R^2^ for the models that include: (1) auto-correlation (AC), (2) temperature and AC, (3) visitors at transit stations and AC, and (4) temperature, visitors at transit stations and AC term for USA post-COVID-19 pandemic.

| | Model | Temperature | Transit Stations | AC | AIC | $R^2$ |
| --- | --- | --- | --- | --- | --- | --- |
| IVA | 1 | - - | - - | 0.49 (0.0) | -2118.0 | 0.37 |
|  | 2 | -0.11 (0.14) | - - | 0.48 (0.0) | -2118.0 | 0.38 |
|  | 3 | - - | 0.05 (0.44) | 0.47 (0.0) | -2117.0 | 0.38 |
|  | 4 | -0.24 (0.02) | 0.19 (0.04) | 0.42 (0.0) | -2120.0 | 0.4 |
| RSV | 1 | - - | - - | 0.94 (0.0) | -1949.0 | 0.75 |
|  | 2 | 0.01 (0.81) | - - | 0.93 (0.0) | -1947.0 | 0.75 |
|  | 3 | - - | 0.36 (0.0) | 0.81 (0.0) | -1977.0 | 0.8 |
|  | 4 | -0.05 (0.33) | 0.36 (0.0) | 0.81 (0.0) | -1976.0 | 0.8 |
| hCoV | 1 | - - | - - | 0.53 (0.0) | -1983.0 | 0.55 |
|  | 2 | -0.06 (0.32) | - - | 0.52 (0.0) | -1982.0 | 0.56 |
|  | 3 | - - | 0.26 (0.0) | 0.58 (0.0) | -2015.0 | 0.66 |
|  | 4 | -0.22 (0.0) | 0.3 (0.0) | 0.54 (0.0) | -2026.0 | 0.69 |
| hMPV | 1 | - - | - - | 0.63 (0.0) | -2054.0 | 0.51 |
|  | 2 | -0.01 (0.89) | - - | 0.63 (0.0) | -2052.0 | 0.51 |
|  | 3 | - - | 0.38 (0.0) | 0.74 (0.0) | -2097.0 | 0.65 |
|  | 4 | -0.23 (0.0) | 0.43 (0.0) | 0.71 (0.0) | -2104.0 | 0.67 |

**Fig 6.**
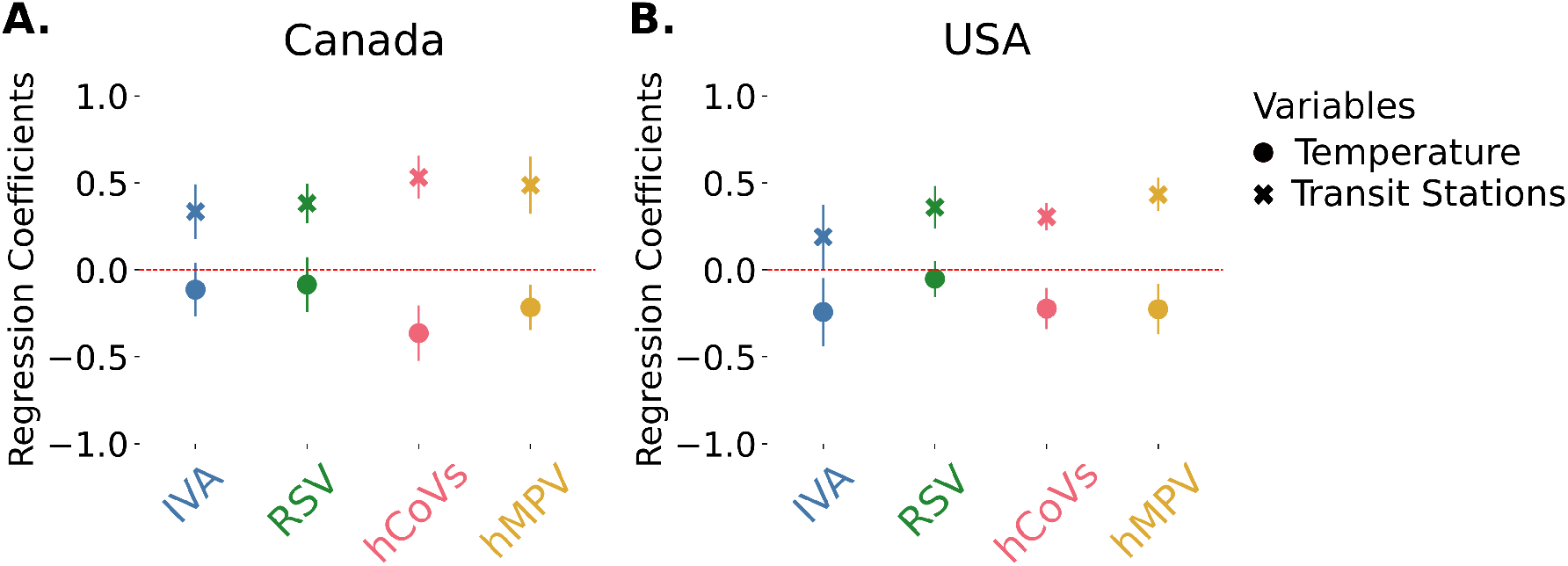
Effects of visits to transit stations (GMR) during the post-COVID-19 pandemic period in Canada and the USA. (A) temperature and visitors to transit stations coefficients for the incidence of all the viruses with the 95% confidence intervals in the post-COVID-19 pandemic period in Canada. (B) temperature and visitors in transit stations regression coefficients for the incidence of all the viruses with the 95% confidence intervals in the post-COVID-19 pandemic period in the USA (S5 Table)

In summary, if the role of mobility could be minimized before the COVID-19 pandemic, it becomes fundamental to understand respiratory viral dynamics after March 2020, and help explain “off-season” infections.

## Discussion

From the different factors that impact the seasonal dynamics of respiratory viruses, weather has received the most attention [35], and there is extensive data consistent with a strong weather effect on viral dynamics. However, and given the association between weather and human behavior (e.g. school start coinciding with the end of summer or the increase in indoor activities during the colder months), understanding the relative importance of each has proven difficult.

Here, we took advantage of the external shock provided by the first years of the COVID-19 pandemic and provide evidence indicating that in the pre-COVID-19 pandemic period weather had a significant effect on the epidemiological dynamics of all these viruses but that this effect is much less significant after the pandemic, at least in the USA and Canada. This disruption was observed in two different ways: first, in many temperate countries of the Northern Hemisphere, these non-SARS-CoV2 respiratory viruses virtually disappeared from March-April 2020 until at least March 2021; second, these viruses ressurged but with previously unobserved dynamics, peaking outside of their traditional seasons. Several explanations have been presented for both observations and we discuss them below.

The initial reduction in IVs and NIRVs in Canada and in the USA had been reported [59, 60] and, at least for IVs and RSV, non-pharmacological interventions (NPIs) are the most commonly mentioned factor to explain it [31, 61, 62]. In the case of Canada, a relation was established between this reduction and the early implementation of the initial lock-down, including stay-at-home orders, travel restrictions, school closures and mask mandates [59]. Another proposed explanation is viral interference between SARS-CoV-2 and other viruses [31, 61], which could have led to less infections. The case of IVB is remarkable, as it was suggested that the IVB/Yamagata lineage could have become partially extinct, due to the lack of animal reservoirs and other epidemiological characteristics [32, 63, 64]. Thus, and although the IVB/Victoria lineage has been detected globally after 2020, the surveillance data for USA and Canada presented here showed extremely low levels of IVB and was excluded from the analysis.

The data also shows that all viruses analyzed had “off-season” surges, with peaks in the spring-summer of 2021 and/or 2022, and/or unusually early seasonal peaks in the winter of 2022-2023. These atypical dynamics have been reported for RSV and IVs around the globe [31, 62, 65] but these changes are more difficult to explain: relaxation or lifting of NPIs [31, 66] could explain their ressurgence but not the new timing. One leading hypothesis is immunity waning in the population due to the reduction of viral circulation in 2020 and 2021, an effect also called “immunity debt” [67, 68]. One implicit implication of this hypothesis is that winter weather is neither a necessary nor sufficient condition for seasonal epidemics: it may facilitate infections but, if enough individuals are susceptible, summer waves could be sustained. If weather is less relevant to circulation than previously believed, some surges (in very young infants or immunocompromized groups) should be expected more frequently throughout the year, and some countries offer relevant exceptions that contradict at least the strong temperature dependency. For example, RSV is mostly a winter virus in the Northern Hemisphere but typically peaks in the summer and fall in Japan; even more interestingly, it had its highest surge on record during the spring of 2021, after having almost disappeared during the 2020 season [69] Another hypothesis is that mobility and mixing are particularly relevant drivers not only behind the probability of transmission, but in the seasonal patterns as well. Mobility has been shown to be a good indicator for the strength of lockdown measures and other NPIs [70, 71] and has been increasingly used in recent years as an indicator of behavioural changes affecting infection dynamics (e.g. [72–75]). Therefore, our results that indicate that mobility better explains the observed dynamics, are in line with previous evidence suggesting that the behavioral changes and the non-pharmacological interventions resulting from the COVID-19 pandemic had a significant effect on the epidemiology of these viruses.

This effect of shifting peaks had also been observed during the 2009 flu pandemic, after a new influenza strain was identified in April 2009 (influenza A (H1N1)pdm09). This strain reached pandemic status in June but, in most of the Northern Hemisphere, only peaked in the fall. In this case, waning immunity was unlikely, as it did not lead to general and long lock-downs, and viral interference was presented as the most likely explanation: the circulation of rhinoviruses might have delayed the H1N1 wave [76], which in turn might have delayed the RSV epidemic [77]. Still, the influenza peaks happened after schools resumed and the temperatures started to drop, in line with expected role of both mobility and weather in influenza transmission. Moreover, the subsequent epidemics, mostly driven by the pandemic strain, went back to their winter dynamics.

The COVID-19 pandemic also made clear how understudied some of the NIRVs are. There is extensive evidence from experimental and epidemiological studies on the effects of temperature or humidity on IVs and, despite being more limited, research on NIRVs dynamics also mostly focuses on weather variables [9, 78–80] but, to our knowledge, experimental evidence only exists for IVs. Even post-pandemic, and as far as we could gather, studies on NIRVs, other than RSV, have been limited to reports on the reductions in the circulation of these viruses immediately after the implementation of the interventions up to January 2022 [81–89].

Therefore, it is quite possible that the correlation with weather described for IVs, specially IVA, does not generalize so well to different NIRVs (and the temperature/humidity dependence of SARS-CoV2 remains an open debate). From our analysis, two different groups of viruses emerged in the pre-pandemic period, both in terms of their epidemiological patterns and their weather susceptibility, with IVA, RSV and hCoVs in one group, with higher apparent weather sensitivity; and IVB and hMPV in the other, with lower weather sensitivity. These results are in agreement with other pre-pandemic studies, on a smaller set of viruses, which showed that, in temperate regions, the start of the RSV and IVA epidemics were earlier and closer in time, followed by the start of IVB and hMPV epidemics [9] (with exceptions). The same study also found an association between temperature and RH, and the activity of IVA, IVB and RSV. Another study, using 7 years of data from patients in Edinburgh, Scotland, identified an effect of temperature on the activity of RSV, IVA, IVB and hMPV, being the largest in RSV and the lowest in hMPV [80]. Our results showing two groups of viruses in terms of epidemiological dynamics corresponding to the two groups with different climatic sensitivities, raise the question of whether there is a causal relation between the different climatic sensitivities and the different epidemiological patterns, and whether, if the former is true, this will still have any effect after the COVID-19 pandemic. Alternative hypotheses might include the possibility of increased likelihood of infection with a virus from the second group after infection with a virus from the first, but experimental studies on transmission and survival of these different NIRVs, as well as epidemiological studies, including longitudinal cohorts, would be needed to test them.

In summary, we have shown evidence indicating that mobility patterns have played a very relevant role in shaping the dynamics of respiratory viruses along this period, prevailing over the effects of weather conditions. Our results reinforce the argument that the seasonal epidemiological dynamics of respiratory viruses are driven by a complex system of interactions between the different factors, which probably led to an equilibrium that was disturbed, and perhaps permanently altered, by the COVID-19 pandemic. Thus, future research will profit from more research on NIRVs and from the implementation of more complex mechanistic models that could tackle these questions.

## Data Availability

All data used in this manuscript is publicly available.

https://www.cdc.gov/surveillance/nrevss/

https://www.cdc.gov/flu/weekly/overview.htm

https://www.canada.ca/en/public-health/services/surveillance/respiratory-virus-detections-canada.html

https://www.canada.ca/en/public-health/services/diseases/flu-influenza/influenza-surveillance/weekly-influenza-reports.html

https://www.cdc.gov/flu/weekly/overview.htm#ILINet

https://www.google.com/covid19/mobility/

https://www.bts.gov/covid-19

## Supporting information

### Supplementary figures

**S1 Fig**. Total number of test performed for each virus in Canada between 2016 and 2023 and Pearson correlation coefficients between the two incidence proxies.

**S2 Fig**. Incidence for each virus analized, weather and mobility time series and correlations in the USA.

**S3 Fig**. Incidence for each virus analized, weather and mobility time series and correlations in Canada.

**S4 Fig**. Correlation and PCA analysis for the different weather variables for independent variable selection.

**S5 Fig**. Regression coefficients and pseudo-R^2^ for the temperature-RH-AC model in the pre-COVID-19 pandemic period limited to the 16-17 and 17-18 seasons in Canada and the USA.

**S6 Fig**. Regression analysis including the population at home variable.

**S7 Fig**. Regression analysis including the residential time variable.

### Supplementary tables

**S1 Table**. Models for the weather analysis for all viruses in the pre-COVID 19 pandemic period in Canada and the USA.

**S2 Table**. Models for the weather analysis for all viruses in the post-COVID 19 pandemic period in Canada and the USA.

**S3 Table**. Models for the mobility analysis (number of trips) for all viruses in the pre and post-COVID 19 pandemic period in the USA.

**S4 Table**. Models for the mobility analysis (population at home) for all viruses in the post-COVID 19 pandemic period in Canada and the USA.

**S5 Table**. Models for the mobility analysis (transit stations) for all viruses in the pre and post-COVID 19 pandemic period in the USA.

**S6 Table**. Models for the mobility analysis (residential times) for all viruses in the post-COVID 19 pandemic period in Canada and the USA.

## Acknowledgments

The authors would like to thank all members of the SPAC lab for comments and critical reading of the manuscript, Pedro Rio for initial discussions, and Raquel Guiomar and Ana Paula Rodrigues from the Instituto Nacional de Saúde Doutor Ricado Jorge, for access to preliminary data. This work was funded by FCT grant DSAIPA/AI/0087/2018.

