## Supplementary figures 1-7 for "The effects of weather and mobility on respiratory viruses dynamics before and after the COVID-19 pandemic"

**A.**

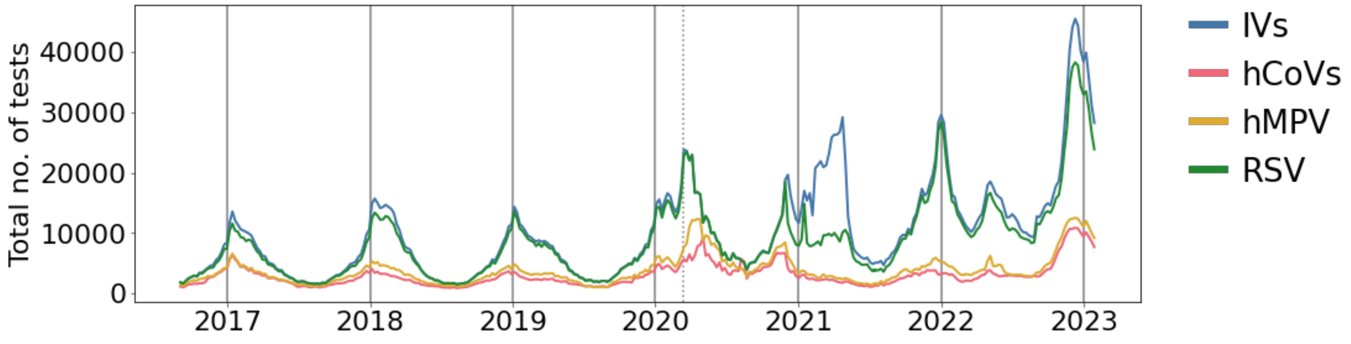


**B.**

| Virus | Correlation coefficient |
|---|---|
| IVA | 0.809012 |
| IVB | 0.917524 |
| hCoVs | 0.802461 |
| hMPV | 0.785859 |
| RSV | 0.743064 |

**S1 Fig. (A)** Total number of test performed in Canada for all viruses from September 2016 until January 2023. Note the number of test for IVs is not reported individually but for both IVA and IVB together. Solid grey vertical lines, 1st January; dotted grey vertical line, WHO pandemic declaration in March 11th, 2020, which approximately marks the separation between pre- and post-COVID19 pandemic periods. **(B)** Pearson correlation coefficient for each virus incidence (calculated with the positivity rate and ILI outpatient rates) and the incidence proxi calculated from the number of positive test and the average Canadian population between 2016 and 2023.

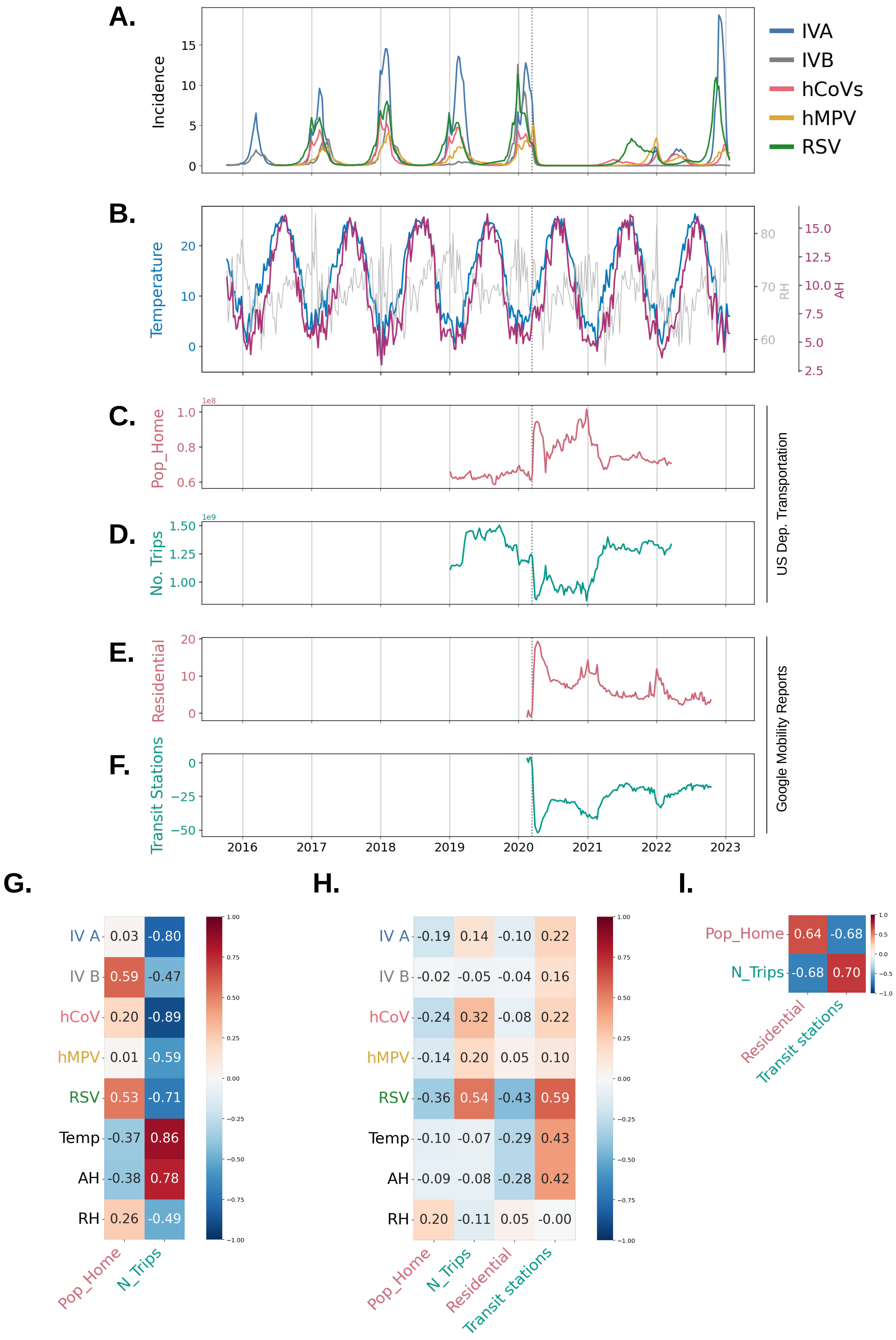


**S2 Fig. (A)** Incidence time series for all viruses and **(B)** temperature, AH and RH time series in the USA from 2015 to 2023. **(C)** Population at home time series and **(D)** number of trips time series for the USA from 2019 to 2022. **(E)** Residential time time series and **(F)** transit station visitors time series for the USA from 2020 to 2022. **(G)** Pearson correlation coefficients between incidence of the different viruses, weather and mobility variables for the USA from 2019 to 2020 (pre-COVID19 ). **(H)** Pearson correlation coefficients between incidence of the different viruses, weather and mobility variables for the USA from 2020 to 2022 (post-COVID19). **(I)** Pearson correlation coefficients between the different mobility measures,for the USA between 2020 and 2022 (post-COVID19).

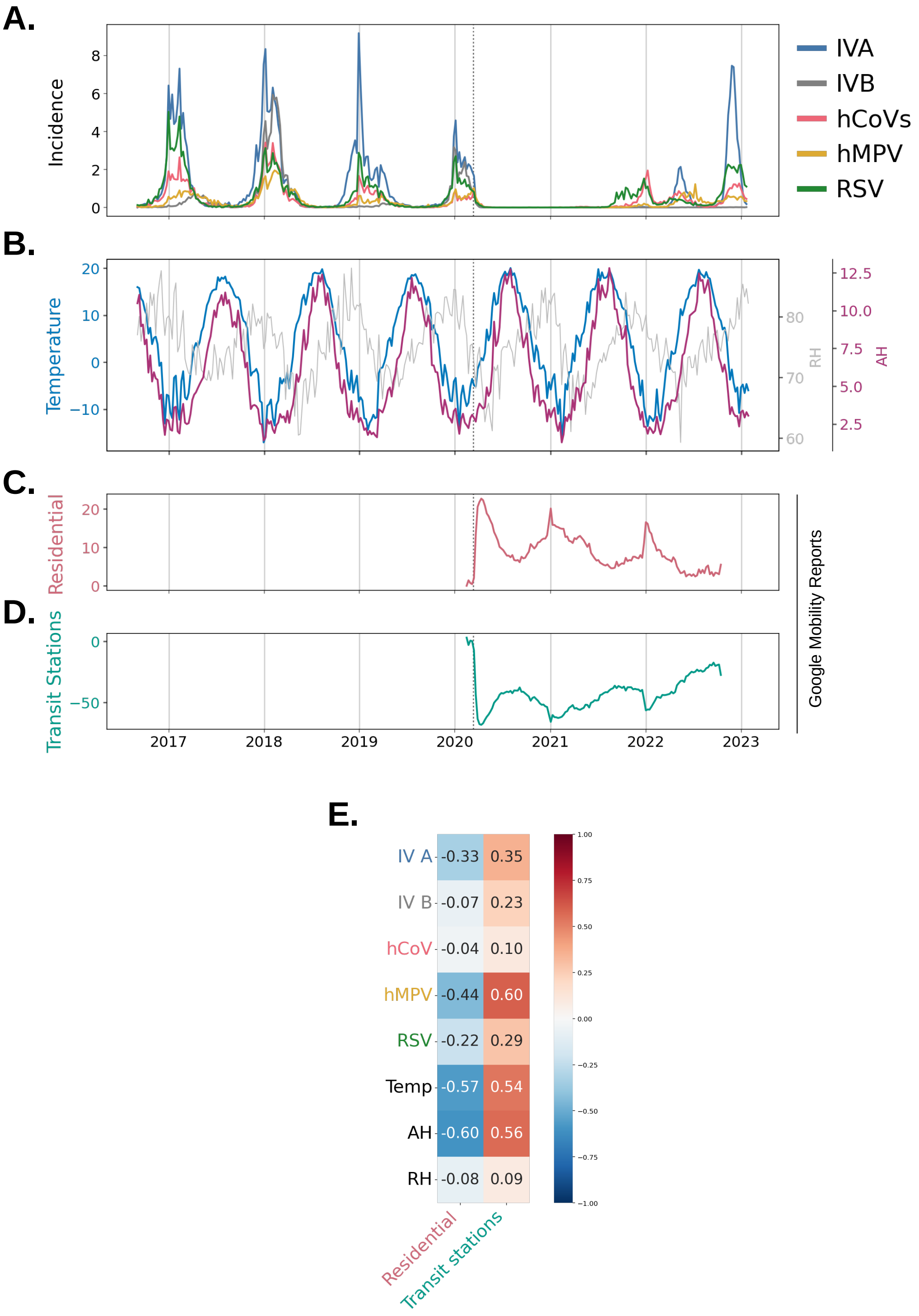


**S3 Fig.** **(A)** Incidence time series for all viruses in the Canada between 2016 and 2013. **(B)** Temperature, AH and RH time series for Canada between 2016 and 2013. **(C)** Residential time time series and **(D)** Transit station visitors time series for Canada between 2020 and 2022. **(E)** Pearson correlation coefficients between the incidence of the different viruses, weather and different mobility measures between between 2020 and 2022 (post-COVID19).

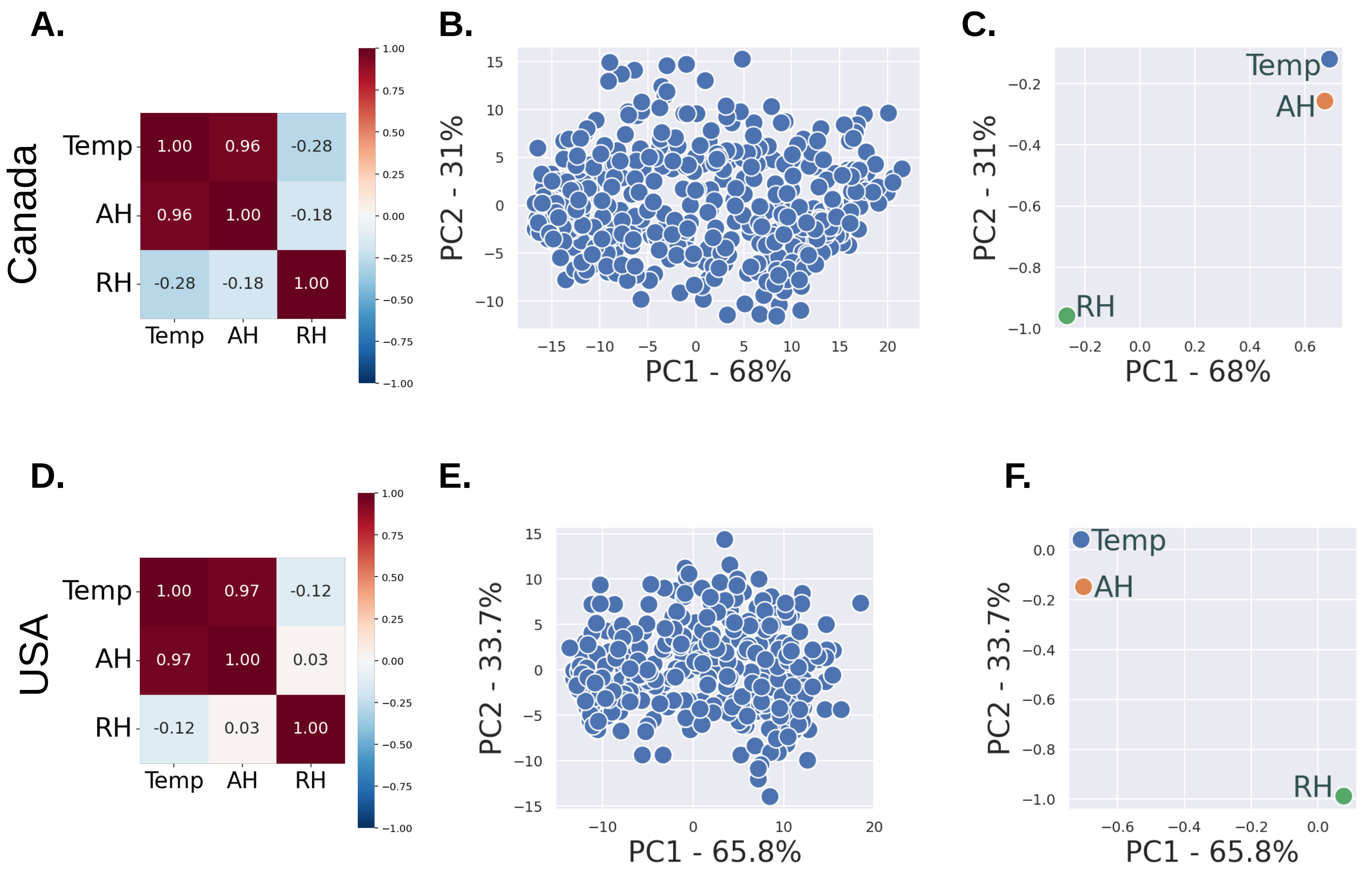


**S4 Fig. (A)** and **(D)** Pearson correlation coefficients for the standarized weather time series in Canada and the USA, respectively, for the whole study period. **(B)** and **(E)** principal component 1 (PC1) and principal component 2 (PC2) for the weather variables in Canada and the USA, respectively. The percentage of variation explained by each PC is shown in the axis. **(C)** and **(F)** loading plots showing the correlation coefficients between each weather variable and PC1 and PC2, (i.e., the elements of the eigenvector) for Canada and the USA, respectively.

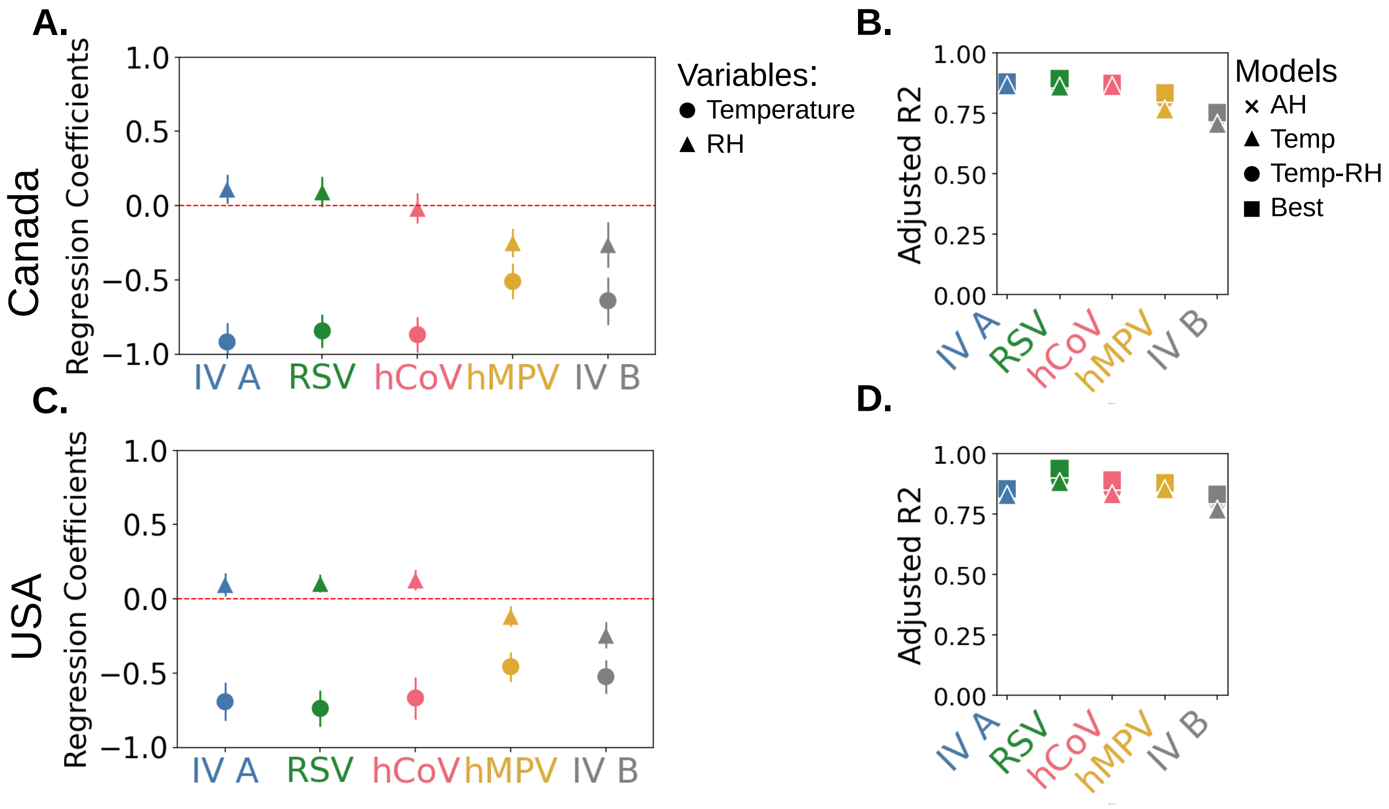


**S5 Fig.** **(A)** and **(C)** regression coefficients for the temperature-RH model with 95% confidence intervals in the pre-COVID19 period for seasons 16-17 and 17-18 in Canada and the USA, respectively. Coefficients for the AC term are not represented. **(B)** and **(D)** **p**seudo-$R^2$ for the best model (square), temperature (triangle), AH (cross) and temperature-RH (circle) models for seasons 16-17 and 17-18 in Canada and the USA, respectively.

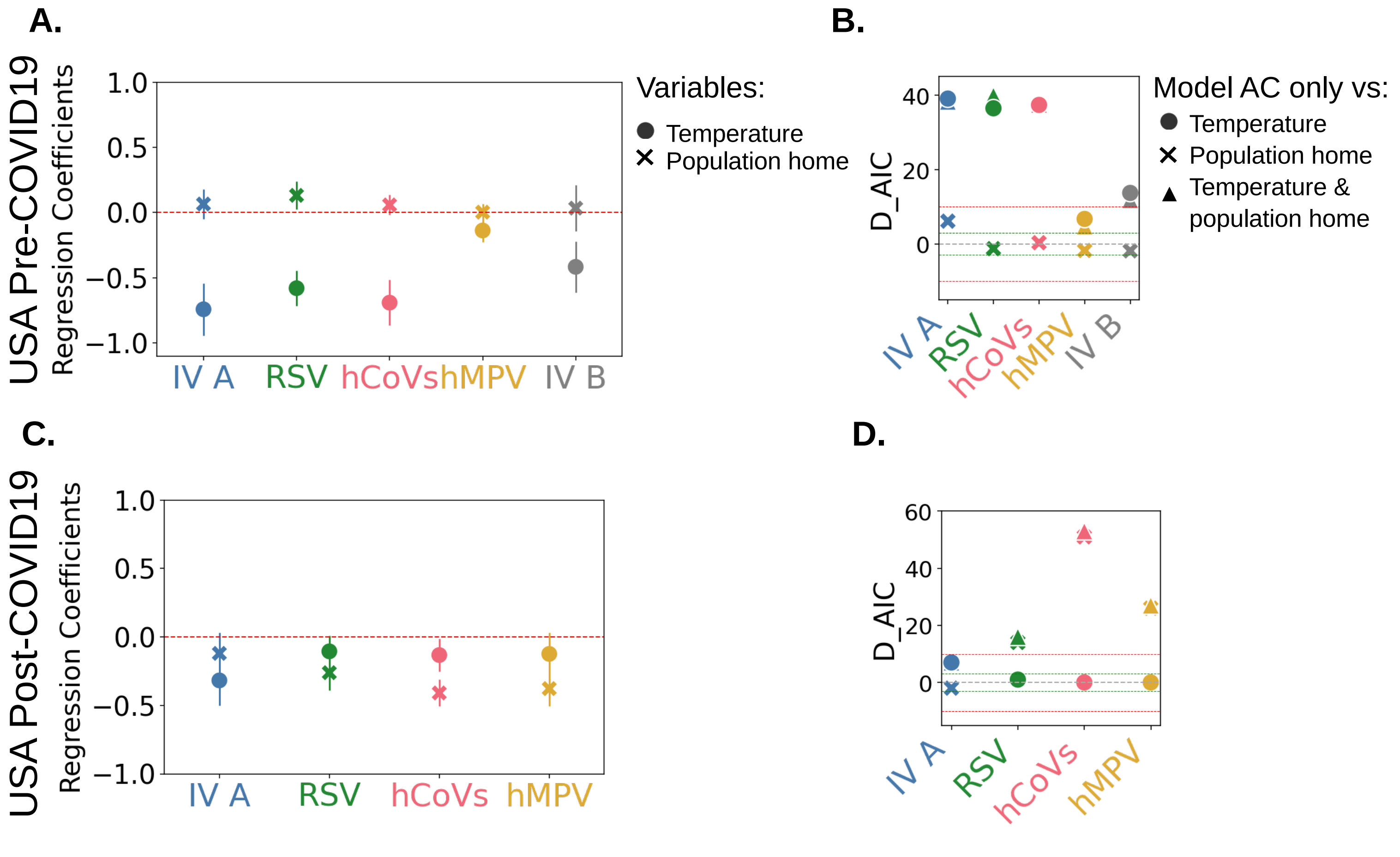


**S6 Fig.** **(A)** and **(C)** regression coefficients for the temperature (circle) and population at home (cross) model with 95% confidence intervals for USA pre- and post-COVID pandemic periods, respectively. **(B)** and **(D)** AIC difference (D_AIC) between the AC only model and the temperature model (circle), the population at home only model (cross) and the temperature-population at home model (triangle) for USA pre- and post-COVID pandemic periods, respectively.

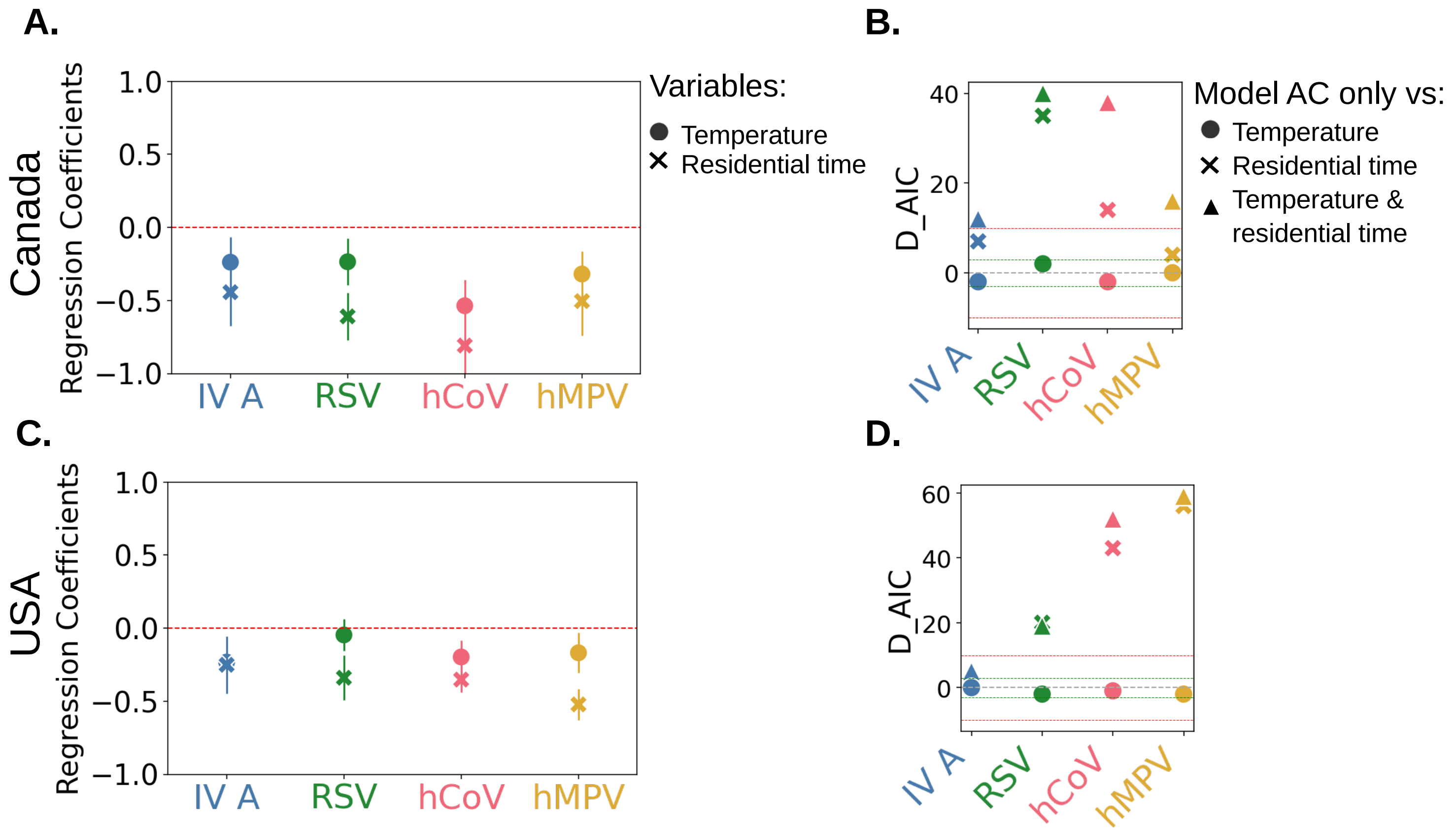


**S7 Fig. (A)** and **(C)** regression coefficients for the temperature (circle) and residential time (cross) model with 95% confidence intervals for the post-COVID19 pandemic period in Canada and the USA, respectively. **(B)** and **(D)** AIC difference (D_AIC) between the AC only model and the temperature only model (circle), the residential time only model (cross) and the temperature-residential time model (triangle) for the post-COVID19 pandemic period in Canada and the USA, respectively
