## Supplementary tables 3-6 for "The effects of weather and mobility on respiratory viruses dynamics before and after the COVID-19 pandemic"

**S3 Table. Regression models results for the number of trips analysis for the USA.**

Temp, temperature; None, no weather variable; Trips, number of trips; p_r2, pseudo $R^2$, Var, variable, Coef, regression coefficient; pV

Only the weather and mobility coefficients are shown.

### Pre-COVID19 pandemic

| Virus | Model | AIC | p_r2 | Var1 | Coef1 | pVal1 | Var2 | Coef2 | pVal2 | Var3 | Coef3 | pVal3 |
|---|---|---|---|---|---|---|---|---|---|---|---|---|
| ***IVA*** | ***AH_RH_Yt-1*** | ***-728,60*** | ***0,90*** | ***AH*** | ***-0,82*** | ***0,00*** | ***RH*** | ***0,12*** | ***0,00*** | | | |
| | ***Temp_Yt-1*** | ***-727,10*** | ***0,89*** | ***Temp*** | ***-0,77*** | ***0,00*** | | | | | | |
| | ***AH_RH_Yt-1_Trips*** | ***-726,60*** | ***0,90*** | ***AH*** | ***-0,80*** | ***0,00*** | ***RH*** | ***0,11*** | ***0,05*** | ***Trips*** | ***-0,03*** | ***0,78*** |
| | ***Temp_RH_Yt-1*** | ***-725,50*** | ***0,89*** | ***Temp*** | ***-0,76*** | ***0,00*** | ***RH*** | ***0,03*** | ***0,55*** | | | |
| | ***Temp_Yt-1_Trips*** | ***-725,10*** | ***0,89*** | ***Temp*** | ***-0,77*** | ***0,00*** | ***Trips*** | ***0,00*** | ***0,97*** | | | |
| | ***AH_Yt-1_Trips*** | ***-725,10*** | ***0,89*** | ***AH*** | ***-0,69*** | ***0,00*** | ***Trips*** | ***-0,19*** | ***0,05*** | | | |
| | Temp_RH_Yt-1_Trips | -723,60 | 0,89 | Temp | -0,80 | 0,00 | RH | 0,04 | 0,46 | Trips | 0,06 | 0,67 |
| | AH_Yt-1 | -722,90 | 0,88 | AH | -0,79 | 0,00 | | | | | | |
| | None_Yt-1_Trips | -702,70 | 0,84 | Trips | -0,50 | 0,00 | | | | | | |
| | None_Yt-1 | -688,00 | 0,79 | Yt-1 | 1,02 | 0,00 | | | | | | |
| ***RSV*** | ***AH_RH_Yt-1_Trips*** | ***-492,50*** | ***0,94*** | ***AH*** | ***-0,72*** | ***0,00*** | ***RH*** | ***0,27*** | ***0,00*** | ***Trips*** | ***0,15*** | ***0,01*** |
| | AH_RH_Yt-1 | -488,80 | 0,93 | AH | -0,61 | 0,00 | RH | 0,23 | 0,00 | | | |
| | Temp_RH_Yt-1_Trips | -484,00 | 0,93 | Temp | -0,66 | 0,00 | RH | 0,15 | 0,00 | Trips | 0,22 | 0,00 |
| | Temp_RH_Yt-1 | -478,10 | 0,91 | Temp | -0,51 | 0,00 | RH | 0,13 | 0,01 | | | |
| | Temp_Yt-1_Trips | -474,40 | 0,90 | Temp | -0,62 | 0,00 | Trips | 0,16 | 0,05 | | | |
| | Temp_Yt-1 | -472,90 | 0,90 | Temp | -0,51 | 0,00 | | | | | | |
| | AH_Yt-1 | -465,90 | 0,88 | AH | -0,49 | 0,00 | | | | | | |
| | AH_Yt-1_Trips | -464,00 | 0,88 | AH | -0,48 | 0,00 | Trips | -0,02 | 0,82 | | | |
| | None_Yt-1_Trips | -443,60 | 0,79 | Trips | -0,31 | 0,00 | | | | | | |
| | None_Yt-1 | -436,40 | 0,74 | Yt-1 | 0,61 | 0,00 | | | | | | |
| ***hCoVs*** | ***AH_RH_Yt-1_Trips*** | ***-850,10*** | ***0,93*** | ***AH*** | ***-0,95*** | ***0,00*** | ***RH*** | ***0,18*** | ***0,00*** | ***Trips*** | ***0,25*** | ***0,02*** |
| | AH_RH_Yt-1 | -846,80 | 0,93 | AH | -0,80 | 0,00 | RH | 0,12 | 0,00 | | | |
| | Temp_RH_Yt-1_Trips | -842,90 | 0,92 | Temp | -0,92 | 0,00 | RH | 0,11 | 0,01 | Trips | 0,37 | 0,00 |
| | AH_Yt-1 | -839,30 | 0,91 | AH | -0,73 | 0,00 | | | | | | |
| | Temp_Yt-1_Trips | -838,70 | 0,92 | Temp | -0,82 | 0,00 | Trips | 0,21 | 0,06 | | | |
| | Temp_Yt-1 | -837,60 | 0,91 | Temp | -0,70 | 0,00 | | | | | | |
| | Temp_RH_Yt-1 | -837,40 | 0,91 | Temp | -0,70 | 0,00 | RH | 0,06 | 0,18 | | | |
| | AH_Yt-1_Trips | -837,30 | 0,91 | AH | -0,73 | 0,00 | Trips | 0,01 | 0,95 | | | |
| | None_Yt-1_Trips | -803,60 | 0,85 | Trips | -0,35 | 0,02 | | | | | | |
| | None_Yt-1 | -800,20 | 0,83 | Yt-1 | 1,04 | 0,00 | | | | | | |
| ***IVB*** | ***Temp_RH_Yt-1_Trips*** | ***-784,90*** | ***0,83*** | ***Temp*** | ***-1,17*** | ***0,00*** | ***RH*** | ***0,16*** | ***0,01*** | ***Trips*** | ***0,82*** | ***0,00*** |
| | Temp_Yt-1_Trips | -780,80 | 0,81 | Temp | -1,09 | 0,00 | Trips | 0,70 | 0,00 | | | |
| | AH_RH_Yt-1_Trips | -777,90 | 0,81 | AH | -1,03 | 0,00 | RH | 0,26 | 0,00 | Trips | 0,63 | 0,00 |
| | AH_Yt-1_Trips | -769,10 | 0,77 | AH | -0,77 | 0,00 | Trips | 0,37 | 0,00 | | | |
| | AH_Yt-1 | -763,00 | 0,74 | AH | -0,44 | 0,00 | | | | | | |
| | Temp_Yt-1 | -762,30 | 0,74 | Temp | -0,41 | 0,00 | | | | | | |
| | AH_RH_Yt-1 | -761,70 | 0,74 | AH | -0,44 | 0,00 | RH | 0,06 | 0,39 | | | |
| | Temp_RH_Yt-1 | -760,30 | 0,74 | Temp | -0,41 | 0,00 | RH | 0,01 | 0,85 | | | |
| | None_Yt-1 | -748,60 | 0,66 | Yt-1 | 0,78 | 0,00 | | | | | | |
| | None_Yt-1_Trips | -748,40 | 0,67 | Trips | -0,14 | 0,17 | | | | | | |
| ***hMPV*** | ***AH_RH_Yt-1*** | ***-554,70*** | ***0,89*** | ***AH*** | ***-0,12*** | ***0,01*** | ***RH*** | ***0,07*** | ***0,02*** | | | |
| | ***AH_RH_Yt-1_Trips*** | ***-554,40*** | ***0,89*** | ***AH*** | ***-0,18*** | ***0,01*** | ***RH*** | ***0,11*** | ***0,01*** | ***Trips*** | ***0,09*** | ***0,19*** |
| | ***Temp_RH_Yt-1_Trips*** | ***-554,10*** | ***0,89*** | ***Temp*** | ***-0,20*** | ***0,01*** | ***RH*** | ***0,08*** | ***0,03*** | ***Trips*** | ***0,12*** | ***0,11*** |
| | ***Temp_RH_Yt-1*** | ***-553,70*** | ***0,89*** | ***Temp*** | ***-0,11*** | ***0,02*** | ***RH*** | ***0,05*** | ***0,14*** | | | |
| | ***Temp_Yt-1*** | ***-553,60*** | ***0,88*** | ***Temp*** | ***-0,14*** | ***0,00*** | | | | | | |
| | ***Temp_Yt-1_Trips*** | ***-551,80*** | ***0,88*** | ***Temp*** | ***-0,16*** | ***0,02*** | ***Trips*** | ***0,03*** | ***0,64*** | | | |
| | AH_Yt-1 | -551,20 | 0,87 | AH | -0,13 | 0,01 | | | | | | |
| | AH_Yt-1_Trips | -549,80 | 0,88 | AH | -0,10 | 0,09 | Trips | -0,04 | 0,46 | | | |
| | None_Yt-1_Trips | -548,80 | 0,87 | Trips | -0,09 | 0,04 | | | | | | |
| | None_Yt-1 | -546,90 | 0,85 | Yt-1 | 0,55 | 0,00 | | | | | | |

**Post-COVID19 pandemic**

| Virus | Model | AIC | p_r2 | Var1 | Coef1 | pVal1 | Var2 | Coef2 | pVal2 | Var3 | Coef3 | pVal3 |
|---|---|---|---|---|---|---|---|---|---|---|---|---|
| ***IVA*** | ***Temp_Yt-1_Trips*** | ***-1780,00*** | ***0,49*** | ***Temp*** | ***-0,32*** | ***0,00*** | ***Trips*** | ***0,20*** | ***0,04*** | | | |
| | ***Temp_RH_Yt-1_Trips*** | ***-1779,00*** | ***0,49*** | ***Temp*** | ***-0,33*** | ***0,00*** | ***RH*** | ***0,08*** | ***0,33*** | ***Trips*** | ***0,23*** | ***0,02*** |
| | ***AH_RH_Yt-1_Trips*** | ***-1778,00*** | ***0,49*** | ***AH*** | ***-0,32*** | ***0,00*** | ***RH*** | ***0,15*** | ***0,08*** | ***Trips*** | ***0,23*** | ***0,02*** |
| | ***Temp_Yt-1*** | ***-1778,00*** | ***0,47*** | ***Temp*** | ***-0,27*** | ***0,00*** | | | | | | |
| | ***AH_Yt-1_Trips*** | ***-1777,00*** | ***0,47*** | ***AH*** | ***-0,28*** | ***0,00*** | ***Trips*** | ***0,16*** | ***0,09*** | | | |
| | AH_Yt-1 | -1776,00 | 0,46 | AH | -0,25 | 0,01 | | | | | | |
| | Temp_RH_Yt-1 | -1776,00 | 0,47 | Temp | -0,27 | 0,00 | RH | 0,01 | 0,92 | | | |
| | AH_RH_Yt-1 | -1775,00 | 0,46 | AH | -0,26 | 0,01 | RH | 0,07 | 0,41 | | | |
| | None_Yt-1 | -1771,00 | 0,42 | Yt-1 | 0,57 | 0,00 | | | | | | |
| | None_Yt-1_Trips | -1770,00 | 0,43 | Trips | 0,12 | 0,21 | | | | | | |
| ***RSV*** | ***AH_RH_Yt-1_Trips*** | ***-1629,00*** | ***0,87*** | ***AH*** | ***-0,04*** | ***0,53*** | ***RH*** | ***0,11*** | ***0,03*** | ***Trips*** | ***0,63*** | ***0,00*** |
| | ***None_Yt-1_Trips*** | ***-1628,00*** | ***0,86*** | ***Trips*** | ***0,61*** | ***0,00*** | | | | | | |
| | ***Temp_RH_Yt-1_Trips*** | ***-1628,00*** | ***0,87*** | ***Temp*** | ***-0,02*** | ***0,72*** | ***RH*** | ***0,11*** | ***0,04*** | ***Trips*** | ***0,64*** | ***0,00*** |
| | ***Temp_Yt-1_Trips*** | ***-1626,00*** | ***0,86*** | ***Temp*** | ***-0,05*** | ***0,43*** | ***Trips*** | ***0,59*** | ***0,00*** | | | |
| | ***AH_Yt-1_Trips*** | ***-1626,00*** | ***0,86*** | ***AH*** | ***-0,04*** | ***0,51*** | ***Trips*** | ***0,59*** | ***0,00*** | | | |
| | AH_Yt-1 | -1580,00 | 0,78 | AH | -0,12 | 0,05 | | | | | | |
| | Temp_Yt-1 | -1579,00 | 0,78 | Temp | -0,11 | 0,07 | | | | | | |
| | None_Yt-1 | -1578,00 | 0,77 | Yt-1 | 1,07 | 0,00 | | | | | | |
| | Temp_RH_Yt-1 | -1578,00 | 0,78 | Temp | -0,12 | 0,06 | RH | -0,05 | 0,49 | | | |
| | AH_RH_Yt-1 | -1578,00 | 0,78 | AH | -0,12 | 0,05 | RH | -0,02 | 0,73 | | | |
| ***hCoVs*** | ***None_Yt-1_Trips*** | ***-1687,00*** | ***0,86*** | ***Trips*** | ***0,73*** | ***0,00*** | | | | | | |
| | ***Temp_Yt-1_Trips*** | ***-1685,00*** | ***0,86*** | ***Temp*** | ***-0,02*** | ***0,70*** | ***Trips*** | ***0,72*** | ***0,00*** | | | |
| | ***AH_Yt-1_Trips*** | ***-1685,00*** | ***0,86*** | ***AH*** | ***-0,03*** | ***0,52*** | ***Trips*** | ***0,72*** | ***0,00*** | | | |
| | ***Temp_RH_Yt-1_Trips*** | ***-1685,00*** | ***0,86*** | ***Temp*** | ***-0,02*** | ***0,71*** | ***RH*** | ***-0,06*** | ***0,13*** | ***Trips*** | ***0,71*** | ***0,00*** |
| | ***AH_RH_Yt-1_Trips*** | ***-1685,00*** | ***0,86*** | ***AH*** | ***-0,01*** | ***0,75*** | ***RH*** | ***-0,06*** | ***0,16*** | ***Trips*** | ***0,72*** | ***0,00*** |
| | Temp_RH_Yt-1 | -1563,00 | 0,55 | Temp | -0,12 | 0,11 | RH | -0,20 | 0,01 | | | |
| | AH_RH_Yt-1 | -1562,00 | 0,55 | AH | -0,10 | 0,22 | RH | -0,18 | 0,02 | | | |
| | None_Yt-1 | -1558,00 | 0,52 | Yt-1 | 0,51 | 0,00 | | | | | | |
| | Temp_Yt-1 | -1558,00 | 0,52 | Temp | -0,10 | 0,20 | | | | | | |
| | AH_Yt-1 | -1558,00 | 0,53 | AH | -0,11 | 0,14 | | | | | | |
| ***IVB*** | ***AH_RH_Yt-1_Trips*** | ***-2190,00*** | ***0,92*** | ***AH*** | ***-0,20*** | ***0,00*** | ***RH*** | ***0,09*** | ***0,04*** | ***Trips*** | ***-0,34*** | ***0,00*** |
| | ***Temp_Yt-1_Trips*** | ***-2187,00*** | ***0,92*** | ***Temp*** | ***-0,19*** | ***0,00*** | ***Trips*** | ***-0,38*** | ***0,00*** | | | |
| | ***AH_Yt-1_Trips*** | ***-2187,00*** | ***0,92*** | ***AH*** | ***-0,20*** | ***0,00*** | ***Trips*** | ***-0,40*** | ***0,00*** | | | |
| | ***Temp_RH_Yt-1_Trips*** | ***-2187,00*** | ***0,92*** | ***Temp*** | ***-0,18*** | ***0,00*** | ***RH*** | ***0,05*** | ***0,26*** | ***Trips*** | ***-0,35*** | ***0,00*** |
| | None_Yt-1_Trips | -2174,00 | 0,91 | Trips | -0,38 | 0,00 | | | | | | |
| | AH_RH_Yt-1 | -2151,00 | 0,88 | AH | -0,17 | 0,00 | RH | 0,24 | 0,00 | | | |
| | Temp_RH_Yt-1 | -2148,00 | 0,88 | Temp | -0,14 | 0,01 | RH | 0,21 | 0,00 | | | |
| | Temp_Yt-1 | -2129,00 | 0,86 | Temp | -0,16 | 0,01 | | | | | | |
| | AH_Yt-1 | -2126,00 | 0,85 | AH | -0,12 | 0,06 | | | | | | |
| | None_Yt-1 | -2124,00 | 0,85 | Yt-1 | 0,66 | 0,00 | | | | | | |
| ***hMPV*** | ***None_Yt-1_Trips*** | ***-1719,00*** | ***0,68*** | ***Trips*** | ***0,51*** | ***0,00*** | | | | | | |
| | ***Temp_Yt-1_Trips*** | ***-1717,00*** | ***0,68*** | ***Temp*** | ***-0,05*** | ***0,51*** | ***Trips*** | ***0,50*** | ***0,00*** | | | |
| | ***AH_Yt-1_Trips*** | ***-1717,00*** | ***0,68*** | ***AH*** | ***-0,05*** | ***0,56*** | ***Trips*** | ***0,50*** | ***0,00*** | | | |
| | ***AH_RH_Yt-1_Trips*** | ***-1717,00*** | ***0,69*** | ***AH*** | ***-0,08*** | ***0,36*** | ***RH*** | ***0,09*** | ***0,19*** | ***Trips*** | ***0,50*** | ***0,00*** |
| | ***Temp_RH_Yt-1_Trips*** | ***-1716,00*** | ***0,69*** | ***Temp*** | ***-0,06*** | ***0,48*** | ***RH*** | ***0,07*** | ***0,26*** | ***Trips*** | ***0,50*** | ***0,00*** |
| | None_Yt-1 | -1678,00 | 0,53 | Yt-1 | 0,72 | 0,00 | | | | | | |
| | Temp_Yt-1 | -1678,00 | 0,54 | Temp | -0,13 | 0,14 | | | | | | |
| | AH_Yt-1 | -1678,00 | 0,53 | AH | -0,12 | 0,20 | | | | | | |
| | Temp_RH_Yt-1 | -1676,00 | 0,54 | Temp | -0,13 | 0,14 | RH | 0,01 | 0,92 | | | |
| | AH_RH_Yt-1 | -1676,00 | 0,53 | AH | -0,12 | 0,19 | RH | 0,04 | 0,66 | | | |

### S4 Table. Regression models results for the population at home analysis for the USA.

Temp, temperature;  None, no weather variable; Phome, population at home; p_r2, pseudo $R^2$, Var, variable, Coef, regression coefficient; pVal, Only the weather and mobility coefficients are shown.

#### Pre-COVID19 pandemic

| Virus | Model | AIC | p_r2 | Var2 | Coef2 | pVal2 | Var3 | Coef3 | pVal3 | Var3 | Coef3 | pVal3 |
|---|---|---|---|---|---|---|---|---|---|---|---|---|
| ***IVA*** | ***AH_RH_Yt-1*** | ***-728,60*** | ***0,90*** | ***AH*** | ***-0,82*** | ***0,00*** | ***RH*** | ***0,12*** | ***0,00*** | | | |
| | ***Temp_Yt-1*** | ***-727,10*** | ***0,89*** | ***Temp*** | ***-0,77*** | ***0,00*** | | | | | | |
| | ***AH_RH_Yt-1_PHome*** | ***-726,90*** | ***0,90*** | ***AH*** | ***-0,80*** | ***0,00*** | ***RH*** | ***0,12*** | ***0,01*** | ***PHome*** | ***0,04*** | ***0,54*** |
| | ***Temp_Yt-1_PHome*** | ***-726,30*** | ***0,89*** | ***Temp*** | ***-0,74*** | ***0,00*** | ***PHome*** | ***0,07*** | ***0,26*** | | | |
| | Temp_RH_Yt-1 | -725,50 | 0,89 | Temp | -0,76 | 0,00 | RH | 0,03 | 0,55 | | | |
| | Temp_RH_Yt-1_PHome | -724,60 | 0,89 | Temp | -0,73 | 0,00 | RH | 0,03 | 0,60 | PHome | 0,06 | 0,28 |
| | AH_Yt-1 | -722,90 | 0,88 | AH | -0,79 | 0,00 | | | | | | |
| | AH_Yt-1_PHome | -722,00 | 0,88 | AH | -0,76 | 0,00 | PHome | 0,06 | 0,30 | | | |
| | None_Yt-1_PHome | -694,10 | 0,81 | PHome | 0,21 | 0,00 | | | | | | |
| | None_Yt-1 | -688,00 | 0,79 | Yt-1 | 1,02 | 0,00 | | | | | | |
| ***RSV*** | ***AH_RH_Yt-1_PHome*** | ***-493,30*** | ***0,94*** | ***AH*** | ***-0,68*** | ***0,00*** | ***RH*** | ***0,25*** | ***0,00*** | ***PHome*** | ***0,11*** | ***0,01*** |
| | AH_RH_Yt-1 | -488,80 | 0,93 | AH | -0,61 | 0,00 | RH | 0,23 | 0,00 | | | |
| | Temp_RH_Yt-1_PHome | -483,20 | 0,93 | Temp | -0,59 | 0,00 | RH | 0,13 | 0,00 | PHome | 0,14 | 0,01 |
| | Temp_RH_Yt-1 | -478,10 | 0,91 | Temp | -0,51 | 0,00 | RH | 0,13 | 0,01 | | | |
| | Temp_Yt-1_PHome | -476,30 | 0,91 | Temp | -0,58 | 0,00 | | | | | | |
| | Temp_Yt-1 | -472,90 | 0,90 | Temp | -0,51 | 0,00 | | | | | | |
| | AH_Yt-1 | -465,90 | 0,88 | AH | -0,49 | 0,00 | | | | | | |
| | AH_Yt-1_PHome | -465,60 | 0,88 | AH | -0,53 | 0,00 | PHome | 0,08 | 0,20 | | | |
| | None_Yt-1 | -436,40 | 0,74 | Yt-1 | 0,61 | 0,00 | | | | | | |
| | None_Yt-1_PHome | -435,10 | 0,74 | PHome | -0,07 | 0,41 | | | | | | |
| ***hCoVs*** | ***AH_RH_Yt-1*** | ***-846,80*** | ***0,93*** | ***AH*** | ***-0,80*** | ***0,00*** | ***RH*** | ***0,12*** | ***0,00*** | | | |
| | ***AH_RH_Yt-1_PHome*** | ***-845,10*** | ***0,93*** | ***AH*** | ***-0,80*** | ***0,00*** | ***RH*** | ***0,12*** | ***0,00*** | ***PHome*** | ***0,02*** | ***0,60*** |
| | AH_Yt-1 | -839,30 | 0,91 | AH | -0,73 | 0,00 | | | | | | |
| | AH_Yt-1_PHome | -838,60 | 0,92 | AH | -0,72 | 0,00 | PHome | 0,04 | 0,24 | | | |
| | Temp_Yt-1_PHome | -837,70 | 0,91 | Temp | -0,69 | 0,00 | PHome | 0,06 | 0,13 | | | |
| | Temp_Yt-1 | -837,60 | 0,91 | Temp | -0,70 | 0,00 | | | | | | |
| | Temp_RH_Yt-1 | -837,40 | 0,91 | Temp | -0,70 | 0,00 | RH | 0,06 | 0,18 | | | |
| | Temp_RH_Yt-1_PHome | -836,90 | 0,92 | Temp | -0,70 | 0,00 | RH | 0,05 | 0,27 | PHome | 0,05 | 0,21 |
| | None_Yt-1_PHome | -800,50 | 0,84 | PHome | 0,08 | 0,12 | | | | | | |
| | None_Yt-1 | -800,20 | 0,83 | Yt-1 | 1,04 | 0,00 | | | | | | |
| ***IVB*** | ***AH_Yt-1*** | ***-763,00*** | ***0,74*** | ***AH*** | ***-0,44*** | ***0,00*** | | | | | | |
| | ***Temp_Yt-1*** | ***-762,30*** | ***0,74*** | ***Temp*** | ***-0,41*** | ***0,00*** | | | | | | |
| | ***AH_RH_Yt-1*** | ***-761,70*** | ***0,74*** | ***AH*** | ***-0,44*** | ***0,00*** | ***RH*** | ***0,06*** | ***0,39*** | | | |
| | ***AH_Yt-1_PHome*** | ***-761,00*** | ***0,74*** | ***AH*** | ***-0,44*** | ***0,00*** | ***PHome*** | ***0,03*** | ***0,78*** | | | |
| | ***Temp_Yt-1_PHome*** | ***-760,40*** | ***0,74*** | ***Temp*** | ***-0,42*** | ***0,00*** | ***PHome*** | ***0,03*** | ***0,71*** | | | |
| | ***Temp_RH_Yt-1*** | ***-760,30*** | ***0,74*** | ***Temp*** | ***-0,41*** | ***0,00*** | ***RH*** | ***0,01*** | ***0,85*** | | | |
| | AH_RH_Yt-1_PHome | -759,70 | 0,74 | AH | -0,44 | 0,00 | RH | 0,06 | 0,41 | PHome | 0,02 | 0,87 |
| | Temp_RH_Yt-1_PHome | -758,40 | 0,74 | Temp | -0,41 | 0,00 | RH | 0,01 | 0,90 | PHome | 0,03 | 0,72 |
| | None_Yt-1 | -748,60 | 0,66 | Yt-1 | 0,78 | 0,00 | | | | | | |
| | None_Yt-1_PHome | -746,70 | 0,66 | PHome | 0,02 | 0,87 | | | | | | |
| ***hMPV*** | ***AH_RH_Yt-1*** | ***-554,70*** | ***0,89*** | ***AH*** | ***-0,12*** | ***0,01*** | ***RH*** | ***0,07*** | ***0,02*** | | | |
| | ***Temp_RH_Yt-1*** | ***-553,70*** | ***0,89*** | ***Temp*** | ***-0,11*** | ***0,02*** | ***RH*** | ***0,05*** | ***0,14*** | | | |
| | ***Temp_Yt-1*** | ***-553,60*** | ***0,88*** | ***Temp*** | ***-0,14*** | ***0,00*** | | | | | | |
| | ***AH_RH_Yt-1_PHome*** | ***-553,00*** | ***0,89*** | ***AH*** | ***-0,13*** | ***0,01*** | ***RH*** | ***0,08*** | ***0,01*** | ***PHome*** | ***-0,02*** | ***0,63*** |
| | ***Temp_RH_Yt-1_PHome*** | ***-551,80*** | ***0,89*** | ***Temp*** | ***-0,11*** | ***0,02*** | ***RH*** | ***0,05*** | ***0,13*** | ***PHome*** | ***-0,01*** | ***0,76*** |
| | Temp_Yt-1_PHome | -551,60 | 0,88 | Temp | -0,14 | 0,00 | PHome | 0,00 | 0,92 | | | |
| | AH_Yt-1 | -551,20 | 0,87 | AH | -0,13 | 0,01 | | | | | | |
| | AH_Yt-1_PHome | -549,30 | 0,87 | AH | -0,13 | 0,01 | PHome | 0,01 | 0,82 | | | |
| | None_Yt-1 | -546,90 | 0,85 | Yt-1 | 0,55 | 0,00 | | | | | | |
| | None_Yt-1_PHome | -545,10 | 0,85 | PHome | 0,02 | 0,61 | | | | | | |

### Post-COVID19 pandemic

| Virus | Model | AIC | p_r2 | Var1 | Coef1 | pVal1 | Var2 | Coef2 | pVal2 | Var3 | Coef3 | pVal3 |
|---|---|---|---|---|---|---|---|---|---|---|---|---|
| IVA | Temp_Yt-1_PHome | -1778,00 | 0,48 | Temp | -0,32 | 0,00 | PHome | -0,12 | 0,12 | | | |
| | Temp_Yt-1 | -1778,00 | 0,47 | Temp | -0,27 | 0,00 | | | | | | |
| | Temp_RH_Yt-1_PHome | -1777,00 | 0,48 | Temp | -0,32 | 0,00 | RH | 0,10 | 0,29 | PHome | -0,17 | 0,06 |
| | AH_RH_Yt-1_PHome | -1777,00 | 0,48 | AH | -0,32 | 0,00 | RH | 0,17 | 0,07 | PHome | -0,17 | 0,05 |
| | AH_Yt-1 | -1776,00 | 0,46 | AH | -0,25 | 0,01 | | | | | | |
| | Temp_RH_Yt-1 | -1776,00 | 0,47 | Temp | -0,27 | 0,00 | RH | 0,01 | 0,92 | | | |
| | AH_Yt-1_PHome | -1775,00 | 0,46 | AH | -0,27 | 0,00 | PHome | -0,09 | 0,25 | | | |
| | AH_RH_Yt-1 | -1775,00 | 0,46 | AH | -0,26 | 0,01 | RH | 0,07 | 0,41 | | | |
| | None_Yt-1 | -1771,00 | 0,42 | Yt-1 | 0,57 | 0,00 | | | | | | |
| | None_Yt-1_PHome | -1769,00 | 0,42 | PHome | -0,04 | 0,56 | | | | | | |
| RSV | Temp_Yt-1_PHome | -1594,00 | 0,81 | Temp | -0,10 | 0,08 | PHome | -0,26 | 0,00 | | | |
| | AH_Yt-1_PHome | -1593,00 | 0,81 | AH | -0,10 | 0,09 | PHome | -0,25 | 0,00 | | | |
| | Temp_RH_Yt-1_PHome | -1593,00 | 0,81 | Temp | -0,09 | 0,12 | RH | 0,08 | 0,25 | PHome | -0,29 | 0,00 |
| | AH_RH_Yt-1_PHome | -1593,00 | 0,82 | AH | -0,10 | 0,08 | RH | 0,10 | 0,14 | PHome | -0,29 | 0,00 |
| | None_Yt-1_PHome | -1592,00 | 0,81 | PHome | -0,28 | 0,00 | | | | | | |
| | AH_Yt-1 | -1580,00 | 0,78 | AH | -0,12 | 0,05 | | | | | | |
| | Temp_Yt-1 | -1579,00 | 0,78 | Temp | -0,11 | 0,07 | | | | | | |
| | None_Yt-1 | -1578,00 | 0,77 | Yt-1 | 1,07 | 0,00 | | | | | | |
| | Temp_RH_Yt-1 | -1578,00 | 0,78 | Temp | -0,12 | 0,06 | RH | -0,05 | 0,49 | | | |
| | AH_RH_Yt-1 | -1578,00 | 0,78 | AH | -0,12 | 0,05 | RH | -0,02 | 0,73 | | | |
| hCoVs | Temp_Yt-1_PHome | -1611,00 | 0,72 | Temp | -0,13 | 0,03 | PHome | -0,41 | 0,00 | | | |
| | AH_Yt-1_PHome | -1611,00 | 0,72 | AH | -0,14 | 0,04 | PHome | -0,40 | 0,00 | | | |
| | Temp_RH_Yt-1_PHome | -1610,00 | 0,72 | Temp | -0,13 | 0,03 | RH | -0,04 | 0,59 | PHome | -0,40 | 0,00 |
| | None_Yt-1_PHome | -1609,00 | 0,71 | PHome | -0,41 | 0,00 | | | | | | |
| | AH_RH_Yt-1_PHome | -1609,00 | 0,72 | AH | -0,14 | 0,04 | RH | -0,01 | 0,91 | PHome | -0,40 | 0,00 |
| | Temp_RH_Yt-1 | -1563,00 | 0,55 | Temp | -0,12 | 0,11 | RH | -0,20 | 0,01 | | | |
| | AH_RH_Yt-1 | -1562,00 | 0,55 | AH | -0,10 | 0,22 | RH | -0,18 | 0,02 | | | |
| | None_Yt-1 | -1558,00 | 0,52 | Yt-1 | 0,51 | 0,00 | | | | | | |
| | Temp_Yt-1 | -1558,00 | 0,52 | Temp | -0,10 | 0,20 | | | | | | |
| | AH_Yt-1 | -1558,00 | 0,53 | AH | -0,11 | 0,14 | | | | | | |
| IVB | AH_Yt-1_PHome | -2182,00 | 0,91 | AH | -0,14 | 0,01 | PHome | 0,22 | 0,00 | | | |
| | Temp_Yt-1_PHome | -2180,00 | 0,91 | Temp | -0,12 | 0,02 | PHome | 0,21 | 0,00 | | | |
| | AH_RH_Yt-1_PHome | -2180,00 | 0,91 | AH | -0,14 | 0,01 | RH | 0,01 | 0,91 | PHome | 0,21 | 0,00 |
| | Temp_RH_Yt-1_PHome | -2179,00 | 0,91 | Temp | -0,12 | 0,02 | RH | -0,03 | 0,61 | PHome | 0,22 | 0,00 |
| | None_Yt-1_PHome | -2177,00 | 0,91 | PHome | 0,22 | 0,00 | | | | | | |
| | AH_RH_Yt-1 | -2151,00 | 0,88 | AH | -0,17 | 0,00 | RH | 0,24 | 0,00 | | | |
| | Temp_RH_Yt-1 | -2148,00 | 0,88 | Temp | -0,14 | 0,01 | RH | 0,21 | 0,00 | | | |
| | Temp_Yt-1 | -2129,00 | 0,86 | Temp | -0,16 | 0,01 | | | | | | |
| | AH_Yt-1 | -2126,00 | 0,85 | AH | -0,12 | 0,06 | | | | | | |
| | None_Yt-1 | -2124,00 | 0,85 | Yt-1 | 0,66 | 0,00 | | | | | | |
| hMPV | Temp_Yt-1_PHome | -1705,00 | 0,64 | Temp | -0,12 | 0,12 | PHome | -0,38 | 0,00 | | | |
| | Temp_RH_Yt-1_PHome | -1705,00 | 0,65 | Temp | -0,12 | 0,13 | RH | 0,11 | 0,12 | PHome | -0,40 | 0,00 |
| | AH_RH_Yt-1_PHome | -1705,00 | 0,65 | AH | -0,14 | 0,09 | RH | 0,13 | 0,05 | PHome | -0,40 | 0,00 |
| | None_Yt-1_PHome | -1704,00 | 0,64 | PHome | -0,39 | 0,00 | | | | | | |
| | AH_Yt-1_PHome | -1704,00 | 0,64 | AH | -0,11 | 0,20 | PHome | -0,38 | 0,00 | | | |
| | None_Yt-1 | -1678,00 | 0,53 | Yt-1 | 0,72 | 0,00 | | | | | | |
| | Temp_Yt-1 | -1678,00 | 0,54 | Temp | -0,13 | 0,14 | | | | | | |
| | AH_Yt-1 | -1678,00 | 0,53 | AH | -0,12 | 0,20 | | | | | | |
| | Temp_RH_Yt-1 | -1676,00 | 0,54 | Temp | -0,13 | 0,14 | RH | 0,01 | 0,92 | | | |
| | AH_RH_Yt-1 | -1676,00 | 0,53 | AH | -0,12 | 0,19 | RH | 0,04 | 0,66 | | | |

### S5 Table. Regression models results for the transit stations analysis (post-COVID19 pandemic).

Temp, temperature; None, no weather variable; Transit, transit stations; p_r2, pseudo $R^2$, Var, variable, Coef, regression coefficient; pVal, p-value

Only the weather and mobility coefficients are shown.

#### Canada

| Virus | Model | AIC | p_r1 | Var1 | Coef1 | pVal1 | Var2 | Coef2 | pVal2 | Var3 | Coef3 | pVal3 |
|---|---|---|---|---|---|---|---|---|---|---|---|---|
| ***IVA*** | ***AH_Yt-1_Transit*** | ***-2939,00*** | ***0,51*** | ***AH*** | ***-0,17*** | ***0,04*** | ***Transit*** | ***0,37*** | ***0,00*** | | | |
| | ***AH_RH_Yt-1_Transit*** | ***-2938,00*** | ***0,51*** | ***AH*** | ***-0,18*** | ***0,03*** | ***RH*** | ***-0,09*** | ***0,26*** | ***Transit*** | ***0,40*** | ***0,00*** |
| | ***None_Yt-1_Transit*** | ***-2936,00*** | ***0,49*** | ***Transit*** | ***0,31*** | ***0,00*** | | | | | | |
| | ***Temp_Yt-1_Transit*** | ***-2936,00*** | ***0,50*** | ***Temp*** | ***-0,11*** | ***0,15*** | ***Transit*** | ***0,34*** | ***0,00*** | | | |
| | ***Temp_RH_Yt-1_Transit*** | ***-2936,00*** | ***0,50*** | ***Temp*** | ***-0,14*** | ***0,09*** | ***RH*** | ***-0,10*** | ***0,21*** | ***Transit*** | ***0,38*** | ***0,00*** |
| | None_Yt-1 | -2924,00 | 0,43 | Yt-1 | 0,77 | 0,00 | | | | | | |
| | Temp_Yt-1 | -2922,00 | 0,43 | Temp | -0,02 | 0,81 | | | | | | |
| | AH_Yt-1 | -2922,00 | 0,43 | AH | -0,03 | 0,70 | | | | | | |
| | AH_RH_Yt-1 | -2921,00 | 0,44 | AH | -0,03 | 0,69 | RH | 0,02 | 0,74 | | | |
| | Temp_RH_Yt-1 | -2920,00 | 0,43 | Temp | -0,02 | 0,84 | RH | 0,02 | 0,77 | | | |
| ***RSV*** | ***AH_Yt-1_Transit*** | ***-2368,00*** | ***0,65*** | ***AH*** | ***-0,13*** | ***0,10*** | ***Transit*** | ***0,39*** | ***0,00*** | | | |
| | ***None_Yt-1_Transit*** | ***-2367,00*** | ***0,65*** | ***Transit*** | ***0,36*** | ***0,00*** | | | | ***precision*** | ***8,61*** | ***0,00*** |
| | ***Temp_Yt-1_Transit*** | ***-2366,00*** | ***0,65*** | ***Temp*** | ***-0,08*** | ***0,29*** | ***Transit*** | ***0,38*** | ***0,00*** | | | |
| | ***AH_RH_Yt-1_Transit*** | ***-2366,00*** | ***0,65*** | ***AH*** | ***-0,13*** | ***0,10*** | ***RH*** | ***0,00*** | ***0,99*** | ***Transit*** | ***0,39*** | ***0,00*** |
| | Temp_RH_Yt-1_Transit | -2364,00 | 0,65 | Temp | -0,08 | 0,30 | RH | -0,02 | 0,83 | Transit | 0,38 | 0,00 |
| | Temp_Yt-1 | -2340,00 | 0,57 | Temp | 0,14 | 0,07 | | | | | | |
| | None_Yt-1 | -2338,00 | 0,56 | Yt-1 | 0,74 | 0,00 | | | | | | |
| | Temp_RH_Yt-1 | -2338,00 | 0,57 | Temp | 0,14 | 0,08 | RH | -0,04 | 0,56 | | | |
| | AH_Yt-1 | -2337,00 | 0,56 | AH | 0,08 | 0,29 | | | | | | |
| | AH_RH_Yt-1 | -2336,00 | 0,56 | AH | 0,08 | 0,27 | RH | -0,06 | 0,43 | | | |
| ***hCoVs*** | ***AH_RH_Yt-1_Transit*** | ***-2354,00*** | ***0,63*** | ***AH*** | ***-0,43*** | ***0,00*** | ***RH*** | ***-0,12*** | ***0,06*** | ***Transit*** | ***0,53*** | ***0,00*** |
| | ***AH_Yt-1_Transit*** | ***-2353,00*** | ***0,62*** | ***AH*** | ***-0,43*** | ***0,00*** | ***Transit*** | ***0,54*** | ***0,00*** | | | |
| | ***Temp_RH_Yt-1_Transit*** | ***-2351,00*** | ***0,62*** | ***Temp*** | ***-0,40*** | ***0,00*** | ***RH*** | ***-0,19*** | ***0,00*** | ***Transit*** | ***0,53*** | ***0,00*** |
| | Temp_Yt-1_Transit | -2345,00 | 0,60 | Temp | -0,36 | 0,00 | Transit | 0,53 | 0,00 | | | |
| | None_Yt-1_Transit | -2330,00 | 0,54 | Transit | 0,37 | 0,00 | | | | | | |
| | None_Yt-1 | -2311,00 | 0,47 | Yt-1 | 0,58 | 0,00 | | | | | | |
| | Temp_RH_Yt-1 | -2311,00 | 0,48 | Temp | -0,06 | 0,50 | RH | -0,13 | 0,06 | | | |
| | AH_RH_Yt-1 | -2311,00 | 0,49 | AH | -0,09 | 0,29 | RH | -0,12 | 0,07 | | | |
| | AH_Yt-1 | -2310,00 | 0,47 | AH | -0,08 | 0,32 | | | | | | |
| | Temp_Yt-1 | -2309,00 | 0,47 | Temp | -0,02 | 0,77 | | | | | | |
| ***IVB*** | ***AH_Yt-1_Transit*** | ***-3423,00*** | ***0,55*** | ***AH*** | ***-0,15*** | ***0,11*** | ***Transit*** | ***0,23*** | ***0,02*** | ***Yt-1*** | ***0,53*** | ***0,00*** |
| | ***None_Yt-1_Transit*** | ***-3422,00*** | ***0,54*** | ***Transit*** | ***0,15*** | ***0,05*** | | | | | | |
| | ***AH_RH_Yt-1_Transit*** | ***-3422,00*** | ***0,56*** | ***AH*** | ***-0,16*** | ***0,09*** | ***RH*** | ***-0,09*** | ***0,29*** | ***Transit*** | ***0,26*** | ***0,01*** |
| | ***Temp_Yt-1_Transit*** | ***-3421,00*** | ***0,55*** | ***Temp*** | ***-0,09*** | ***0,34*** | ***Transit*** | ***0,20*** | ***0,04*** | | | |
| | ***Temp_RH_Yt-1_Transit*** | ***-3421,00*** | ***0,55*** | ***Temp*** | ***-0,11*** | ***0,22*** | ***RH*** | ***-0,11*** | ***0,24*** | ***Transit*** | ***0,24*** | ***0,02*** |
| | ***None_Yt-1*** | ***-3420,00*** | ***0,53*** | ***Yt-1*** | ***0,63*** | ***0,00*** | | | | | | |
| | Temp_Yt-1 | -3418,00 | 0,53 | Temp | 0,02 | 0,84 | | | | | | |
| | AH_Yt-1 | -3418,00 | 0,53 | AH | -0,02 | 0,80 | | | | | | |
| | Temp_RH_Yt-1 | -3416,00 | 0,53 | Temp | 0,02 | 0,84 | RH | -0,01 | 0,90 | | | |
| | AH_RH_Yt-1 | -3416,00 | 0,53 | AH | -0,02 | 0,81 | RH | -0,01 | 0,89 | | | |
| ***hMPV*** | ***AH_Yt-1_Transit*** | ***-2503,00*** | ***0,57*** | ***AH*** | ***-0,24*** | ***0,00*** | ***Transit*** | ***0,50*** | ***0,00*** | | | |
| | ***AH_RH_Yt-1_Transit*** | ***-2501,00*** | ***0,58*** | ***AH*** | ***-0,24*** | ***0,00*** | ***RH*** | ***-0,06*** | ***0,49*** | ***Transit*** | ***0,52*** | ***0,00*** |
| | ***Temp_Yt-1_Transit*** | ***-2500,00*** | ***0,57*** | ***Temp*** | ***-0,22*** | ***0,00*** | ***Transit*** | ***0,49*** | ***0,00*** | | | |
| | Temp_RH_Yt-1_Transit | -2499,00 | 0,57 | Temp | -0,23 | 0,00 | RH | -0,09 | 0,31 | Transit | 0,51 | 0,00 |
| | None_Yt-1_Transit | -2492,00 | 0,53 | Transit | 0,43 | 0,00 | | | | | | |
| | None_Yt-1 | -2475,00 | 0,46 | Yt-1 | 0,64 | 0,00 | | | | | | |
| | Temp_Yt-1 | -2475,00 | 0,47 | Temp | -0,08 | 0,25 | | | | | | |
| | AH_Yt-1 | -2475,00 | 0,47 | AH | -0,09 | 0,23 | | | | | | |
| | Temp_RH_Yt-1 | -2473,00 | 0,47 | Temp | -0,08 | 0,29 | RH | 0,05 | 0,54 | | | |
| | AH_RH_Yt-1 | -2473,00 | 0,47 | AH | -0,09 | 0,23 | RH | 0,06 | 0,45 | | | |

### USA

| Virus | Model | AIC | p_r2 | Var1 | Coef1 | pVal1 | Var2 | Coef2 | pVal2 | Var3 | Coef3 | pVal3 |
|---|---|---|---|---|---|---|---|---|---|---|---|---|
| ***IVA*** | ***AH_Yt-1_Transit*** | ***-2121,00*** | ***0,41*** | ***AH*** | ***-0,25*** | ***0,01*** | ***Transit*** | ***0,18*** | ***0,04*** | | | |
| | ***Temp_Yt-1_Transit*** | ***-2120,00*** | ***0,40*** | ***Temp*** | ***-0,24*** | ***0,02*** | ***Transit*** | ***0,19*** | ***0,05*** | | | |
| | ***Temp_RH_Yt-1_Transit*** | ***-2120,00*** | ***0,41*** | ***Temp*** | ***-0,23*** | ***0,02*** | ***RH*** | ***-0,10*** | ***0,17*** | ***Transit*** | ***0,16*** | ***0,09*** |
| | ***AH_RH_Yt-1_Transit*** | ***-2120,00*** | ***0,41*** | ***AH*** | ***-0,23*** | ***0,03*** | ***RH*** | ***-0,06*** | ***0,50*** | ***Transit*** | ***0,16*** | ***0,10*** |
| | ***AH_Yt-1*** | ***-2119,00*** | ***0,39*** | ***AH*** | ***-0,14*** | ***0,08*** | | | | | | |
| | ***Temp_RH_Yt-1*** | ***-2119,00*** | ***0,40*** | ***Temp*** | ***-0,12*** | ***0,13*** | ***RH*** | ***-0,13*** | ***0,08*** | | | |
| | ***AH_RH_Yt-1*** | ***-2119,00*** | ***0,40*** | ***AH*** | ***-0,12*** | ***0,15*** | ***RH*** | ***-0,11*** | ***0,16*** | | | |
| | ***None_Yt-1*** | ***-2118,00*** | ***0,37*** | | | | | | | | | |
| | ***Temp_Yt-1*** | ***-2118,00*** | ***0,38*** | ***Temp*** | ***-0,11*** | ***0,14*** | | | | | | |
| | None_Yt-1_Transit | -2117,00 | 0,38 | Transit | 0,05 | 0,44 | | | | | | |
| ***RSV*** | ***None_Yt-1_Transit*** | ***-1977,00*** | ***0,80*** | ***Transit*** | ***0,36*** | ***0,00*** | | | | | | |
| | ***Temp_Yt-1_Transit*** | ***-1976,00*** | ***0,80*** | ***Temp*** | ***-0,05*** | ***0,33*** | ***Transit*** | ***0,36*** | ***0,00*** | | | |
| | ***AH_Yt-1_Transit*** | ***-1976,00*** | ***0,80*** | ***AH*** | ***-0,04*** | ***0,36*** | ***Transit*** | ***0,35*** | ***0,00*** | | | |
| | ***Temp_RH_Yt-1_Transit*** | ***-1974,00*** | ***0,80*** | ***Temp*** | ***-0,05*** | ***0,31*** | ***RH*** | ***0,03*** | ***0,59*** | ***Transit*** | ***0,37*** | ***0,00*** |
| | ***AH_RH_Yt-1_Transit*** | ***-1974,00*** | ***0,80*** | ***AH*** | ***-0,05*** | ***0,29*** | ***RH*** | ***0,04*** | ***0,47*** | ***Transit*** | ***0,37*** | ***0,00*** |
| | None_Yt-1 | -1949,00 | 0,75 | | | | | | | | | |
| | Temp_Yt-1 | -1947,00 | 0,75 | Temp | 0,01 | 0,81 | | | | | | |
| | AH_Yt-1 | -1947,00 | 0,75 | AH | -0,01 | 0,87 | | | | | | |
| | Temp_RH_Yt-1 | -1947,00 | 0,75 | Temp | 0,01 | 0,89 | RH | -0,07 | 0,24 | | | |
| | AH_RH_Yt-1 | -1947,00 | 0,75 | AH | 0,00 | 0,99 | RH | -0,07 | 0,25 | | | |
| ***hCoVs*** | ***Temp_RH_Yt-1_Transit*** | ***-2039,00*** | ***0,72*** | ***Temp*** | ***-0,22*** | ***0,00*** | ***RH*** | ***-0,20*** | ***0,00*** | ***Transit*** | ***0,27*** | ***0,00*** |
| | ***AH_RH_Yt-1_Transit*** | ***-2036,00*** | ***0,71*** | ***AH*** | ***-0,21*** | ***0,00*** | ***RH*** | ***-0,16*** | ***0,00*** | ***Transit*** | ***0,26*** | ***0,00*** |
| | AH_Yt-1_Transit | -2029,00 | 0,69 | AH | -0,25 | 0,00 | Transit | 0,30 | 0,00 | | | |
| | Temp_Yt-1_Transit | -2026,00 | 0,69 | Temp | -0,22 | 0,00 | Transit | 0,31 | 0,00 | | | |
| | None_Yt-1_Transit | -2015,00 | 0,66 | Transit | 0,26 | 0,00 | | | | | | |
| | Temp_RH_Yt-1 | -2001,00 | 0,62 | Temp | -0,09 | 0,14 | RH | -0,27 | 0,00 | | | |
| | AH_RH_Yt-1 | -2000,00 | 0,62 | AH | -0,07 | 0,26 | RH | -0,26 | 0,00 | | | |
| | None_Yt-1 | -1983,00 | 0,55 | | | | | | | | | |
| | AH_Yt-1 | -1983,00 | 0,56 | AH | -0,11 | 0,10 | | | | | | |
| | Temp_Yt-1 | -1982,00 | 0,56 | Temp | -0,06 | 0,32 | | | | | | |
| ***IVB*** | ***None_Yt-1_Transit*** | ***-2813,00*** | ***0,90*** | ***Transit*** | ***-0,23*** | ***0,00*** | | | | | | |
| | ***AH_Yt-1_Transit*** | ***-2812,00*** | ***0,90*** | ***AH*** | ***-0,03*** | ***0,44*** | ***Transit*** | ***-0,22*** | ***0,00*** | | | |
| | ***Temp_Yt-1_Transit*** | ***-2811,00*** | ***0,90*** | ***Temp*** | ***-0,02*** | ***0,70*** | ***Transit*** | ***-0,22*** | ***0,00*** | | | |
| | ***AH_RH_Yt-1_Transit*** | ***-2810,00*** | ***0,90*** | ***AH*** | ***-0,04*** | ***0,36*** | ***RH*** | ***0,02*** | ***0,61*** | ***Transit*** | ***-0,21*** | ***0,00*** |
| | Temp_RH_Yt-1_Transit | -2809,00 | 0,90 | Temp | -0,02 | 0,67 | RH | 0,01 | 0,83 | Transit | -0,22 | 0,00 |
| | AH_RH_Yt-1 | -2776,00 | 0,87 | AH | -0,17 | 0,00 | RH | 0,21 | 0,00 | | | |
| | Temp_RH_Yt-1 | -2773,00 | 0,87 | Temp | -0,14 | 0,00 | RH | 0,18 | 0,00 | | | |
| | Temp_Yt-1 | -2755,00 | 0,85 | Temp | -0,15 | 0,00 | | | | | | |
| | AH_Yt-1 | -2751,00 | 0,85 | AH | -0,12 | 0,02 | | | | | | |
| | None_Yt-1 | -2747,00 | 0,84 | | | | | | | | | |
| ***hMPV*** | ***Temp_Yt-1_Transit*** | ***-2104,00*** | ***0,67*** | ***Temp*** | ***-0,23*** | ***0,00*** | ***Transit*** | ***0,43*** | ***0,00*** | | | |
| | ***AH_Yt-1_Transit*** | ***-2104,00*** | ***0,67*** | ***AH*** | ***-0,22*** | ***0,00*** | ***Transit*** | ***0,42*** | ***0,00*** | | | |
| | ***Temp_RH_Yt-1_Transit*** | ***-2103,00*** | ***0,67*** | ***Temp*** | ***-0,22*** | ***0,00*** | ***RH*** | ***-0,04*** | ***0,58*** | ***Transit*** | ***0,42*** | ***0,00*** |
| | ***AH_RH_Yt-1_Transit*** | ***-2102,00*** | ***0,67*** | ***AH*** | ***-0,23*** | ***0,00*** | ***RH*** | ***0,01*** | ***0,87*** | ***Transit*** | ***0,42*** | ***0,00*** |
| | None_Yt-1_Transit | -2097,00 | 0,65 | Transit | 0,39 | 0,00 | | | | | | |
| | None_Yt-1 | -2054,00 | 0,51 | | | | | | | | | |
| | AH_Yt-1 | -2053,00 | 0,51 | AH | -0,03 | 0,72 | | | | | | |
| | Temp_Yt-1 | -2052,00 | 0,51 | Temp | -0,01 | 0,89 | | | | | | |
| | Temp_RH_Yt-1 | -2052,00 | 0,52 | Temp | -0,02 | 0,84 | RH | -0,10 | 0,17 | | | |
| | AH_RH_Yt-1 | -2052,00 | 0,52 | AH | -0,01 | 0,92 | RH | -0,10 | 0,19 | | | |

### S5 Table. Regression models results for the residential time analysis (post-COVID19 pandemic).

Temp, temperature; None, no weather variable; Resid, residential time; p_r2, pseudo $R^2$, Var, variable, Coef, regression coefficient; pVal, p-value.

Only the weather and mobility coefficients are shown.

#### Canada

| Virus | Model | AIC | p_r2 | Var-1 | Coef-1 | pVal-1 | Var-2 | Coef-2 | pVal-2 | Var-3 | Coef-3 | pVal-3 |
|---|---|---|---|---|---|---|---|---|---|---|---|---|
| ***IVA*** | ***AH_Yt-1_Resid*** | ***-2940,00*** | ***0,51*** | ***AH*** | ***-0,33*** | ***0,00*** | ***Residid*** | ***-0,54*** | ***0,00*** | | | |
| | ***AH_RH_Yt-1_Resid*** | ***-2939,00*** | ***0,51*** | ***AH*** | ***-0,35*** | ***0,00*** | ***RH*** | ***-0,07*** | ***0,32*** | ***Residid*** | ***-0,58*** | ***0,00*** |
| | ***Temp_Yt-1_Resid*** | ***-2936,00*** | ***0,49*** | ***Temp*** | ***-0,24*** | ***0,01*** | ***Residid*** | ***-0,44*** | ***0,00*** | | | |
| | Temp_RH_Yt-1_Resid | -2935,00 | 0,50 | Temp | -0,28 | 0,00 | RH | -0,11 | 0,18 | Residid | -0,50 | 0,00 |
| | None_Yt-1_Resid | -2931,00 | 0,47 | Residid | -0,27 | 0,01 | | | | | | |
| | None_Yt-1 | -2924,00 | 0,43 | | | | | | | | | |
| | Temp_Yt-1 | -2922,00 | 0,43 | Temp | -0,02 | 0,81 | | | | | | |
| | AH_Yt-1 | -2922,00 | 0,43 | AH | -0,03 | 0,70 | | | | | | |
| | AH_RH_Yt-1 | -2921,00 | 0,44 | AH | -0,03 | 0,69 | RH | 0,02 | 0,74 | | | |
| | Temp_RH_Yt-1 | -2920,00 | 0,43 | Temp | -0,02 | 0,84 | RH | 0,02 | 0,77 | | | |
| ***RSV*** | ***AH_Yt-1_Resid*** | ***-2381,00*** | ***0,69*** | ***AH*** | ***-0,26*** | ***0,00*** | ***Residid*** | ***-0,60*** | ***0,00*** | | | |
| | ***AH_RH_Yt-1_Resid*** | ***-2379,00*** | ***0,69*** | ***AH*** | ***-0,26*** | ***0,00*** | ***RH*** | ***0,00*** | ***1,00*** | ***Residid*** | ***-0,60*** | ***0,00*** |
| | ***Temp_Yt-1_Resid*** | ***-2378,00*** | ***0,68*** | ***Temp*** | ***-0,23*** | ***0,00*** | ***Residid*** | ***-0,61*** | ***0,00*** | | | |
| | Temp_RH_Yt-1_Resid | -2376,00 | 0,68 | Temp | -0,23 | 0,01 | RH | -0,03 | 0,69 | Residid | -0,60 | 0,00 |
| | None_Yt-1_Resid | -2373,00 | 0,66 | Residid | -0,47 | 0,00 | | | | | | |
| | Temp_Yt-1 | -2340,00 | 0,57 | Temp | 0,14 | 0,07 | | | | | | |
| | None_Yt-1 | -2338,00 | 0,56 | | | | | | | | | |
| | Temp_RH_Yt-1 | -2338,00 | 0,57 | Temp | 0,14 | 0,08 | RH | -0,04 | 0,56 | | | |
| | AH_Yt-1 | -2337,00 | 0,56 | AH | 0,08 | 0,29 | | | | | | |
| | AH_RH_Yt-1 | -2336,00 | 0,56 | AH | 0,08 | 0,27 | RH | -0,06 | 0,43 | | | |
| ***hCoVs*** | ***AH_Yt-1_Resid*** | ***-2356,00*** | ***0,63*** | ***AH*** | ***-0,58*** | ***0,00*** | ***Residid*** | ***-0,77*** | ***0,00*** | | | |
| | ***AH_RH_Yt-1_Resid*** | ***-2355,00*** | ***0,63*** | ***AH*** | ***-0,57*** | ***0,00*** | ***RH*** | ***-0,06*** | ***0,31*** | ***Residid*** | ***-0,73*** | ***0,00*** |
| | Temp_RH_Yt-1_Resid | -2352,00 | 0,62 | Temp | -0,55 | 0,00 | RH | -0,16 | 0,01 | Residid | -0,77 | 0,00 |
| | Temp_Yt-1_Resid | -2349,00 | 0,61 | Temp | -0,54 | 0,00 | Residid | -0,81 | 0,00 | | | |
| | None_Yt-1_Resid | -2325,00 | 0,53 | Residid | -0,35 | 0,00 | | | | | | |
| | None_Yt-1 | -2311,00 | 0,47 | | | | | | | | | |
| | Temp_RH_Yt-1 | -2311,00 | 0,48 | Temp | -0,06 | 0,50 | RH | -0,13 | 0,06 | | | |
| | AH_RH_Yt-1 | -2311,00 | 0,49 | AH | -0,09 | 0,29 | RH | -0,12 | 0,07 | | | |
| | AH_Yt-1 | -2310,00 | 0,47 | AH | -0,08 | 0,32 | | | | | | |
| | Temp_Yt-1 | -2309,00 | 0,47 | Temp | -0,02 | 0,77 | | | | | | |
| ***IVB*** | ***None_Yt-1*** | ***-3420,00*** | ***0,53*** | ***Yt-1*** | ***0,63*** | ***0,00*** | | | | | | |
| | ***None_Yt-1_Resid*** | ***-3418,00*** | ***0,53*** | ***Residid*** | ***-0,03*** | ***0,69*** | | | | | | |
| | ***Temp_Yt-1*** | ***-3418,00*** | ***0,53*** | ***Temp*** | ***0,02*** | ***0,84*** | | | | | | |
| | ***AH_Yt-1*** | ***-3418,00*** | ***0,53*** | ***AH*** | ***-0,02*** | ***0,80*** | | | | | | |
| | ***AH_Yt-1_Resid*** | ***-3417,00*** | ***0,53*** | ***AH*** | ***-0,06*** | ***0,56*** | ***Residid*** | ***-0,06*** | ***0,52*** | | | |
| | Temp_Yt-1_Resid | -3416,00 | 0,53 | Temp | 0,00 | 0,99 | Residid | -0,03 | 0,74 | | | |
| | Temp_RH_Yt-1 | -3416,00 | 0,53 | Temp | 0,02 | 0,84 | RH | -0,01 | 0,90 | | | |
| | AH_RH_Yt-1 | -3416,00 | 0,53 | AH | -0,02 | 0,81 | RH | -0,01 | 0,89 | | | |
| | AH_RH_Yt-1_Resid | -3415,00 | 0,53 | AH | -0,06 | 0,54 | RH | -0,03 | 0,75 | Residid | -0,07 | 0,48 |
| | Temp_RH_Yt-1_Resid | -3414,00 | 0,53 | Temp | -0,01 | 0,94 | RH | -0,02 | 0,79 | Residid | -0,04 | 0,69 |
| ***hMPV*** | ***AH_Yt-1_Resid*** | ***-2493,00*** | ***0,54*** | ***AH*** | ***-0,35*** | ***0,00*** | ***Residid*** | ***-0,54*** | ***0,00*** | | | |
| | ***Temp_Yt-1_Resid*** | ***-2491,00*** | ***0,54*** | ***Temp*** | ***-0,32*** | ***0,00*** | ***Residid*** | ***-0,50*** | ***0,00*** | | | |
| | ***AH_RH_Yt-1_Resid*** | ***-2491,00*** | ***0,54*** | ***AH*** | ***-0,35*** | ***0,00*** | ***RH*** | ***0,00*** | ***0,98*** | ***Residid*** | ***-0,54*** | ***0,00*** |
| | Temp_RH_Yt-1_Resid | -2489,00 | 0,54 | Temp | -0,33 | 0,00 | RH | -0,04 | 0,61 | Residid | -0,52 | 0,00 |
| | None_Yt-1_Resid | -2479,00 | 0,49 | Residid | -0,22 | 0,02 | | | | | | |
| | None_Yt-1 | -2475,00 | 0,46 | Yt-1 | 0,64 | 0,00 | | | | | | |
| | Temp_Yt-1 | -2475,00 | 0,47 | Temp | -0,08 | 0,25 | | | | | | |
| | AH_Yt-1 | -2475,00 | 0,47 | AH | -0,09 | 0,23 | | | | | | |
| | Temp_RH_Yt-1 | -2473,00 | 0,47 | Temp | -0,08 | 0,29 | RH | 0,05 | 0,54 | | | |
| | AH_RH_Yt-1 | -2473,00 | 0,47 | AH | -0,09 | 0,23 | RH | 0,06 | 0,45 | | | |

**Canada**

| Virus | Model | AIC | p_r2 | Var1 | Coef1 | pVal1 | Var2 | Coef2 | pVal2 | Var3 | Coef3 | pVal3 |
|---|---|---|---|---|---|---|---|---|---|---|---|---|
| ***IVA*** | ***AH_Yt-1_Resid*** | ***-2124,00*** | ***0,42*** | ***AH*** | ***-0,24*** | ***0,01*** | ***Resid*** | ***-0,24*** | ***0,01*** | | | |
| | ***Temp_Yt-1_Resid*** | ***-2123,00*** | ***0,42*** | ***Temp*** | ***-0,23*** | ***0,01*** | ***Resid*** | ***-0,25*** | ***0,01*** | | | |
| | ***Temp_RH_Yt-1_Resid*** | ***-2123,00*** | ***0,42*** | ***Temp*** | ***-0,22*** | ***0,01*** | ***RH*** | ***-0,09*** | ***0,23*** | ***Resid*** | ***-0,23*** | ***0,03*** |
| | ***AH_RH_Yt-1_Resid*** | ***-2123,00*** | ***0,42*** | ***AH*** | ***-0,22*** | ***0,02*** | ***RH*** | ***-0,05*** | ***0,57*** | ***Resid*** | ***-0,22*** | ***0,03*** |
| | None_Yt-1_Resid | -2119,00 | 0,39 | Resid | -0,13 | 0,10 | | | | | | |
| | AH_Yt-1 | -2119,00 | 0,39 | AH | -0,14 | 0,08 | | | | | | |
| | Temp_RH_Yt-1 | -2119,00 | 0,40 | Temp | -0,12 | 0,13 | RH | -0,13 | 0,08 | | | |
| | AH_RH_Yt-1 | -2119,00 | 0,40 | AH | -0,12 | 0,15 | RH | -0,11 | 0,16 | | | |
| | None_Yt-1 | -2118,00 | 0,37 | Yt-1 | 0,49 | 0,00 | | | | | | |
| | Temp_Yt-1 | -2118,00 | 0,38 | Temp | -0,11 | 0,14 | | | | | | |
| ***RSV*** | ***None_Yt-1_Resid*** | ***-1969,00*** | ***0,78*** | ***Resid*** | ***-0,33*** | ***0,00*** | | | | | | |
| | ***Temp_Yt-1_Resid*** | ***-1968,00*** | ***0,78*** | ***Temp*** | ***-0,05*** | ***0,41*** | ***Resid*** | ***-0,34*** | ***0,00*** | | | |
| | ***AH_Yt-1_Resid*** | ***-1968,00*** | ***0,78*** | ***AH*** | ***-0,04*** | ***0,45*** | ***Resid*** | ***-0,33*** | ***0,00*** | | | |
| | ***Temp_RH_Yt-1_Resid*** | ***-1966,00*** | ***0,78*** | ***Temp*** | ***-0,05*** | ***0,39*** | ***RH*** | ***0,04*** | ***0,58*** | ***Resid*** | ***-0,36*** | ***0,00*** |
| | ***AH_RH_Yt-1_Resid*** | ***-1966,00*** | ***0,78*** | ***AH*** | ***-0,05*** | ***0,36*** | ***RH*** | ***0,05*** | ***0,48*** | ***Resid*** | ***-0,35*** | ***0,00*** |
| | None_Yt-1 | -1949,00 | 0,75 | Yt-1 | 0,94 | 0,00 | | | | | | |
| | Temp_Yt-1 | -1947,00 | 0,75 | Temp | 0,01 | 0,81 | | | | | | |
| | AH_Yt-1 | -1947,00 | 0,75 | AH | -0,01 | 0,87 | | | | | | |
| | Temp_RH_Yt-1 | -1947,00 | 0,75 | Temp | 0,01 | 0,89 | RH | -0,07 | 0,24 | | | |
| | AH_RH_Yt-1 | -1947,00 | 0,75 | AH | 0,00 | 0,99 | RH | -0,07 | 0,25 | | | |
| ***hCoVs*** | ***Temp_RH_Yt-1_Resid*** | ***-2041,00*** | ***0,72*** | ***Temp*** | ***-0,19*** | ***0,00*** | ***RH*** | ***-0,15*** | ***0,00*** | ***Resid*** | ***-0,30*** | ***0,00*** |
| | ***AH_RH_Yt-1_Resid*** | ***-2038,00*** | ***0,72*** | ***AH*** | ***-0,18*** | ***0,01*** | ***RH*** | ***-0,11*** | ***0,03*** | ***Resid*** | ***-0,29*** | ***0,00*** |
| | AH_Yt-1_Resid | -2036,00 | 0,71 | AH | -0,21 | 0,00 | Resid | -0,34 | 0,00 | | | |
| | Temp_Yt-1_Resid | -2035,00 | 0,71 | Temp | -0,20 | 0,00 | Resid | -0,35 | 0,00 | | | |
| | None_Yt-1_Resid | -2026,00 | 0,68 | Resid | -0,32 | 0,00 | | | | | | |
| | Temp_RH_Yt-1 | -2001,00 | 0,62 | Temp | -0,09 | 0,14 | RH | -0,27 | 0,00 | | | |
| | AH_RH_Yt-1 | -2000,00 | 0,62 | AH | -0,07 | 0,26 | RH | -0,26 | 0,00 | | | |
| | None_Yt-1 | -1983,00 | 0,55 | Yt-1 | 0,53 | 0,00 | | | | | | |
| | AH_Yt-1 | -1983,00 | 0,56 | AH | -0,11 | 0,10 | | | | | | |
| | Temp_Yt-1 | -1982,00 | 0,56 | Temp | -0,06 | 0,32 | | | | | | |
| ***IVB*** | ***AH_Yt-1_Resid*** | ***-2815,00*** | ***0,91*** | ***AH*** | ***-0,08*** | ***0,07*** | ***Resid*** | ***0,22*** | ***0,00*** | | | |
| | ***None_Yt-1_Resid*** | ***-2814,00*** | ***0,90*** | ***Resid*** | ***0,22*** | ***0,00*** | | | | | | |
| | ***Temp_Yt-1_Resid*** | ***-2814,00*** | ***0,90*** | ***Temp*** | ***-0,06*** | ***0,16*** | ***Resid*** | ***0,21*** | ***0,00*** | | | |
| | ***AH_RH_Yt-1_Resid*** | ***-2813,00*** | ***0,91*** | ***AH*** | ***-0,09*** | ***0,06*** | ***RH*** | ***0,02*** | ***0,65*** | ***Resid*** | ***0,21*** | ***0,00*** |
| | ***Temp_RH_Yt-1_Resid*** | ***-2812,00*** | ***0,90*** | ***Temp*** | ***-0,06*** | ***0,16*** | ***RH*** | ***0,00*** | ***0,96*** | ***Resid*** | ***0,21*** | ***0,00*** |
| | AH_RH_Yt-1 | -2776,00 | 0,87 | AH | -0,17 | 0,00 | RH | 0,21 | 0,00 | | | |
| | Temp_RH_Yt-1 | -2773,00 | 0,87 | Temp | -0,14 | 0,00 | RH | 0,18 | 0,00 | | | |
| | Temp_Yt-1 | -2755,00 | 0,85 | Temp | -0,15 | 0,00 | | | | | | |
| | AH_Yt-1 | -2751,00 | 0,85 | AH | -0,12 | 0,02 | | | | | | |
| | None_Yt-1 | -2747,00 | 0,84 | Yt-1 | 0,59 | 0,00 | | | | | | |
| ***hMPV*** | ***Temp_Yt-1_Resid*** | ***-2113,00*** | ***0,69*** | ***Temp*** | ***-0,17*** | ***0,02*** | ***Resid*** | ***-0,52*** | ***0,00*** | | | |
| | ***AH_Yt-1_Resid*** | ***-2113,00*** | ***0,69*** | ***AH*** | ***-0,16*** | ***0,03*** | ***Resid*** | ***-0,51*** | ***0,00*** | | | |
| | ***Temp_RH_Yt-1_Resid*** | ***-2111,00*** | ***0,69*** | ***Temp*** | ***-0,17*** | ***0,02*** | ***RH*** | ***0,01*** | ***0,92*** | ***Resid*** | ***-0,52*** | ***0,00*** |
| | ***AH_RH_Yt-1_Resid*** | ***-2111,00*** | ***0,69*** | ***AH*** | ***-0,17*** | ***0,02*** | ***RH*** | ***0,04*** | ***0,55*** | ***Resid*** | ***-0,52*** | ***0,00*** |
| | ***None_Yt-1_Resid*** | ***-2110,00*** | ***0,68*** | ***Resid*** | ***-0,49*** | ***0,00*** | | | | | | |
| | None_Yt-1 | -2054,00 | 0,51 | Yt-1 | 0,63 | 0,00 | | | | | | |
| | AH_Yt-1 | -2053,00 | 0,51 | AH | -0,03 | 0,72 | | | | | | |
| | Temp_Yt-1 | -2052,00 | 0,51 | Temp | -0,01 | 0,89 | | | | | | |
| | Temp_RH_Yt-1 | -2052,00 | 0,52 | Temp | -0,02 | 0,84 | RH | -0,10 | 0,17 | | | |
| | AH_RH_Yt-1 | -2052,00 | 0,52 | AH | -0,01 | 0,92 | RH | -0,10 | 0,19 | | | |
