## Supplementary tables 1-2 for "The effects of weather and mobility on respiratory viruses dynamics before and after the COVID-19 pandemic"

**S1 Table. Regression models results for the weather analysis for Canada and the USA pre-COVID19 pandemic.**

Temp, temperature; None, no weather variable; p_r2, pseudo $R^2$, Var, variable, Coef, regression coefficient; pVal, p-value.

Only the weather and mobility coefficients are shown.

### Canada

| Virus | Model | AIC | p_r2 | Var-1 | Coef-1 | pVal-1 | Var-2 | Coef-2 | pVal-2 |
|---|---|---|---|---|---|---|---|---|---|
| ***IVA*** | ***AH_RH_Yt-1*** | ***-2400,00*** | ***0,83*** | ***AH*** | ***-0,94*** | ***0,00*** | ***RH*** | ***0,09*** | ***0,02*** |
| | ***AH_Yt-1*** | ***-2397.00*** | ***0,83*** | ***AH*** | ***-0,93*** | ***0,00*** | | | |
| | AH_RH_Yt-1/2 | -2386.00 | 0,84 | AH | -0,92 | 0,00 | RH | 0,10 | 0,01 |
| | Temp_Yt-1 | -2382.00 | 0,81 | Temp | -0,77 | 0,00 | | | |
| | Temp_RH_Yt-1 | -2382.00 | 0,81 | Temp | -0,77 | 0,00 | RH | 0,06 | 0,17 |
| | AH_Yt-1/2 | -2382.00 | 0,83 | AH | -0,90 | 0,00 | | | |
| | Temp_Yt-1/2 | -2367.00 | 0,82 | Temp | -0,75 | 0,00 | | | |
| | Temp_RH_Yt-1/2 | -2367.00 | 0,82 | Temp | -0,75 | 0,00 | RH | 0,07 | 0,12 |
| | Temp | -23009.00 | 0,69 | Temp | -0,99 | 0,00 | | | |
| | Temp_RH | -23009.00 | 0,69 | Temp | -0,99 | 0,00 | RH | 0,06 | 0,24 |
| | AH_RH | -2288.00 | 0,66 | AH | -1,09 | 0,00 | RH | 0,11 | 0,01 |
| | AH | -2284.00 | 0,64 | AH | -1,09 | 0,00 | | | |
| | None_Yt-1 | -2243.00 | 0,59 | | | | | | |
| | None_Yt-1/2 | -2239.00 | 0,62 | | | | | | |
| | None | -20096.00 | 0,00 | | | | | | |
| ***RSV*** | ***AH_RH_Yt-1*** | ***-26300.00*** | ***0,84*** | ***AH*** | ***-0,96*** | ***0,00*** | ***RH*** | ***0,09*** | ***0,01*** |
| | AH_Yt-1 | -2625.00 | 0,84 | AH | -0,94 | 0,00 | | | |
| | AH_RH_Yt-1/2 | -2623.00 | 0,85 | AH | -0,93 | 0,00 | RH | 0,11 | 0,00 |
| | AH_Yt-1/2 | -2616.00 | 0,85 | AH | -0,92 | 0,00 | | | |
| | Temp_Yt-1 | -26009.00 | 0,82 | Temp | -0,78 | 0,00 | | | |
| | Temp_RH_Yt-1 | -26009.00 | 0,82 | Temp | -0,78 | 0,00 | RH | 0,06 | 0,20 |
| | Temp_RH_Yt-1/2 | -2599.00 | 0,83 | Temp | -0,75 | 0,00 | RH | 0,07 | 0,09 |
| | Temp_Yt-1/2 | -2598.00 | 0,83 | Temp | -0,75 | 0,00 | | | |
| | Temp | -2539.00 | 0,71 | Temp | -0,99 | 0,00 | | | |
| | Temp_RH | -2537.00 | 0,71 | Temp | -0,99 | 0,00 | RH | 0,03 | 0,52 |
| | AH_RH | -25200.00 | 0,69 | AH | -1,12 | 0,00 | RH | 0,09 | 0,05 |
| | AH | -2519.00 | 0,68 | AH | -1,12 | 0,00 | | | |
| | None_Yt-1/2 | -2455.00 | 0,62 | | | | | | |
| | None_Yt-1 | -2454.00 | 0,58 | | | | | | |
| | None | -2311.00 | 0,00 | | | | | | |
| ***hCoVs*** | ***AH_Yt-1*** | ***-2755.00*** | ***0,81*** | ***AH*** | ***-0,84*** | ***0,00*** | | | |
| | ***AH_RH_Yt-1*** | ***-2755.00*** | ***0,81*** | ***AH*** | ***-0,85*** | ***0,00*** | ***RH*** | ***-0,06*** | ***0,14*** |
| | Temp_RH_Yt-1 | -2746.00 | 0,80 | Temp | -0,74 | 0,00 | RH | -0,11 | 0,01 |
| | AH_Yt-1/2 | -2744.00 | 0,82 | AH | -0,83 | 0,00 | | | |
| | AH_RH_Yt-1/2 | -2743.00 | 0,82 | AH | -0,84 | 0,00 | RH | -0,04 | 0,28 |
| | Temp_Yt-1 | -2742.00 | 0,79 | Temp | -0,72 | 0,00 | | | |
| | Temp_RH_Yt-1/2 | -2738.00 | 0,81 | Temp | -0,73 | 0,00 | RH | -0,09 | 0,03 |
| | Temp_Yt-1/2 | -2735.00 | 0,81 | Temp | -0,72 | 0,00 | | | |
| | Temp_RH | -2647.00 | 0,62 | Temp | -0,93 | 0,00 | RH | -0,15 | 0,00 |
| | Temp | -26400.00 | 0,60 | Temp | -0,90 | 0,00 | | | |
| | AH | -26200.00 | 0,56 | AH | -0,93 | 0,00 | | | |
| | AH_RH | -26200.00 | 0,56 | AH | -0,95 | 0,00 | RH | -0,08 | 0,11 |
| | None_Yt-1 | -2618.00 | 0,59 | | | | | | |
| | None_Yt-1/2 | -26100.00 | 0,61 | | | | | | |
| | None | -2473.00 | 0,00 | | | | | | |

| Virus | Model | AIC | p_r2 | Var-1 | Coef-1 | pVal-1 | Var-2 | Coef-2 | pVal-2 |
|---|---|---|---|---|---|---|---|---|---|
| ***IVB*** | ***AH_RH_Yt-1*** | ***-2803,00*** | ***0,66*** | ***AH*** | ***-0,53*** | ***0,00*** | ***RH*** | ***-0,13*** | ***0,02*** |
| | ***AH_Yt-1*** | ***-2800,00*** | ***0,65*** | ***AH*** | ***-0,50*** | ***0,00*** | | | |
| | Temp_RH_Yt-1 | -2793,00 | 0,64 | Temp | -0,46 | 0,00 | RH | -0,16 | 0,01 |
| | Temp_Yt-1 | -2789,00 | 0,63 | Temp | -0,42 | 0,00 | | | |
| | AH_RH_Yt-1/2 | -2785,00 | 0,66 | AH | -0,53 | 0,00 | RH | -0,13 | 0,03 |
| | AH_Yt-1/2 | -2783,00 | 0,65 | AH | -0,50 | 0,00 | | | |
| | Temp_RH_Yt-1/2 | -2777,00 | 0,65 | Temp | -0,48 | 0,00 | RH | -0,16 | 0,02 |
| | Temp_Yt-1/2 | -2773,00 | 0,63 | Temp | -0,44 | 0,00 | | | |
| | None_Yt-1 | -2752,00 | 0,54 | | | | | | |
| | None_Yt-1/2 | -2734,00 | 0,54 | | | | | | |
| | AH_RH | -2689,00 | 0,26 | AH | -0,54 | 0,00 | RH | -0,15 | 0,02 |
| | AH | -2686,00 | 0,24 | AH | -0,50 | 0,00 | | | |
| | Temp_RH | -2686,00 | 0,25 | Temp | -0,52 | 0,00 | RH | -0,19 | 0,01 |
| | Temp | -2681,00 | 0,22 | Temp | -0,47 | 0,00 | | | |
| | None | -2637,00 | 0,00 | | | | | | |
| ***hMPV*** | ***AH_RH_Yt-1*** | ***-2804,00*** | ***0,79*** | ***AH*** | ***-0,55*** | ***0,00*** | ***RH*** | ***-0,19*** | ***0,00*** |
| | Temp_RH_Yt-1 | -2787,00 | 0,77 | Temp | -0,46 | 0,00 | RH | -0,25 | 0,00 |
| | AH_RH_Yt-1/2 | -2786,00 | 0,79 | AH | -0,55 | 0,00 | RH | -0,19 | 0,00 |
| | AH_Yt-1 | -2779,00 | 0,76 | AH | -0,49 | 0,00 | | | |
| | Temp_RH_Yt-1/2 | -2770,00 | 0,77 | Temp | -0,47 | 0,00 | RH | -0,24 | 0,00 |
| | AH_Yt-1/2 | -2762,00 | 0,76 | AH | -0,49 | 0,00 | | | |
| | Temp_Yt-1 | -2756,00 | 0,72 | Temp | -0,37 | 0,00 | | | |
| | Temp_Yt-1/2 | -2742,00 | 0,73 | Temp | -0,38 | 0,00 | | | |
| | None_Yt-1 | -2711,00 | 0,64 | | | | | | |
| | None_Yt-1/2 | -2694,00 | 0,65 | | | | | | |
| | AH_RH | -2673,00 | 0,52 | AH | -0,73 | 0,00 | RH | -0,30 | 0,00 |
| | Temp_RH | -2668,00 | 0,51 | Temp | -0,66 | 0,00 | RH | -0,36 | 0,00 |
| | AH | -2637,00 | 0,41 | AH | -0,65 | 0,00 | | | |
| | Temp | -2626,00 | 0,38 | Temp | -0,58 | 0,00 | | | |
| | None | -2540,00 | 0,00 | | | | | | |

### USA

| Virus | Model | AIC | p_r2 | Var-1 | Coef-1 | pVal-1 | Var-2 | Coef-2 | pVal-2 |
|---|---|---|---|---|---|---|---|---|---|
| ***IVA*** | ***AH_RH_Yt-1*** | ***-2866,00*** | ***0,82*** | ***AH*** | ***-0,70*** | ***0,00*** | ***RH*** | ***0,10*** | ***0,00*** |
| | AH_Yt-1 | -2857,00 | 0,81 | AH | -0,66 | 0,00 | | | |
| | AH_RH_Yt-1/2 | -2853,00 | 0,83 | AH | -0,68 | 0,00 | RH | 0,10 | 0,00 |
| | Temp_Yt-1 | -2851,00 | 0,81 | Temp | -0,59 | 0,00 | | | |
| | Temp_RH_Yt-1 | -2850,00 | 0,81 | Temp | -0,59 | 0,00 | RH | 0,03 | 0,30 |
| | AH_Yt-1/2 | -2844,00 | 0,82 | AH | -0,64 | 0,00 | | | |
| | Temp_Yt-1/2 | -2835,00 | 0,81 | Temp | -0,58 | 0,00 | | | |
| | Temp_RH_Yt-1/2 | -2834,00 | 0,81 | Temp | -0,58 | 0,00 | RH | 0,03 | 0,30 |
| | None_Yt-1 | -2748,00 | 0,70 | | | | | | |
| | None_Yt-1/2 | -2738,00 | 0,71 | | | | | | |
| | Temp | -2622,00 | 0,44 | Temp | -0,77 | 0,00 | | | |
| | Temp_RH | -2621,00 | 0,45 | Temp | -0,79 | 0,00 | RH | -0,06 | 0,24 |
| | AH | -2602,00 | 0,39 | AH | -0,74 | 0,00 | | | |
| | AH_RH | -2601,00 | 0,40 | AH | -0,74 | 0,00 | RH | 0,07 | 0,19 |
| | None | -2488,00 | 0,00 | | | | | | |
| ***RSV*** | ***AH_RH_Yt-1*** | ***-2321,00*** | ***0,94*** | ***AH*** | ***-1,08*** | ***0,00*** | ***RH*** | ***0,22*** | ***0,00*** |
| | AH_RH_Yt-1/2 | -2302,00 | 0,94 | AH | -1,08 | 0,00 | RH | 0,21 | 0,00 |
| | Temp_RH_Yt-1 | -2250,00 | 0,91 | Temp | -0,83 | 0,00 | RH | 0,12 | 0,00 |
| | Temp_RH_Yt-1/2 | -2231,00 | 0,91 | Temp | -0,82 | 0,00 | RH | 0,12 | 0,00 |
| | AH_Yt-1 | -2227,00 | 0,90 | AH | -0,86 | 0,00 | | | |
| | Temp_Yt-1 | -2225,00 | 0,90 | Temp | -0,77 | 0,00 | | | |
| | AH_Yt-1/2 | -2211,00 | 0,90 | AH | -0,86 | 0,00 | | | |
| | Temp_Yt-1/2 | -2207,00 | 0,90 | Temp | -0,76 | 0,00 | | | |
| | Temp_RH | -2149,00 | 0,83 | Temp | -1,11 | 0,00 | RH | 0,18 | 0,00 |
| | AH_RH | -2149,00 | 0,83 | AH | -1,32 | 0,00 | RH | 0,32 | 0,00 |
| | Temp | -2121,00 | 0,80 | Temp | -1,09 | 0,00 | | | |
| | AH | -2063,00 | 0,72 | AH | -1,13 | 0,00 | | | |
| | None_Yt-1 | -2043,00 | 0,71 | | | | | | |
| | None_Yt-1/2 | -2028,00 | 0,71 | | | | | | |
| | None | -1844,00 | 0,00 | | | | | | |
| ***hCoVs*** | ***AH_RH_Yt-1*** | ***-3230,00*** | ***0,90*** | ***AH*** | ***-0,81*** | ***0,00*** | ***RH*** | ***0,18*** | ***0,00*** |
| | AH_RH_Yt-1/2 | -3211,00 | 0,90 | AH | -0,82 | 0,00 | RH | 0,18 | 0,00 |
| | Temp_RH_Yt-1 | -3183,00 | 0,87 | Temp | -0,66 | 0,00 | RH | 0,13 | 0,00 |
| | AH_Yt-1 | -3177,00 | 0,86 | AH | -0,67 | 0,00 | | | |
| | Temp_RH_Yt-1/2 | -3165,00 | 0,87 | Temp | -0,68 | 0,00 | RH | 0,12 | 0,00 |
| | Temp_Yt-1 | -3162,00 | 0,85 | Temp | -0,63 | 0,00 | | | |
| | AH_Yt-1/2 | -3157,00 | 0,86 | AH | -0,67 | 0,00 | | | |
| | Temp_Yt-1/2 | -3145,00 | 0,85 | Temp | -0,65 | 0,00 | | | |
| | None_Yt-1 | -3069,00 | 0,76 | | | | | | |
| | None_Yt-1/2 | -3049,00 | 0,76 | | | | | | |
| | Temp_RH | -3046,00 | 0,70 | Temp | -1,05 | 0,00 | RH | 0,09 | 0,01 |
| | Temp | -3042,00 | 0,69 | Temp | -1,05 | 0,00 | | | |
| | AH_RH | -3008,00 | 0,64 | AH | -1,03 | 0,00 | RH | 0,22 | 0,00 |
| | AH | -2983,00 | 0,58 | AH | -0,97 | 0,00 | | | |
| | None | -2817,00 | 0,00 | | | | | | |

| Virus | Model | AIC | p_r2 | Var-1 | Coef-1 | pVal-1 | Var-2 | Coef-2 | pVal-2 |
|---|---|---|---|---|---|---|---|---|---|
| ***IVB*** | ***AH_RH_Yt-1*** | ***-3061,00*** | ***0,70*** | ***AH*** | ***-0,50*** | ***0,00*** | ***RH*** | ***-0,14*** | ***0,00*** |
| | ***Temp_RH_Yt-1*** | ***-3060,00*** | ***0,70*** | ***Temp*** | ***-0,47*** | ***0,00*** | ***RH*** | ***-0,21*** | ***0,00*** |
| | AH_Yt-1 | -3051,00 | 0,68 | AH | -0,51 | 0,00 | | | |
| | AH_RH_Yt-1/2 | -3045,00 | 0,70 | AH | -0,50 | 0,00 | RH | -0,14 | 0,00 |
| | Temp_RH_Yt-1/2 | -3043,00 | 0,70 | Temp | -0,47 | 0,00 | RH | -0,21 | 0,00 |
| | Temp_Yt-1 | -3037,00 | 0,66 | Temp | -0,44 | 0,00 | | | |
| | AH_Yt-1/2 | -3037,00 | 0,69 | AH | -0,50 | 0,00 | | | |
| | Temp_Yt-1/2 | -3024,00 | 0,67 | Temp | -0,43 | 0,00 | | | |
| | None_Yt-1 | -2973,00 | 0,55 | | | | | | |
| | None_Yt-1/2 | -2964,00 | 0,57 | | | | | | |
| | Temp_RH | -2868,00 | 0,25 | Temp | -0,47 | 0,00 | RH | -0,19 | 0,00 |
| | AH_RH | -2868,00 | 0,25 | AH | -0,48 | 0,00 | RH | -0,11 | 0,05 |
| | AH | -2866,00 | 0,23 | AH | -0,48 | 0,00 | | | |
| | Temp | -2859,00 | 0,21 | Temp | -0,43 | 0,00 | | | |
| | None | -2807,00 | 0,00 | | | | | | |
| ***hMPV*** | ***AH_Yt-1*** | ***-2425,00*** | ***0,88*** | ***AH*** | ***-0,58*** | ***0,00*** | | | |
| | ***AH_RH_Yt-1*** | ***-2423,00*** | ***0,88*** | ***AH*** | ***-0,58*** | ***0,00*** | ***RH*** | ***0,01*** | ***0,67*** |
| | Temp_RH_Yt-1 | -2407,00 | 0,86 | Temp | -0,49 | 0,00 | RH | -0,06 | 0,03 |
| | Temp_Yt-1 | -2404,00 | 0,86 | Temp | -0,49 | 0,00 | | | |
| | AH_Yt-1/2 | -2393,00 | 0,88 | AH | -0,58 | 0,00 | | | |
| | AH_RH_Yt-1/2 | -2392,00 | 0,88 | AH | -0,59 | 0,00 | RH | 0,02 | 0,57 |
| | Temp_RH_Yt-1/2 | -2377,00 | 0,87 | Temp | -0,50 | 0,00 | RH | -0,05 | 0,06 |
| | Temp_Yt-1/2 | -2376,00 | 0,86 | Temp | -0,51 | 0,00 | | | |
| | None_Yt-1 | -2311,00 | 0,76 | | | | | | |
| | None_Yt-1/2 | -2281,00 | 0,76 | | | | | | |
| | Temp_RH | -2218,00 | 0,52 | Temp | -0,71 | 0,00 | RH | -0,15 | 0,00 |
| | AH | -2215,00 | 0,50 | AH | -0,75 | 0,00 | | | |
| | AH_RH | -2214,00 | 0,51 | AH | -0,75 | 0,00 | RH | -0,06 | 0,27 |
| | Temp | -2212,00 | 0,50 | Temp | -0,69 | 0,00 | | | |
| | None | -2095,00 | 0,00 | | | | | | |

**S2 Table. Regression models results for the weather analysis for Canada and the USA post_COVID19 pandemic**

Temp, temperature; None, no weather variable; p_r2, pseudo $R^2$, Var, variable, Coef, regression coefficient; pVal, p-value.

Only the weather and mobility coefficients are shown.

### Canada

| Virus | Model | AIC | p_r2 | Var-1 | Coef-1 | pVal-1 | Var-2 | Coef-2 | pVal-2 |
|---|---|---|---|---|---|---|---|---|---|
| ***IVA*** | ***None_Yt-1*** | ***-2991,00*** | ***0,40*** | | | | | | |
| | ***Temp_Yt-1*** | ***-2990,00*** | ***0,41*** | ***Temp*** | ***-0,06*** | ***0,49*** | ***Yt-1*** | ***0,69*** | ***0,00*** |
| | ***AH_Yt-1*** | ***-2990,00*** | ***0,41*** | ***AH*** | ***-0,07*** | ***0,35*** | ***Yt-1*** | ***0,68*** | ***0,00*** |
| | ***None_Yt-1/2*** | ***-2989,00*** | ***0,40*** | | | | | | |
| | ***Temp_RH_Yt-1*** | ***-2988,00*** | ***0,41*** | ***Temp*** | ***-0,05*** | ***0,54*** | ***RH*** | ***0,04*** | ***0,63*** |
| | ***AH_RH_Yt-1*** | ***-2988,00*** | ***0,41*** | ***AH*** | ***-0,07*** | ***0,35*** | ***RH*** | ***0,04*** | ***0,57*** |
| | ***Temp_Yt-1/2*** | ***-2988,00*** | ***0,41*** | ***Temp*** | ***-0,06*** | ***0,48*** | | | |
| | ***AH_Yt-1/2*** | ***-2988,00*** | ***0,41*** | ***AH*** | ***-0,07*** | ***0,34*** | | | |
| | Temp_RH_Yt-1/2 | -2986,00 | 0,41 | Temp | -0,05 | 0,54 | RH | 0,04 | 0,62 |
| | AH_RH_Yt-1/2 | -2986,00 | 0,41 | AH | -0,07 | 0,35 | RH | 0,05 | 0,55 |
| | None | -2916,00 | 0,00 | | | | | | |
| | Temp | -2916,00 | 0,01 | Temp | -0,12 | 0,14 | | | |
| | AH | -2916,00 | 0,02 | AH | -0,12 | 0,13 | | | |
| | Temp_RH | -2915,00 | 0,02 | Temp | -0,10 | 0,19 | RH | 0,07 | 0,36 |
| | AH_RH | -2915,00 | 0,02 | AH | -0,12 | 0,13 | RH | 0,09 | 0,26 |
| ***RSV*** | ***None_Yt-1*** | ***-2467,00*** | ***0,61*** | | | | | | |
| | ***Temp_Yt-1*** | ***-2465,00*** | ***0,61*** | ***Temp*** | ***0,02*** | ***0,76*** | | | |
| | ***AH_Yt-1*** | ***-2465,00*** | ***0,61*** | ***AH*** | ***0,01*** | ***0,92*** | | | |
| | ***None_Yt-1/2*** | ***-2465,00*** | ***0,61*** | | | | | | |
| | Temp_RH_Yt-1 | -2463,00 | 0,61 | Temp | 0,02 | 0,83 | RH | -0,04 | 0,59 |
| | AH_RH_Yt-1 | -2463,00 | 0,61 | AH | 0,01 | 0,93 | RH | -0,04 | 0,56 |
| | Temp_Yt-1/2 | -2463,00 | 0,61 | Temp | 0,03 | 0,73 | | | |
| | AH_Yt-1/2 | -2463,00 | 0,61 | AH | 0,01 | 0,90 | | | |
| | Temp_RH_Yt-1/2 | -2461,00 | 0,61 | Temp | 0,02 | 0,81 | RH | -0,04 | 0,58 |
| | AH_RH_Yt-1/2 | -2461,00 | 0,61 | AH | 0,01 | 0,90 | RH | -0,04 | 0,55 |
| | Temp_RH | -2331,00 | 0,05 | Temp | -0,13 | 0,08 | RH | 0,13 | 0,08 |
| | AH_RH | -2331,00 | 0,05 | AH | -0,14 | 0,07 | RH | 0,15 | 0,05 |
| | Temp | -2330,00 | 0,03 | Temp | -0,16 | 0,03 | | | |
| | AH | -2329,00 | 0,03 | AH | -0,15 | 0,05 | | | |
| | None | -2327,00 | 0,00 | | | | | | |
| ***hCoVs*** | ***None_Yt-1*** | ***-2484,00*** | ***0,55*** | | | | | | |
| | ***AH_Yt-1*** | ***-2484,00*** | ***0,56*** | ***AH*** | ***-0,10*** | ***0,18*** | | | |
| | ***None_Yt-1/2*** | ***-2484,00*** | ***0,56*** | | | | | | |
| | ***AH_Yt-1/2*** | ***-2484,00*** | ***0,56*** | ***AH*** | ***-0,09*** | ***0,25*** | | | |
| | ***Temp_Yt-1*** | ***-2482,00*** | ***0,55*** | ***Temp*** | ***-0,04*** | ***0,62*** | | | |
| | ***AH_RH_Yt-1*** | ***-2482,00*** | ***0,56*** | ***AH*** | ***-0,11*** | ***0,17*** | ***RH*** | ***-0,04*** | ***0,54*** |
| | ***Temp_Yt-1/2*** | ***-2482,00*** | ***0,56*** | ***Temp*** | ***-0,02*** | ***0,83*** | | | |
| | ***AH_RH_Yt-1/2*** | ***-2482,00*** | ***0,57*** | ***AH*** | ***-0,09*** | ***0,23*** | ***RH*** | ***-0,05*** | ***0,48*** |
| | ***Temp_RH_Yt-1*** | ***-2481,00*** | ***0,55*** | ***Temp*** | ***-0,05*** | ***0,53*** | ***RH*** | ***-0,04*** | ***0,53*** |
| | ***Temp_RH_Yt-1/2*** | ***-2481,00*** | ***0,56*** | ***Temp*** | ***-0,03*** | ***0,72*** | ***RH*** | ***-0,05*** | ***0,50*** |
| | Temp | -2375,00 | 0,08 | Temp | -0,27 | 0,00 | | | |
| | AH | -2374,00 | 0,07 | AH | -0,25 | 0,00 | | | |
| | Temp_RH | -2373,00 | 0,08 | Temp | -0,28 | 0,00 | RH | -0,04 | 0,59 |
| | AH_RH | -2372,00 | 0,07 | AH | -0,26 | 0,00 | RH | -0,01 | 0,93 |
| | None | -2365,00 | 0,00 | | | | | | |

| Virus | Model | AIC | p_r2 | Var-1 | Coef-1 | pVal-1 | Var-2 | Coef-2 | pVal-2 |
|---|---|---|---|---|---|---|---|---|---|
| ***IVB*** | ***None_Yt-1/2*** | ***-3695,00*** | ***0,54*** | | | | | | |
| | ***AH_Yt-1/2*** | ***-3694,00*** | ***0,54*** | ***AH*** | ***-0,08*** | ***0,27*** | | | |
| | ***AH_RH_Yt-1/2*** | ***-3694,00*** | ***0,55*** | ***AH*** | ***-0,08*** | ***0,26*** | ***RH*** | ***0,10*** | ***0,22*** |
| | ***Temp_Yt-1/2*** | ***-3693,00*** | ***0,54*** | ***Temp*** | ***-0,05*** | ***0,46*** | | | |
| | ***Temp_RH_Yt-1/2*** | ***-3693,00*** | ***0,54*** | ***Temp*** | ***-0,05*** | ***0,54*** | ***RH*** | ***0,09*** | ***0,26*** |
| | None_Yt-1 | -3678,00 | 0,48 | | | | | | |
| | AH_Yt-1 | -3677,00 | 0,48 | AH | -0,08 | 0,26 | | | |
| | AH_RH_Yt-1 | -3677,00 | 0,49 | AH | -0,08 | 0,26 | RH | 0,10 | 0,19 |
| | Temp_Yt-1 | -3676,00 | 0,48 | Temp | -0,06 | 0,45 | | | |
| | Temp_RH_Yt-1 | -3676,00 | 0,48 | Temp | -0,04 | 0,54 | RH | 0,10 | 0,22 |
| | None | -3583,00 | 0,00 | | | 0,00 | | | |
| | Temp | -3582,00 | 0,01 | Temp | -0,07 | 0,38 | | | |
| | AH | -3582,00 | 0,01 | AH | -0,09 | 0,26 | | | |
| | Temp_RH | -3580,00 | 0,01 | Temp | -0,06 | 0,44 | RH | 0,04 | 0,63 |
| | AH_RH | -3580,00 | 0,01 | AH | -0,09 | 0,27 | RH | 0,05 | 0,57 |
| **hMPV** | **AH_RH_Yt-1/2** | **-2682,00** | **0,61** | **AH** | **-0,07** | **0,21** | **RH** | **0,16** | **0,02** |
| | **Temp_RH_Yt-1/2** | **-2681,00** | **0,61** | **Temp** | **-0,06** | **0,35** | **RH** | **0,16** | **0,02** |
| | Temp_Yt-1/2 | -2678,00 | 0,60 | Temp | -0,09 | 0,13 | | | |
| | AH_Yt-1/2 | -2678,00 | 0,60 | AH | -0,09 | 0,12 | | | |
| | None_Yt-1/2 | -2677,00 | 0,59 | | | | | | |
| | AH_RH_Yt-1 | -2663,00 | 0,55 | AH | -0,13 | 0,04 | RH | 0,15 | 0,03 |
| | Temp_RH_Yt-1 | -2662,00 | 0,55 | Temp | -0,12 | 0,07 | RH | 0,14 | 0,05 |
| | Temp_Yt-1 | -2660,00 | 0,54 | Temp | -0,15 | 0,02 | | | |
| | AH_Yt-1 | -2660,00 | 0,54 | AH | -0,15 | 0,02 | | | |
| | None_Yt-1 | -2656,00 | 0,52 | | | | | | |
| | None | -2548,00 | 0,00 | | | | | | |
| | Temp | -2548,00 | 0,02 | Temp | -0,12 | 0,12 | | | |
| | AH | -2548,00 | 0,01 | AH | -0,11 | 0,15 | | | |
| | Temp_RH | -2547,00 | 0,02 | Temp | -0,11 | 0,17 | RH | 0,05 | 0,51 |
| | AH_RH | -2547,00 | 0,02 | AH | -0,11 | 0,16 | RH | 0,06 | 0,39 |

### USA

| Virus | Model | AIC | p_r2 | Var-1 | Coef-1 | pVal-1 | Var-2 | Coef-2 | pVal-2 |
|---|---|---|---|---|---|---|---|---|---|
| ***IVA*** | ***AH_Yt-1*** | ***-2180,00*** | ***0,51*** | ***AH*** | ***-0,17*** | ***0,03*** | | | |
| | ***AH_RH_Yt-1*** | ***-2180,00*** | ***0,52*** | ***AH*** | ***-0,15*** | ***0,05*** | ***RH*** | ***-0,09*** | ***0,23*** |
| | ***Temp_Yt-1*** | ***-2179,00*** | ***0,51*** | ***Temp*** | ***-0,14*** | ***0,06*** | | | |
| | ***Temp_RH_Yt-1*** | ***-2179,00*** | ***0,51*** | ***Temp*** | ***-0,14*** | ***0,06*** | ***RH*** | ***-0,12*** | ***0,10*** |
| | ***AH_Yt-1/2*** | ***-2179,00*** | ***0,51*** | ***AH*** | ***-0,16*** | ***0,03*** | | | |
| | ***AH_RH_Yt-1/2*** | ***-2179,00*** | ***0,52*** | ***AH*** | ***-0,14*** | ***0,08*** | ***RH*** | ***-0,11*** | ***0,15*** |
| | ***Temp_RH_Yt-1/2*** | ***-2178,00*** | ***0,52*** | ***Temp*** | ***-0,13*** | ***0,10*** | ***RH*** | ***-0,13*** | ***0,07*** |
| | ***None_Yt-1*** | ***-2177,00*** | ***0,49*** | | | | | | |
| | ***Temp_Yt-1/2*** | ***-2177,00*** | ***0,51*** | ***Temp*** | ***-0,13*** | ***0,07*** | | | |
| | None_Yt-1/2 | -2176,00 | 0,50 | | | | | | |
| | Temp | -2082,00 | 0,04 | Temp | -0,20 | 0,01 | | | |
| | AH | -2081,00 | 0,04 | AH | -0,20 | 0,01 | | | |
| | Temp_RH | -2080,00 | 0,05 | Temp | -0,20 | 0,01 | RH | 0,03 | 0,74 |
| | AH_RH | -2080,00 | 0,05 | AH | -0,21 | 0,01 | RH | 0,07 | 0,36 |
| | None | -2077,00 | 0,00 | | | | | | |
| ***RSV*** | ***Temp_Yt-1*** | ***-1956,00*** | ***0,57*** | ***Temp*** | ***0,16*** | ***0,02*** | | | |
| | ***AH_Yt-1*** | ***-1956,00*** | ***0,57*** | ***AH*** | ***0,16*** | ***0,02*** | | | |
| | ***Temp_RH_Yt-1*** | ***-1955,00*** | ***0,58*** | ***Temp*** | ***0,17*** | ***0,01*** | ***RH*** | ***0,08*** | ***0,22*** |
| | ***AH_RH_Yt-1*** | ***-1954,00*** | ***0,57*** | ***AH*** | ***0,15*** | ***0,02*** | ***RH*** | ***0,05*** | ***0,45*** |
| | ***Temp_Yt-1/2*** | ***-1954,00*** | ***0,57*** | ***Temp*** | ***0,18*** | ***0,01*** | | | |
| | ***AH_Yt-1/2*** | ***-1954,00*** | ***0,57*** | ***AH*** | ***0,17*** | ***0,01*** | | | |
| | ***Temp_RH_Yt-1/2*** | ***-1954,00*** | ***0,58*** | ***Temp*** | ***0,18*** | ***0,01*** | ***RH*** | ***0,08*** | ***0,24*** |
| | ***AH_RH_Yt-1/2*** | ***-1953,00*** | ***0,58*** | ***AH*** | ***0,16*** | ***0,02*** | ***RH*** | ***0,05*** | ***0,50*** |
| | None_Yt-1 | -1952,00 | 0,56 | | | | | | |
| | None_Yt-1/2 | -1950,00 | 0,56 | | | | | | |
| | None | -1832,00 | 0,00 | | | | | | |
| | Temp_RH | -1831,00 | 0,02 | Temp | 0,05 | 0,53 | RH | 0,14 | 0,09 |
| | AH_RH | -1831,00 | 0,02 | AH | 0,04 | 0,61 | RH | 0,13 | 0,11 |
| | Temp | -1830,00 | 0,00 | Temp | 0,03 | 0,72 | | | |
| | AH | -1830,00 | 0,00 | AH | 0,04 | 0,56 | | | |
| ***hCoVs*** | ***Temp_RH_Yt-1*** | ***-2583,00*** | ***0,65*** | ***Temp*** | ***-0,16*** | ***0,01*** | ***RH*** | ***-0,20*** | ***0,00*** |
| | ***AH_RH_Yt-1*** | ***-2581,00*** | ***0,65*** | ***AH*** | ***-0,14*** | ***0,02*** | ***RH*** | ***-0,17*** | ***0,00*** |
| | ***Temp_RH_Yt-1/2*** | ***-2581,00*** | ***0,65*** | ***Temp*** | ***-0,15*** | ***0,01*** | ***RH*** | ***-0,20*** | ***0,00*** |
| | AH_RH_Yt-1/2 | -2579,00 | 0,65 | AH | -0,14 | 0,02 | RH | -0,17 | 0,00 |
| | AH_Yt-1 | -2573,00 | 0,62 | AH | -0,15 | 0,01 | | | |
| | Temp_Yt-1 | -2571,00 | 0,62 | Temp | -0,12 | 0,04 | | | |
| | AH_Yt-1/2 | -2571,00 | 0,62 | AH | -0,15 | 0,01 | | | |
| | Temp_Yt-1/2 | -2570,00 | 0,62 | Temp | -0,12 | 0,04 | | | |
| | None_Yt-1 | -2569,00 | 0,60 | | | | | | |
| | None_Yt-1/2 | -2567,00 | 0,61 | | | | | | |
| | AH | -2438,00 | 0,05 | AH | -0,20 | 0,00 | | | |
| | Temp | -2437,00 | 0,05 | Temp | -0,19 | 0,01 | | | |
| | Temp_RH | -2437,00 | 0,06 | Temp | -0,20 | 0,00 | RH | -0,09 | 0,25 |
| | AH_RH | -2436,00 | 0,06 | AH | -0,20 | 0,00 | RH | -0,04 | 0,58 |
| | None | -2432,00 | 0,00 | | | | | | |

| Virus | Model | AIC | p_r2 | Var-1 | Coef-1 | pVal-1 | Var-2 | Coef-2 | pVal-2 |
|---|---|---|---|---|---|---|---|---|---|
| ***IVB*** | ***AH_RH_Yt-1/2*** | ***-3011,00*** | ***0,85*** | ***AH*** | ***-0,23*** | ***0,00*** | ***RH*** | ***0,25*** | ***0,00*** |
| | Temp_RH_Yt-1/2 | -3007,00 | 0,85 | Temp | -0,20 | 0,00 | RH | 0,21 | 0,00 |
| | AH_RH_Yt-1 | -2995,00 | 0,83 | AH | -0,22 | 0,00 | RH | 0,24 | 0,00 |
| | Temp_RH_Yt-1 | -2991,00 | 0,83 | Temp | -0,19 | 0,00 | RH | 0,20 | 0,00 |
| | Temp_Yt-1/2 | -2984,00 | 0,82 | Temp | -0,22 | 0,00 | | | |
| | AH_Yt-1/2 | -2978,00 | 0,81 | AH | -0,18 | 0,00 | | | |
| | Temp_Yt-1 | -2971,00 | 0,80 | Temp | -0,22 | 0,00 | | | |
| | None_Yt-1/2 | -2967,00 | 0,79 | | | | | | |
| | AH_Yt-1 | -2965,00 | 0,79 | AH | -0,18 | 0,00 | | | |
| | None_Yt-1 | -2956,00 | 0,77 | | | | | | |
| | AH_RH | -2738,00 | 0,05 | AH | -0,15 | 0,03 | RH | 0,15 | 0,03 |
| | Temp_RH | -2737,00 | 0,04 | Temp | -0,13 | 0,05 | RH | 0,12 | 0,09 |
| | Temp | -2736,00 | 0,03 | Temp | -0,14 | 0,04 | | | |
| | AH | -2736,00 | 0,02 | AH | -0,13 | 0,07 | | | |
| | None | -2734,00 | 0,00 | | | | | | |
| ***hMPV*** | ***None_Yt-1*** | ***-2203,00*** | ***0,52*** | | | | | | |
| | ***Temp_Yt-1*** | ***-2202,00*** | ***0,53*** | ***Temp*** | ***-0,09*** | ***0,22*** | | | |
| | ***AH_Yt-1*** | ***-2202,00*** | ***0,53*** | ***AH*** | ***-0,09*** | ***0,21*** | | | |
| | ***None_Yt-1/2*** | ***-2201,00*** | ***0,52*** | | | | | | |
| | ***Temp_RH_Yt-1*** | ***-2200,00*** | ***0,53*** | ***Temp*** | ***-0,09*** | ***0,20*** | ***RH*** | ***-0,03*** | ***0,65*** |
| | ***AH_RH_Yt-1*** | ***-2200,00*** | ***0,53*** | ***AH*** | ***-0,09*** | ***0,22*** | ***RH*** | ***-0,01*** | ***0,87*** |
| | ***Temp_Yt-1/2*** | ***-2200,00*** | ***0,53*** | ***Temp*** | ***-0,09*** | ***0,22*** | | | |
| | AH_Yt-1/2 | -2200,00 | 0,53 | AH | -0,09 | 0,22 | | | |
| | Temp_RH_Yt-1/2 | -2198,00 | 0,53 | Temp | -0,09 | 0,20 | RH | -0,03 | 0,66 |
| | AH_RH_Yt-1/2 | -2198,00 | 0,53 | AH | -0,09 | 0,22 | RH | -0,01 | 0,87 |
| | Temp | -2096,00 | 0,03 | Temp | -0,14 | 0,05 | | | |
| | AH | -2095,00 | 0,02 | AH | -0,13 | 0,07 | | | |
| | Temp_RH | -2095,00 | 0,03 | Temp | -0,13 | 0,06 | RH | 0,07 | 0,35 |
| | AH_RH | -2095,00 | 0,03 | AH | -0,14 | 0,05 | RH | 0,11 | 0,17 |
| | None | -2094,00 | 0,00 | | | | | | |
